# Potassium Channel Regulation Dysfunction as a Potential Driving Factor of Membrane Potential Disturbance in Restless Legs Syndrome

**DOI:** 10.1101/2025.07.17.25331613

**Authors:** Xin Yao

## Abstract

**Background:** Restless legs syndrome (RLS) is a common neurological disorder whose pathophysiology remains poorly understood. Previous neuroelectrophysiological studies have found a widespread increase in neural excitability. Potassium channels are fundamental determinants of neuronal excitability through their control of resting membrane potential and firing threshold. Mendelian randomization (MR) analysis provides a powerful method for studying the potential relationship between the regulatory function of potassium channels and RLS.

**Methods:** In the exploration phase, based on MR analysis and enrichment analysis results of opioids use, cerebrospinal fluid (CSF) and brain metabolites, and different RLS datasets, it is proposed that potassium channels may play a potential role in RLS. Subsequently, cis-MR across different RLS datasets based on *KCN*-genes OpenGWAS eQTLs and brain single cell eQTLs (sc-eQTLs) were used for further discovery. Using data from external experiments, differential expression analysis of *KCN*-genes was performed in human neural stem cells with *MEIS1* gene knockout and overexpression. In addition, through assuming the regulation of potassium channels in the opioids treatment and the role of mediating factors, an exploratory simulation model was designed to simulate the dynamic relationship and explore the potential influencing factors between opioids and RLS.

**Results:** This study conducted exploratory MR analysis between the genome-wide association study (GWAS) data for opioids use and RLS, and four candidate mediators were found and evaluated in mediation analysis using 1402 CSF and brain metabolites as mediators. Enrichment analyses across different RLS datasets of genes mapped from instrumental variable linked to RLS-related candidate metabolites showed, the mechanism of RLS may be related to potassium channels, membrane potential regulation, synaptic function and addiction. Based on these clues, further exploration was conducted on the causal relationship between potassium channels and RLS. eQTLs of *KCN*-designated genes (*KCN*-genes) were obtained from the OpenGWAS website and subjected to single gene cis-MR analysis, and the results showed that 13 *KCN*-genes were significantly correlations with RLS. The further analysis based on sc-eQTLs supports higher expression of *KCNAB1* gene in oligodendrocytes and *KCNMA1* gene in microglia were associated with higher RLS risk. Differential expression analysis of *KCN*-genes between *MEIS1* gene knockout and overexpression showed the regulatory mode of *MEIS1* on different *KCN*-genes.

**Conclusion:** This study provides the genetic support and preliminary multi method evaluation for understanding the relationship between potassium channel regulation and RLS, and proposes “Membrane Potential Disturbance” hypothesis, that may suggest a new perspective and framework for understanding the pathophysiology of RLS.

## Introduction

Restless legs syndrome (RLS) affects approximately 7.12% of the global population (95% CI = 5.15–9.76), equating to 356.07 million affected individuals worldwide, and represents one of the most common neurological disorders^1^. Despite extensive research, its pathophysiology remains poorly understood; RLS is currently defined only by clinical diagnostic criteria. Existing hypotheses focus on brain iron deficiency and dopamine dysregulation, but neither fully explains RLS^2^. For example, cerebrospinal fluid (CSF) levels of 3-Ortho-methyldopa (a dopamine metabolite) are elevated in RLS patients and positively correlated with disease severity, suggesting dopamine synthesis hyperactivity^3^. Additionally, imaging studies and autopsy findings indicate concurrent secondary dopamine receptor downregulation^4^. These observations contradict simple dopamine-deficiency models.

Potassium channels locate in cell membranes and widely distributed^5^. For neuronal cells, potassium channels maintain homeostasis and regulate neuronal firing properties and membrane potential^6^, and participate in the regulation and plasticity of synaptic function^7^. Potassium channels also are fundamental determinants of neuronal excitability through their control of resting membrane potential and firing threshold. Reduced potassium channel function shifts the resting potential toward depolarization, lowering the threshold for action potential initiation and converting neurons from Type II (high-threshold) to Type I (low-threshold) excitability patterns^8^.

Notably, CSF and autopsy studies in RLS patients have found a deficiency of endogenous opioid^9,10^. Consistent with this, mice with knockout opioid receptors exhibit RLS-like symptoms^11^. Clinically, opioids have proven effective: a double-blind, randomised trial showed that controlled-release oxycodone–naloxone produced significant and sustained improvement in severe RLS^12^, and long-term follow-ups reported maintained efficacy without notable tolerance^13,14^. Opioid receptors are known to modulate various potassium channels^15^: regulation of voltage-gated potassium (*Kv*) channels plays a pivotal role in the analgesic, reward-related, addictive, and neuroplastic effects of opioids, exhibiting pronounced brain region-and cell type-specificity. In contrast, activation of G protein–gated inwardly rectifying potassium (*GIRK*) channels produces rapid and robust neuronal hyperpolarization. It is thus biologically plausible that opioids improve RLS by potassium channel regulation. However, due to concerns about adverse drug reactions, tolerance and addiction issues, the opioids use is still limited. The pathogenesis of RLS and the dynamic evolution mechanism of opioids use are unclear, that makes it difficult to carry out effective stratified and individualized treatment, which may further increase concerns about long-term opioids use.

Recent advances in Mendelian randomization (MR) methodology and large-scale GWAS data enable investigation of causal relationships between exposures and outcomes^16^. Moreover, CSF and brain metabolites directly reflect brain biochemical states, making them valuable mediators for understanding complex neurological disorders. This study conducted a three-stage study design (**Fig. 1**):

1. Exploration phase: By investigating the dynamic relationship between opioids use and RLS, incorporating simulation-time-heterogeneous mediating effects of different mediators may enable a more biologically plausible simulation of real-world clinical dynamics. Enrichment analyses showed significant enrichment of pathways associated with potassium channel and membrane potential regulation, suggesting as potential role closely related to an underlying mechanism in RLS.
2. Discovery phase: The cis-Mendelian randomization (cis-MR) analysis^17^ based on OpenGWAS eQTLs supported the significant association between 13 potassium channel-related genes (*KCN*-genes) and RLS. Subsequently, sc-eQTL cis-MR analysis was used to validate, and the result showed upregulation of *KCNAB1* gene in oligodendrocytes and *KCNMA1* gene in microglia.
3. Hypothesis proposed: These findings combined with existing study evidence, led to the proposal of a novel potential pathophysiological framework—“Membrane Potential Disturbance” hypothesis—which suggests membrane potential abnormalities act as a central link between diverse upstream etiologies and downstream similar mechanisms/symptoms, and a potential unified explanation for RLS manifestations, iron metabolism disorders, dopaminergic system change, and therapeutic responses.

**Fig. 1.**
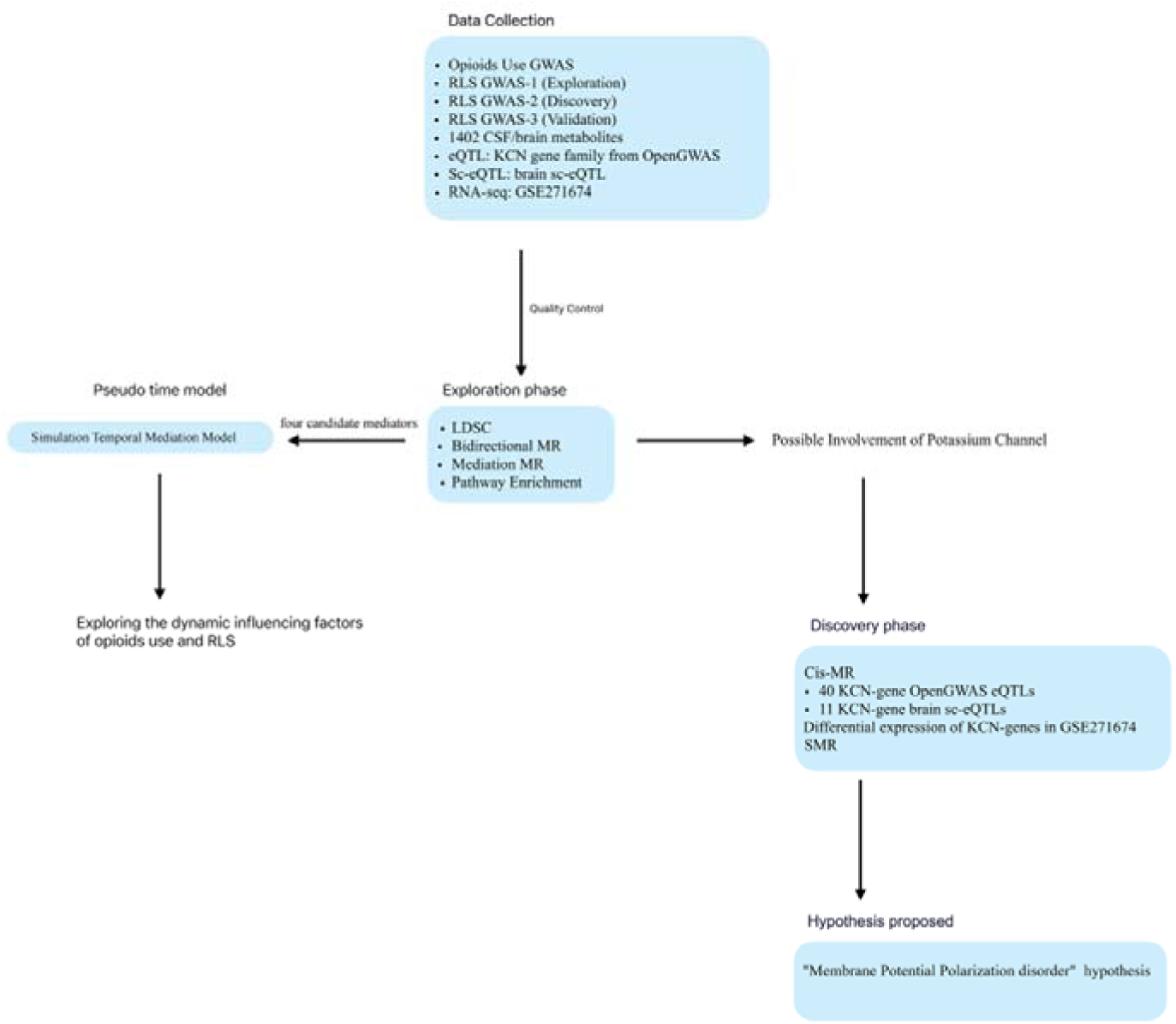
Overall study design. High-resolution versions of all figures can be found in Supplementary Materials.

## Methods

### Data Sources

- **Opioids Use GWAS:** Summary dataset for self-reported prescription opioids use (22,982 cases and 55,826 controls; N = 78,808) from a UK Biobank study^18^.
- **RLS GWAS dataset 1:** Meta-analysis of European-ancestry GWAS (9,851 cases and 38,957 controls; N = 48,808) was obtained from the GWAS Catalog (https://www.ebi.ac.uk/gwas/home, GCST90435387), and derived from a large-scale genome-wide association meta-analysis^19^.
- **RLS GWAS dataset 2:** The second RLS GWAS (7,248 cases and 19,802 controls; N = 27,050) was obtained from the GWAS Catalog (GCST90399568), and derived from the EU-RLS-GENE consortium genome-wide association study^20^.
- **RLS GWAS dataset 3:** The third RLS GWAS (4,599 cases and 495,749 controls; N = 500,348) from the Finngen database (https://www.finngen.fi/en)^21^.
- **CSF and brain metabolite GWAS:** 1402 summary datasets for 440 CSF metabolites and 962 brain tissue metabolites were obtained from the study by Cruchaga et al^22^. (CSF, NiJ=iJ2602; brain tissue, NiJ=iJ1,016; the details can be found in **Supplementary S1-Table 1**).
- **Potassium channel gene eQTLs:** eQTL data for 50 *KCN*-genes datasets was obtained from the OpenGWAS website (https://gwas.mrcieu.ac.uk) which has 19942 eQTLs.
- **Brain sc-eQTL datasets:** The significant brain sc-eQTLs download from scQTLbase^23^ (http://bioinfo.szbl.ac.cn/scQTLbase), and derived from single-nucleus RNA sequencing (snRNA-seq) of 196 adult human brain samples from study by Bryois et al.^24^, profiling eight major CNS cell types (excitatory neurons, inhibitory neurons, oligodendrocytes, OPCs/COPs, astrocytes, microglia, endothelial cells,and pericytes).
- **GSE271674 dataset from Gene Expression Omnibus (GEO) database:** This dataset investigated *MEIS1* transcription factor function in human neural stem cells^25^, comprising 36 samples across six experimental conditions: *MEIS1* overexpression (CRISPRa_MEIS1), *MEIS1* knockout (CRISPR-KO_MEIS1), and their respective controls, with 6 biological replicates per condition (N=36). *MEIS1* harbors the strongest genetic risk factor for RLS and represents a key regulatory node in RLS pathophysiology, making this dataset particularly relevant for validating our findings regarding *KCN*-gene regulation in RLS.

### Genetic Instrument Selection

**Instrument Variable Quality Control Standards:** Stringent quality control criteria were implemented for all genetic instruments:

- Significance thresholds: P<5×10CC for opioids use, RLS1, RLS2 and OpenGWAS eQTL datasets; P<1×10CC for CSF and brain metabolite datasets due to smaller sample sizes; FDR<0.05 for brain sc-eQTL datasets
- Minor allele frequency (MAF): >0.01 to ensure adequate statistical power
- F-statistics: >10 to ensure strong instrument strength, calculated using an improved formula: R² = [2×β²×EAF×(1-EAF)] / [2×β²×EAF×(1-EAF) + 2×SE²×N×EAF×(1-EAF)], then F = [R²×(N-k-1)] / [(1-R²)×k], where β is the effect size, SE is the standard error, EAF is the effect allele frequency, N is the sample size, and k=1 for single SNP evaluation
- Linkage disequilibrium (LD) clumping: r²<0.001, window=10,000kb using European reference panels (GRCh38) for opioids use, RLS1, RLS2 and CSF and brain metabolite datasets; r²<0.3, window=100kb using European reference panels (GRCh38) for genetic region within 100 kb pairs on either side of the target genes from OpenGWAS eQTL datasets and brain sc-eQTL datasets
- Palindromic SNP handling: Removal of A/T and G/C SNPs with ambiguous strand orientation and eaf close to 0.5 (0.45-0.55).

For opioids use, one SNP (rs2618039) reached genome-wide significance, mapping to the *KCND3* gene and encoding a voltage-gated potassium channel (F-statistics=30.6).

For RLS dataset 1, seven independent genetic variants reaching genome-wide significance were selected, after excluding rs7779977 due to outlier behavior and mapping to pseudogenes. All F-statistics for RLS instruments>10.

For CSF and brain metabolites, 1401 datasets containing instrumental variables were obtained by filtering a relaxed threshold (P<1×10CC) yielded sufficient instruments for each metabolite (all F>10). Additional filtering removed SNPs associated with 280 potential confounders (**Supplementary S1-Table 2**).

For *KCN*-gene eQTLs, 40 datasets containing cis-eQTL instrumental variables within 100 kb pairs from the gene were extracted from 50 OpenGWAS KCN-gene eQTLs for cis-MR.

For brain sc-eQTLs, 11 datasets containing instrumental variables within 100 kb pairs of the matched significant *KCN*-genes from the previous cis-MR results were obtained from different brain cell types. The details of these instrumental variables (harmonised_data.csv), and sensitivity analysis and pleiotropy testing results can be found in the corresponding folder in Supplementary Files.

### Statistical Analyses

#### Linkage disequilibrium score correlation (LDSC)^26^

LDSC was used to evaluate the heritability and genetic correlation of opioids use and RLS datasets.

#### Mendelian randomization analysis

The exploratory MR analysis was performed using inverse variance weighted (IVW) method as primary analysis (Wald ratio method for single instrumental variable), supplemented by MR-Egger regression, weighted median method, and MR-PRESSO (NbDistribution = 5000) for pleiotropy detection and outlier correction^27^. Comprehensive sensitivity analyses included Cochran’s Q statistic for heterogeneity testing^28^, MR-Egger intercept test for horizontal pleiotropy detection^29^, and leave-one-out analysis for influential variant identification.

#### Mediation analysis

This exploratory analysis employed a two-stage approach: Stage 1 estimated effects of opioid use on each metabolite. Stage 2 estimated effects of each metabolite on RLS risk. The indirect effect for each mediator was calculated as the product of Stage 1 and Stage 2 effects, and mediation proportion was defined as (indirect effect / total effect) × 100%. Negative mediation proportions indicate masking effects, where risk pathways counteract the direct protective effect of opioids.

#### Simulated temporal mediation model

This exploratory model aims to explore the underlying mechanism by mimicking the dynamic relationship between opioids use and RLS. This model incorporates the following key assumptions and components:

##### Model Assumptions

1. Temporal heterogeneity: Opioid effects on RLS vary across different time periods
2. Multiple pathways: The protective and risk-promoting pathways may play a role at different stages.
3. Masking effects: Long-term risk-promoting mediators counteract and mask the constant direct protective effects.

##### Model Components

1. Direct effect (τ): Constant protective effect, independent of time
2. Short-term protective mediation: immediate neuroprotective mechanisms
3. Long-term risk mediation: neuroadaptive changes
4. Net effect calculation: Total effect = Direct effect + Short-term mediation effect + Long-term mediation effect

##### Mathematical Framework

Total Effect = τ + Σ(a-short-term × b-short-term) + Σ(a-long-term × b-long-term) Where a represents exposure-mediator effects and b represents mediator-outcome effects.

#### Colocalization analysis

To identify shared causal variants between CSF/brain metabolites and RLS dataset 1, Bayesian genetic colocalization analysis^30^ was performed using the coloc R package. For each candidate CSF/brain metabolite identified in the mediation analysis, genomic regions of ±500kb around each instrumental variable SNP were defined. Within these regions, overlapping SNPs from both the metabolite GWAS and RLS GWAS datasets were extracted, requiring a minimum of 10 shared SNPs for analysis. The colocalization analysis calculated five posterior probabilities: PP.H0 (no association), PP.H1 (association with metabolite only), PP.H2 (association with RLS only), PP.H3 (independent associations), and PP.H4 (shared causal variant). PP.H4 > 0.75 was considered as strong evidence for genetic colocalization, indicating that the same causal variant influences both the metabolite level and RLS risk. This analysis helps distinguish between true shared genetic architecture and spurious associations due to linkage disequilibrium.

#### Enrichment analysis gene selection

For functional enrichment analysis, genes were extracted from genomic regions surrounding instrumental variables of candidate CSF and brain metabolites from the mediation analysis. Specifically, genes were derived from ±50kb flanking regions of SNPs serving as instrumental variables for candidate CSF and brain metabolites that showed significant associations (IVW P<0.05) with RLS in the MR analysis. This approach ensures that the enrichment analysis captures genes potentially influenced by the same regulatory mechanisms underlying the metabolite-RLS associations, providing biological context for the observed causal relationships.

A systematic four-step gene filtering strategy was implemented to ensure high-quality gene selection: (1) Duplicate removal: redundant gene terms were eliminated to avoid bias in enrichment analysis; (2) Functional annotation filtering: genes were queried through the GeneCards database (https://www.genecards.org/) to filter genes based on comprehensive functional annotations and disease relevance; (3) SNP-gene correspondence optimization: genes were prioritized to achieve one-to-one correspondence with SNPs where possible, reducing mapping ambiguity; (4) Protein-coding gene restriction: only protein-coding genes were retained to focus on functionally relevant targets with direct therapeutic implications. Through this rigorous filtering process, the final gene set was refined from an initial 2,879 SNP-mapped genes to 486 unique high-quality genes. Gene Ontology (GO) and Kyoto Encyclopedia of Genes and Genomes (KEGG) pathway enrichment analyses were performed to find significantly enriched biological processes, cellular components, molecular functions, and metabolic pathways (p.adjust<0.05).

To validate these findings, a replication analysis for the key steps was conducted using the RLS GWAS dataset 2. The MR analysis was conducted using the same CSF and brain metabolites as exposures with the RLS dataset 2 as an outcome. The functional enrichment analysis was conducted on genes associated with significantly causal metabolites in the validation dataset to assess biological pathway consistency across different RLS populations.

#### Cis-MR

Cis-MR based on OpenGWAS single gene eQTLs, was used to evaluate the link between the potassium channels and the RLS datasets. Subsequently, cis-MR based on sc-eQTL from 8 types of brain cells, was used to validate the involvement of the significant genes.

#### Differential expression analysis in GSE271674 datasets

sample information was extracted from the series matrix file to ensure accurate group assignment and proper sample-to-condition mapping. Differential Expression Analysis: DESeq2 analysis was performed separately for *MEIS1* overexpression versus control and *MEIS1* knockout versus control comparisons. For each comparison, count matrices were filtered to remove low-expression genes (genes with total counts < 10 across all samples), and DESeq2 normalization was applied. Differential expression analysis employed the Wald test with FDR correction for multiple testing (Benjamini-Hochberg method). The results included log2 fold changes, raw p-values, and FDR-adjusted p-values for each gene. Corrected Log2 Fold Change Calculation: To assess the net regulatory effect of MEIS1 on target genes, corrected log2 fold changes were calculated by subtracting the knockout log2 fold change from the overexpression log2 fold change (corrected_log2FC = OE_log2FC - KO_log2FC). Statistical significance of corrected log2 fold changes was evaluated using Fisher’s method to combine p-values from both comparisons, followed by FDR correction. This approach accounts for the bidirectional nature of *MEIS1* manipulation and provides a comprehensive assessment of *MEIS1’s* regulatory impact on target genes. Target Gene Expression Analysis: Potassium channel gene family members were specifically extracted from the differential expression results. For *KCN*-genes, raw expression data were retrieved from the original count matrix, integrated with sample grouping information, and analyzed for expression patterns across experimental conditions.

#### Summary-data-based MR analysis

This is a method (called SMR) that integrates summary-level data from GWAS with data from expression quantitative trait locus (eQTL) studies to identify genes whose expression levels are associated with a complex trait because of pleiotropy^31^. In this study, I used the lite version of the GTEx V8 cis-eQTL summary data (only with SNPs < 1e-5 are included) across 49 human tissues (**Supplementary S1-Table 20**) from the GTEx Consortium (https://gtexportal.org/home/datasets) that download from Yang Lab (https://yanglab.westlake.edu.cn). To avoid omissions, the SMR analysis using cis-eQTL data across all 49 human tissues, then filter to obtain the significant results (SMR pval < 0.05, HEIDI pval > 0.05) related to *KCN*-gene.

## Results

### Genetic Correlation

LDSC analysis showed: (1) the SNP-based heritability of RLS dataset 1 was estimated at h^2^ = 0.1097 (SE = 0.0194). The mean χ2statistic was 1.0843, and the intercept was 0.9933 (SE = 0.0094), indicating minimal confounding from population structure or technical artifacts after genomic control (GC) correction; (2) RLS dataset 2 showed higher heritability: h^2^=0.2073 (SE=0.0442). The mean χ2was 1.1305, with an intercept of 1.0323 (SE = 0.0105), suggesting a robust genetic signal; (3) the genetic correlation between the RLS dataset 1 and 2 was low and non-significant: rg = 0.0122, SE = 0.0572, Z = 0.2132, P = 0.8312, indicating genetic heterogeneity; (4) the genetic correlation between opioids use and RLS dataset 1 showed significant positive: rg=0.0983, SE=0.0467, Z=2.1040, P=0.0354. To further understand the difference between the RLS datasets, Manhattan and QQ plots were plotted (**Fig. 2**).

**Fig. 2.**
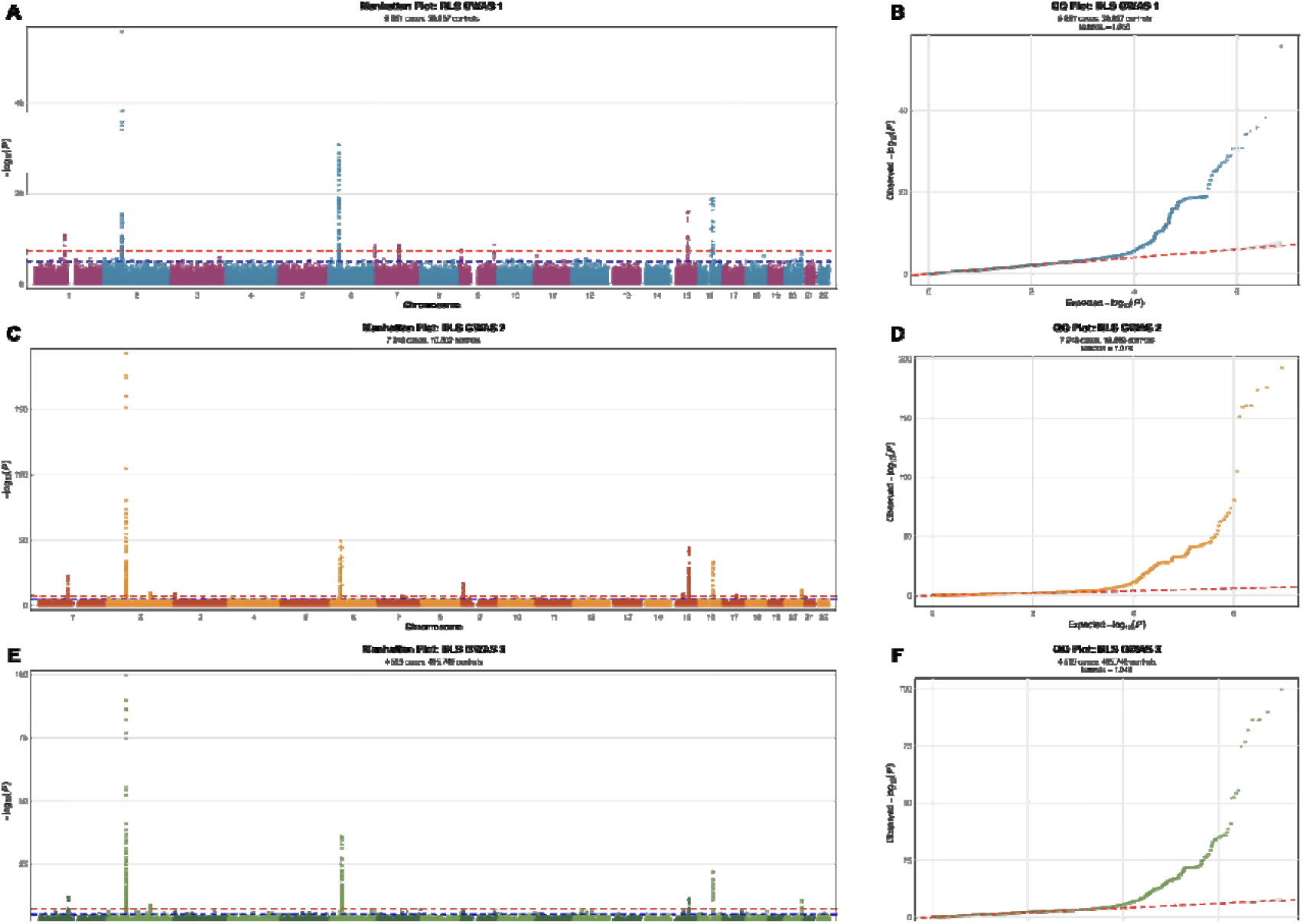
Manhattan Plots and QQ Plots of the RLS datasets.

RLS dataset 1 as an outcome was used to explore the dynamic relationship between opioids use and RLS because of the genetic correlation. RLS dataset 2 and 3 were used to explore the relationship between potassium channels and RLS due to more significant SNPs and stronger genetic signals.

### MR analysis

The exploratory MR analysis using rs2618039 after screening as the only instrumental variable with the Wald ratio method showed a potential negative correlation of opioids use on RLS risk (RLS dataset 1 as outcome: OR=0.613, 95% CI: 0.359-1.046, P=0.072), but not reaching conventional statistical significance. Reverse MR analysis (RLS dataset 1 as exposure and opioids use as outcome) showed a significant positive correlation (OR=1.066, 95% CI: 1.011-1.124, P=0.018). Further details of reverse MR analysis can be found in **Supplementary S1-Table 3 and File 1**.

Although there is ample evidence from clinical observations, further investigation is needed to validate the relationship in genetics between opioids use and RLS. Therefore, a simulated temporal mediation model was proposed to explore the potential dynamic relationship between them.

### Mediation Analysis

#### Stage 1 (Opioids Use → CSF and Brain Metabolites)

Using the Wald ratio method (single instrumental variable analysis), 51 correlations between opioids use and CSF and brain metabolites (P<0.05) were retained as candidates after excluding 2 reverse causality relationships and 2 drug concentration-related exposures (**Fig. 3 A and D**, the details see **Supplementary S1-Table 4**).

**Fig. 3.**
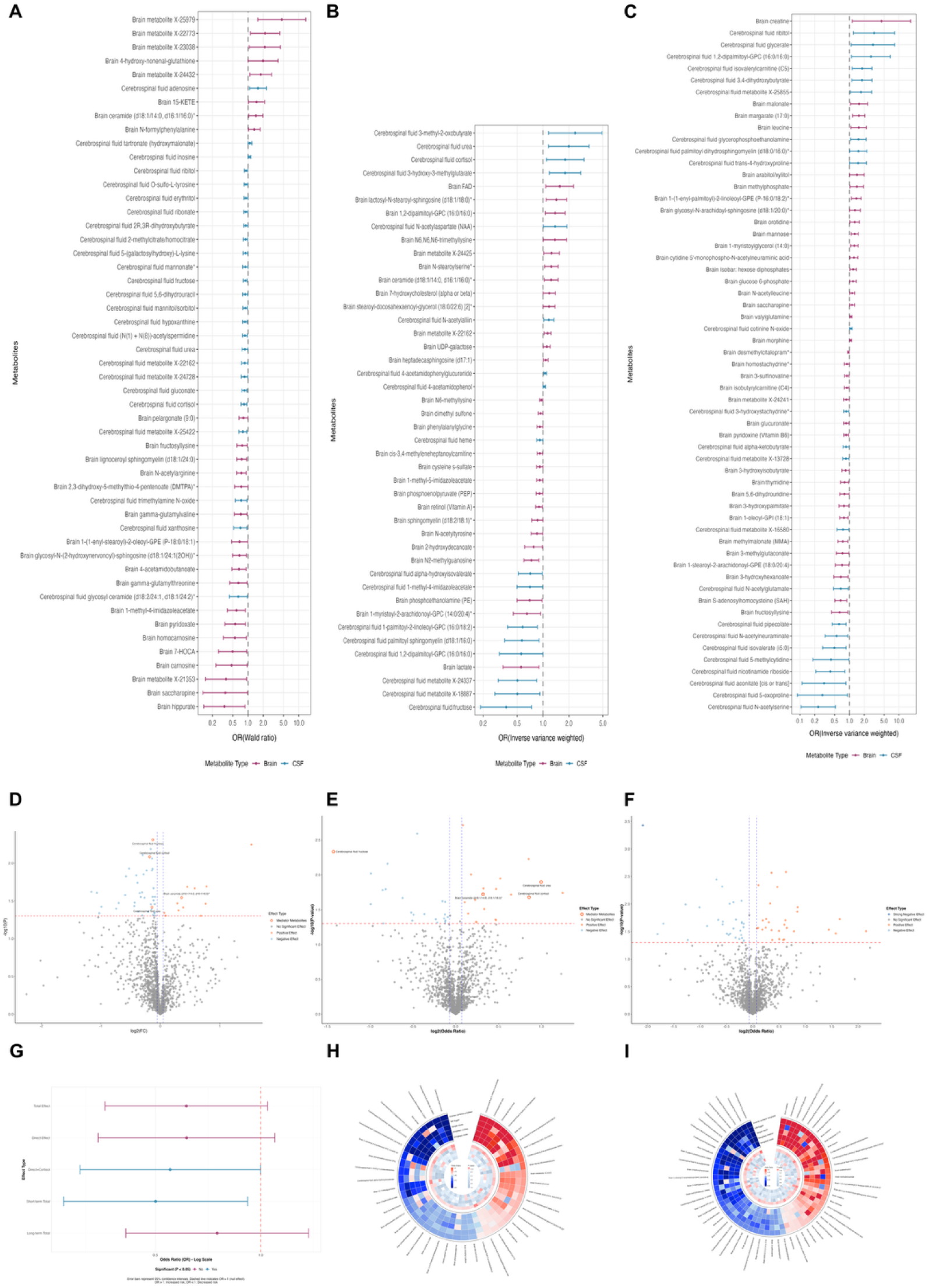
Mediation analysis and simulated temporal mediation model. Forest plots and volcano plots showing correlations of opioids use on 51 candidate CSF and brain metabolites (**A, D**), correlations of 44 candidate CSF and brain metabolites on RLS dataset 1 (**B, E**), and correlations of 59 candidate CSF and brain metabolites on RLS dataset 2 (**C, F**). (**G**) By removing assuming long-term risk mediators, the short-term total efficacy of opioids use becomes significant. Circular heatmaps showing multiple methods results of MR analyses between candidate CSF and brain metabolites and RLS datasets. (**H:** RLS dataset 1, **I:** RLS dataset 2).

#### Stage 2 (CSF and Brain Metabolites → RLS dataset1)

The exploratory MR analysis showed 44 correlations between CSF/brain metabolites and RLS dataset 1 using the IVW method (P<0.05) without reverse causality relationship, which as candidates were included in subsequent mediation analysis (**Fig. 3 B, E and H**; the details see **Supplementary S1-Table 5 and File 2**). To preserve more genetic signals in exploration phase^32^, multiple testing correction was not performed on the candidate MR results in the stage 1 and 2.

#### Candidate Mediators and Temporal Effects

Bootstrap mediation analysis (10,000 iterations, **Supplementary S1-Table 11**) evaluated four candidate CSF/brain mediators and the direct effect: CSF cortisol (GCST90318234, a-effect β=-0.181, P=0.008; b-effect β=0.594, P=0.021; indirect effect=-0.108, mediation proportion=21.96%), urea (GCST90318260, a-effect β=-0.139, P=0.038; b-effect β=0.692, P=0.013; indirect effect=-0.096, mediation proportion=19.60%), fructose (GCST90318283, a-effect β=-0.124, P=0.005; b-effect β=-0.990, P=0.005; indirect effect=0.123, mediation proportion=-25.07%), and brain ceramides (GCST90318755, a-effect β=0.358, P=0.029; b-effect β=0.221, P=0.019; indirect effect=0.079, mediation proportion=-16.12%); the direct effect=-0.488.

The simulation idealized temporal mediation model simulated a situation similar to clinical observations that short-term opioids use was significant negative correlation (OR=0.501, 95% CI: 0.273-0.918, P=0.025), however, the correlation may decrease (OR=0.751, 95% CI: 0.411-1.371, P=0.351) when the risk mediation effect (fructose depletion and ceramide accumulation) was activated and the protective mediation (cortisol and urea suppression) effect was ineffective (**Fig. 3 G**).

### Colocalization Analysis

Based on the colocalization analysis of candidate CSF and brain metabolites and RLS dataset 1, a strong colocalization evidence was identified for SNP (rs11045279) between CSF 4-acetamidophenylglucuronide levels (GCST90317906) and RLS dataset 1, which maps to the *PDE3A* gene region. The posterior probability of colocalization (PP.H4) reached 87.2%, exceeding the conventional threshold for robust genetic colocalization. Parameter sensitivity analysis across different prior combinations (p1/p2=1e-4 to 1e-6, p12=1e-5 to 1e-7) showed robust colocalization with H4 values consistently above 0.8 under mainstream recommended priors, supporting the stability of this finding.

### Enrichment Analysis

To further explore the pathogenesis of RLS, GO and KEGG pathway enrichment analyses were conducted using 486 genes filtered from 2879 genes (**Supplementary S1-Table 6 and 7**) that were obtained from instrumental variable SNPs of 44 candidate MR results between CSF/brain metabolites and RLS dataset 1.

The GO enrichment analysis from 486 input genes showed, 45 biological processes (BP), 23 cellular components (CC), and 16 molecular functions (MF) reached statistical significance (p.adjust<0.05). The analysis showed multiple potassium channel-related pathways were significantly enriched, including potassium channel complex (GO:0034705, CC, 14 genes, p.adjust=3.77e-05, fold enrichment=5.92), voltage-gated potassium channel complex (GO:0008076, CC, 13 genes, p.adjust=3.77e-05, fold enrichment=6.17) and potassium ion transmembrane transport (GO:0071805, BP, 18 genes, p.adjust=0.0032, fold enrichment=3.51), indicating that potassium ion channels may play a important role. Synaptic function-related pathways were prominently represented, with postsynaptic density membrane (GO:0098839, CC, 12 genes, p.adjust=0.00254, fold enrichment=4.03), synapse assembly (GO:0007416, BP, 20 genes, p.adjust=0.00387, fold enrichment=3.09) and positive regulation of axonogenesis (GO:0050772, BP, 10 genes, p.adjust=0.00387, fold enrichment=5.7), supporting the involvement of synapse in RLS pathophysiology.

8 KEGG pathways reached statistical significance (p.adjust<0.05), with morphine addiction pathway showing the strongest enrichment (11 genes, p.adjust=0.00335, fold enrichment=4.98). Other significantly enriched pathways included axon guidance, GABAergic synapse, serotonergic synapse, and cholinergic synapse. GO and KEGG complete results related to RLS dataset 1 can be found in **Supplementary S2**, and the top 20 results also can be seen in **Figure 4 A-D**.

**Fig. 4.**
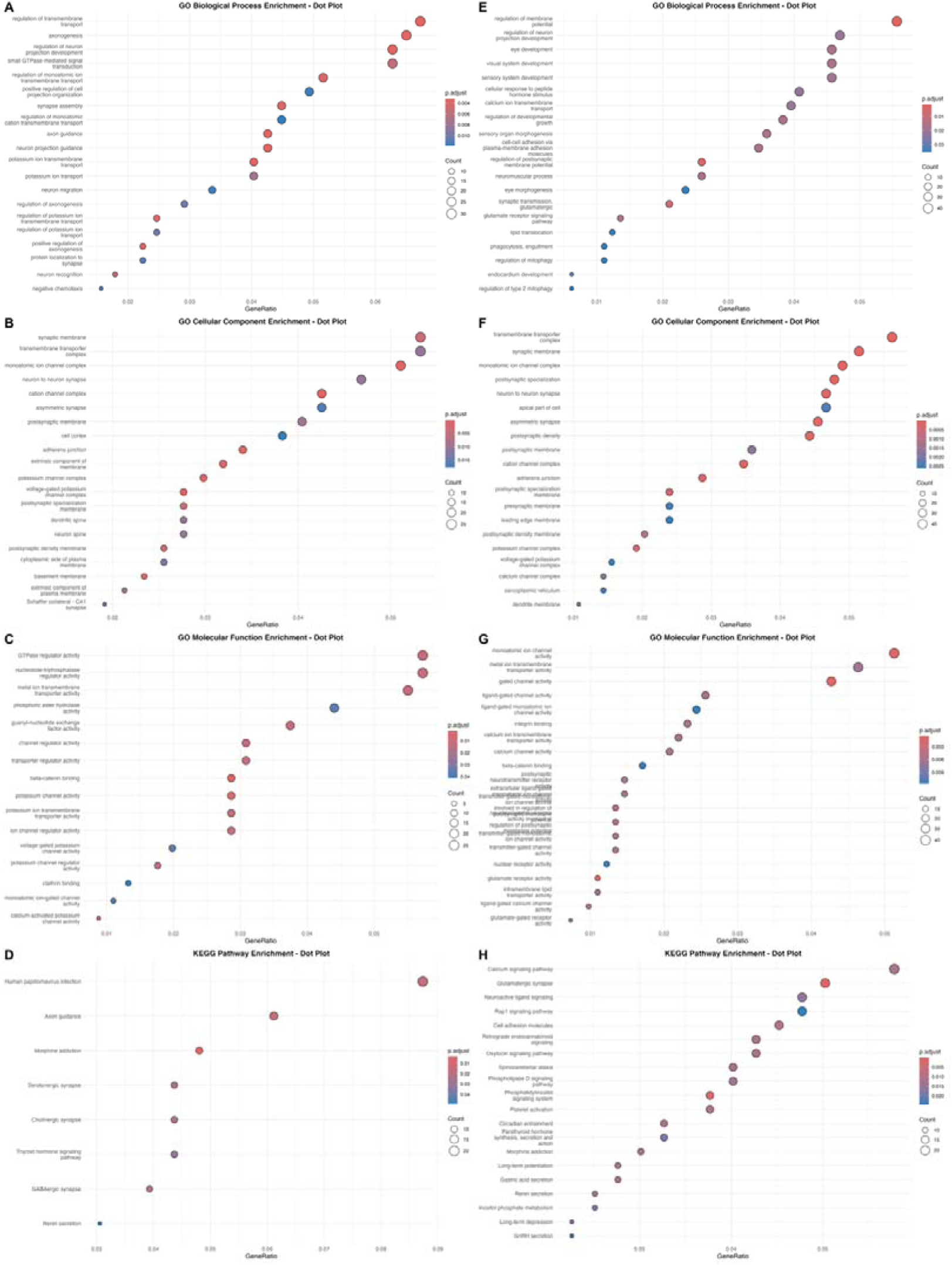
The top 20 results of GO and KEGG enrichment analysis from different RLS datasets.

#### Replication Enrichment Analysis

I conducted MR analysis using the same CSF/brain metabolites and the RLS dataset 2, which showed 59 candidate metabolites with potential correlations (P<0.05, **Fig. 3 C, F and I**; the details see **Supplementary S1-Table 8 and File 3**). Finally, 871 genes were obtained from 5384 genes for enrichment analysis after screening (**Supplementary S1-Table 9 and 10**), and 131 GO enrichment terms and 28 KEGG pathways with statistical significance (p.adjust<0.05) were identified as candidates (**Fig. 4 E-H**; the complete enrichment results can be found in **Supplementary S3**), then compared with the enrichment analysis results of the RLS dataset 1, by the intersection of the two significance results (all p.adjust<0.05). Pearson product-moment correlation analysis results based on fold enrichment at different thresholds between RLS dataset 1 and 2 enrichment results showed consistency in synaptic function, potassium channel and membrane potential regulation, and cell connectivity (**Fig. 5**), supporting robust biological pathway convergence. The detailed data comparison can be seen in **Supplementary S4**.

**Fig. 5.**
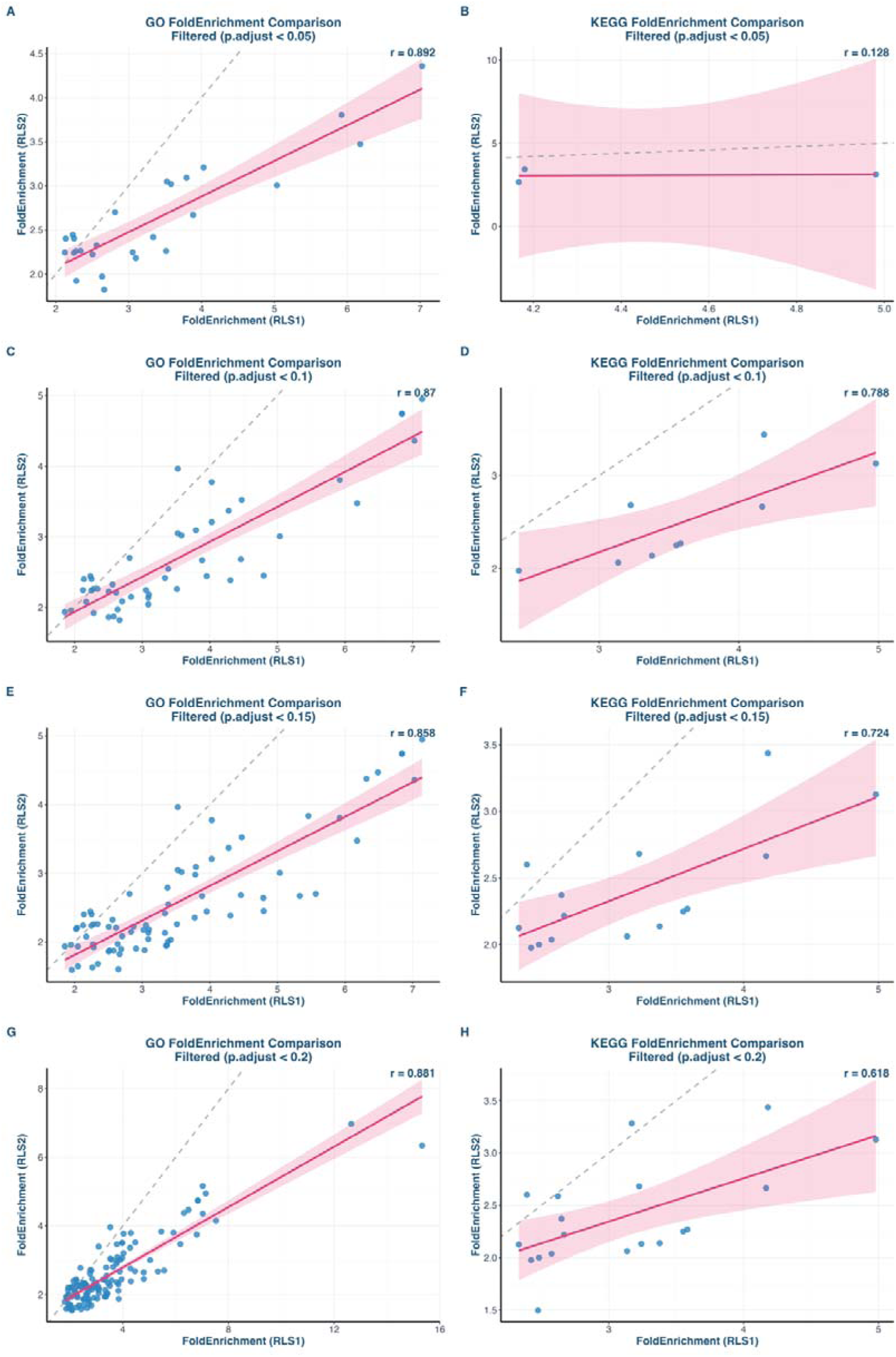
Fold enrichment correlation analysis at different thresholds.

### Cis-MR Analysis

After screening, 40 instrumental variable datasets were obtained from the OpenGWAS eQTLs of 50 *KCN*-genes for single gene cis-MR analysis to evaluate the impact of potassium channels related genes on RLS. For RLS dataset 1, the cis-MR analysis significant result showed that 3 candidate genes (*KCNAB1*, *KCNAB2* and *KCNIP2*) with potential roles (P<0.05), of which 2 genes (*KCNAB2* and *KCNIP2*) passed multiple testing correction (**Fig. 6 A**). For RLS dataset 2, the result showed that 15 candidate genes with potential roles (P<0.05), of which 11 genes (*KCNA6*, *KCNAB1*, *KCNAB2*, *KCNB1*, *KCNH2*, *KCNJ15*, *KCNK13*, *KCNK6*, *KCNMA1*, *KCNN4* and *KCNQ5*) passed multiple testing correction (**Fig. 6 C and Fig. 7**). For RLS dataset 3, the result showed 11 candidate genes with potential roles (P<0.05), of which 2 genes (*KCNK13* and *KCNT1*) passed multiple testing correction (**Fig. 6 E**). The details can be found in **Supplementary S1-Table 12-14 and Files 4-6**.

**Fig. 6.**
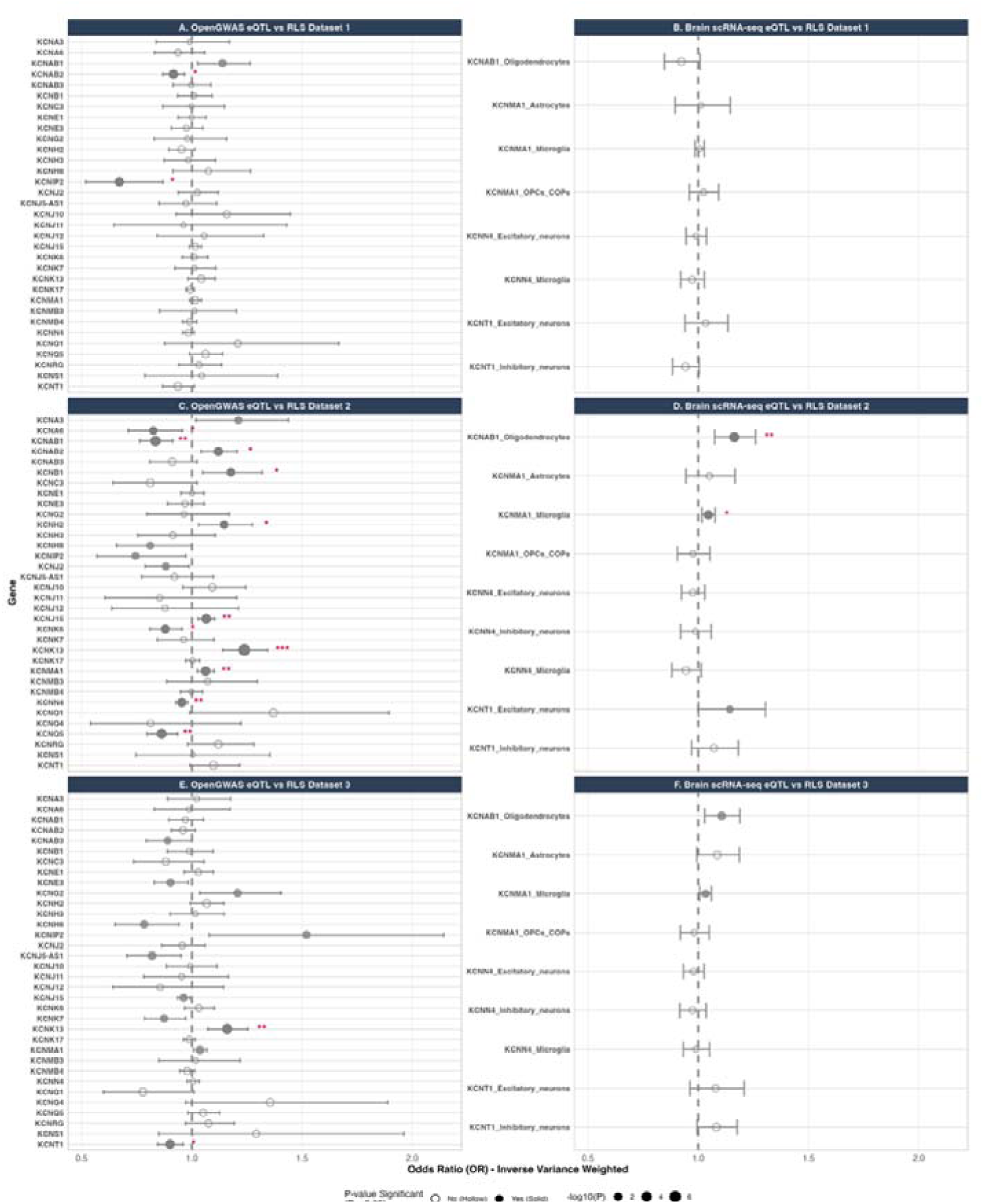
The results of cis-MR analyses between *KCN*-genes and RLS across different RLS datasets.

**Fig. 7.**
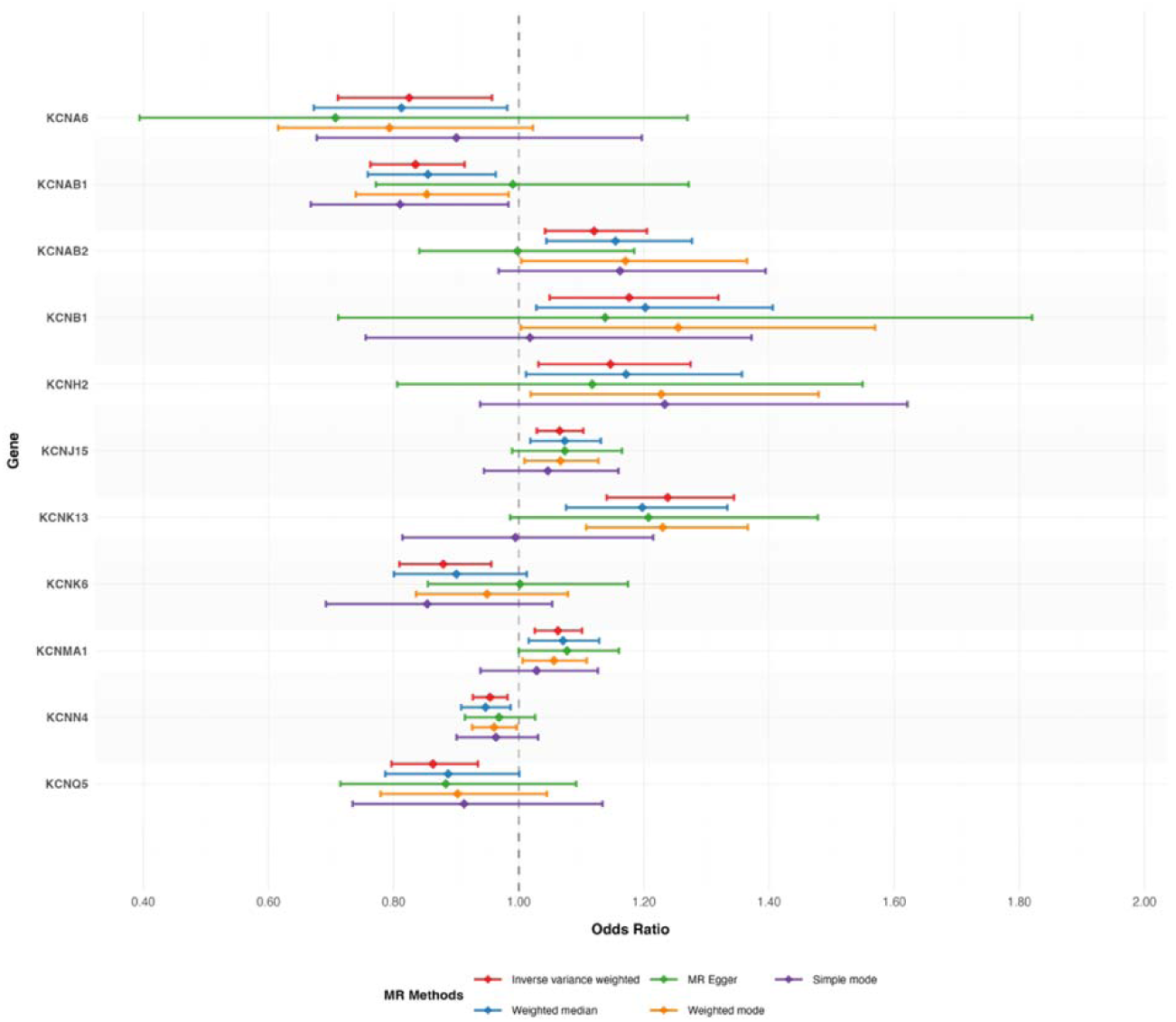
Cis-MR significant results between OpenGWAS *KCN*-gene eQTLs and RLS dataset 2.

Subsequently, based on the 13 significant genes from the previous cis-MR, 11 sc-eQTLs were matched from 8 types of brain cells, including: *KCNMA1* from astrocytes; *KCNK6*, *KCNK13*, *KCNN4* and *KCNT1* from excitatory neurons; *KCNN4* and *KCNT1* from inhibitory neurons; *KCNMA1* and *KCNN4* from microglia; *KCNAB1* from oligodendrocytes; *KCNMA1* from oligodendrocyte precursor cell (OPC). The cis-MR result between these sc-eQTLs and RLS dataset 1 showed no significant genes (**Fig. 6 B**). For RLS dataset 2, the results showed positive correlations of *KCNAB1* from oligodendrocytes (OR=1.16, 95% CI: 1.07-1.26, P=2.17e-04, FDR P=0.002), *KCNMA1* from microglia (OR=1.05, 95% CI: 1.02-1.08, P=0.002, FDR P=0.012), *KCNK6* (OR=1.20, 95% CI: 1.002-1.44, P=0.047, FDR P=0.133) and *KCNT1* (OR=1.143, 95% CI: 1.001-1.305, P=0.048, FDR P=0.133) from excitatory neurons (**Fig. 6 D**). For RLS dataset 3, the results also showed potential positive correlations of *KCNAB1* from oligodendrocytes (OR=1.11, 95% CI: 1.03-1.19, P=0.006, FDR P=0.071), *KCNMA1* from microglia (OR=1.03, 95% CI: 1.01-1.06, P=0.014, FDR P=0.075) (**Fig. 6 F**). These findings further enhance the robustness of the conclusion that potassium channels are involved (The details see **Supplementary S1-Table 15-17 and Files 7-9)**.

### Differential expression analysis of *KCN*-genes in GSE271674 datasets

Through differential expression analysis of genes in the GSE271674 dataset and extraction of genes of interest, the analysis result showed that 1841 significantly differentially expressed genes (**Supplementary S1-Table 18**) were filtered through corrected log2FC (padjust<0.05) from 17495 genes, including 8 *KCN*-genes: *KCNC1*, *KCNH1*, *KCNH2*, *KCNH3*, *KCNJ1*, *KCNJ2*, *KCNJ6*, *KCNMB4* (**Fig. 8**). For example, *MEIS1* activation may result in significant positive expression of *KCNH3* (log2FC=2.20, P=2.04e-16), while *MEIS1* knockout showed no significant change (log2FC=0.48, P=0.487). The corrected log2 fold change, accounting for bidirectional *MEIS1* manipulation, was 1.72 (FDR-corrected P=4.39e-13), indicating *MEIS1*-activation-mediated significant positive expression of *KCNH3* (**Fig. 8 D**). For *KCNMB4* gene, the *MEIS1* activation was a negatively correlation (**Fig. 8 H**). The details can be found in **Supplementary S1-Table 19**. These findings contribute to understanding the potential regulatory patterns of *MEIS1* on *KCN*-genes.

**Fig. 8.**
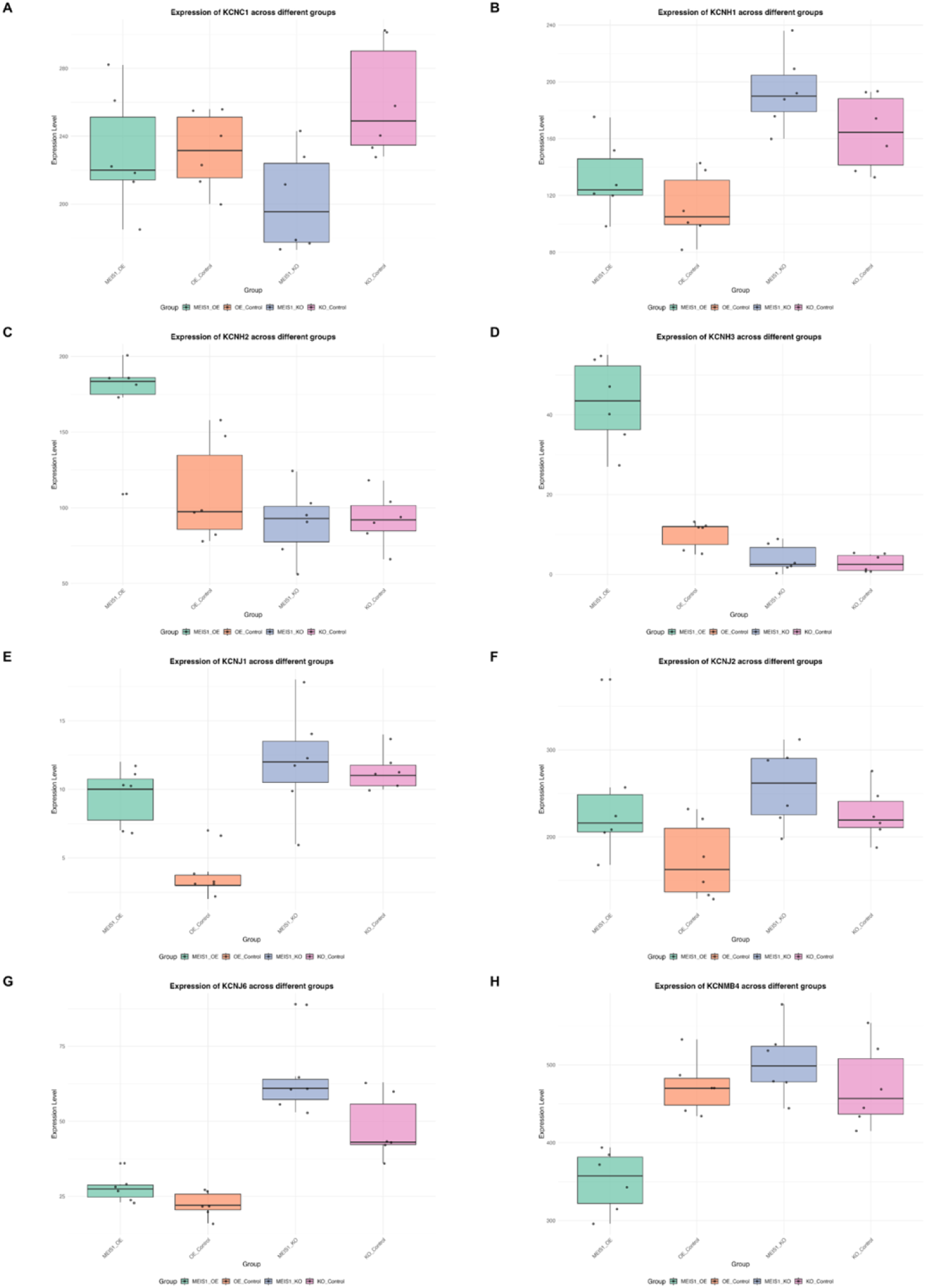
K*C*N-genes expression boxplot across different groups in human neural stem cells from GSE271674 dataset.

### Summary-data-based MR analysis

For RLS dataset 1, the SMR results showed 599 significant results were obtained from 195,137 results, and the significant *KCN*-gene was *KCNT2* in esophagus gastroesophageal junction. For RLS dataset 2, the SMR results showed 11,668 significant results were obtained from 216,156 results, and the multiple significant *KCN*-genes in different tissues were found, such as *KCNAB2*, *KCNH2*, *KCNH7* and *KCNK13*. For RLS dataset 3, the SMR results showed 10,175 significant results were obtained from 225,480 results, and the multiple significant *KCN*-genes in different tissues were found, such as *KCNJ14*, *KCNAB2* and *KCNK10*. The details can be found in **Supplementary S1-Table 21-23**.

## Discussion

In the simulated temporal mediation model, the four candidate mediators (CSF cortisol, urea, fructose, and brain ceramides) were found from mediation analysis, then assigned idealized time attributes, provide direction for subsequent exploring the temporal dynamics of opioids–RLS interactions through simulating a situation similar to clinical observations. Assuming time-dependent mediating factors either enhance or counteract the protection effect at specific stages:

Short-term collaborative enhanced protective mechanism: assuming the risk mediators did not play roles, their effects were removed. (Direct effect + Short-term mediation effect = Total effect - Long-term mediation effect)

- Hypothalamus-pituitary-adrenal (HPA) axis suppression: Opioids can reduce CSF cortisol levels^33,34^, thereby decreasing synaptic excitability^35,36^ and may provide protective effects by avoiding excessive activation of the HPA system^37^.
- Blood-brain barrier modulation: Although the precise mechanism requires further investigation, decreased CSF urea levels may suggest altered blood-brain barrier permeability. An animal study suggested that the blood-brain barrier is involved in the formation of blood and CSF urea concentration gradients^38^. Long-term masking protective mechanism: assuming the protective mediators did not play roles, their effects were removed. (Direct effect + Long-term mediation effect = Total effect - Short-term mediation effect)
- Metabolic dysregulation: Reduced CSF fructose levels may impair dopamine-mediated reward mechanisms and exacerbate RLS symptoms, as fructose may stimulate dopamine secretion^39^ and effect reward mechanisms^40^.
- Tolerance development: Long-term opioids use can lead to the accumulation of the ceramides levels and the development of tolerance^41^. Ceramides play a crucial role in normal and neurological function and disease processes by regulating synaptic structure, neurotransmitter transmission, and plasticity^42^.

In the exploratory MR analysis between opioids use and RLS dataset 1, The identification of rs2618039 in the *KCND3* gene as a strong instrument (F=30.6) for opioids use suggests a genetic clue linking voltage-gated potassium channels to RLS pathophysiology. Searching on the GWAS Catalog website, it was found that rs2618039 is also correlated with body mass index and education attainment (**Supplementary S5**), further searching of the *KCND3* gene showed its association with multisite chronic pain and cardiac electrophysiology (**Supplementary S5**), suggesting the influence of *KCND3* in electrophysiology, which was consistent with previous studies on spinocerebellar ataxia subtypes SCA19/22^43^ and cardiac repolarization^44,45^. For RLS GWAS dataset 1, the functional enrichment analysis of 486 genes derived from 44 candidate metabolites suggests an important clue: the involvement of potassium channels and membrane potential regulation. The replication enrichment analysis with RLS dataset 2 provides robust support for this finding. Despite the low genetic correlation (rg = 0.0122, P = 0.8312) between the two RLS datasets, indicating genetic heterogeneity, the biological pathway analysis showed a significant similarity, indicating a potential common core mechanism across RLS populations——regardless of upstream causes, the mechanism unifies RLS phenotypes.

Potassium channels locate in cell membranes, and based on gene family, structure and functional characteristics, that are grouped into four families: *1*) calcium- and sodium-activated potassium channels (*KCa/Na*) (6/7 TMs); *2*) inwardly rectifying potassium channels (*Kir*) (2 TMs); *3*) two-pore-domain potassium channels (*K2P*) (4 TMs); and *4*) voltage-gated potassium channels (*Kv*) (6 TMs)^5^. Potassium channels play an important role in maintaining resting potential, repolarizing action potential, and regulating cellular excitability^46^. This dysfunction may create a cascade of membrane potential abnormalities: depolarized resting potential, prolonged repolarization time, enhanced neuronal excitability, and amplified synaptic signaling. The resulting membrane potential instability could first to trigger sensory symptoms through aberrant activation of low-threshold sensory channel, due to differences in perceptions.

To further evaluate the correlation between potassium channels and RLS, the cis-MR analysis of 40 bulk eQTL data from OpenGWAS showed 15 candidate genes and 11 significant genes causal-related to RLS dataset 2, including higher expression of *KCNAB1* was associated with a protective effect against RLS. In contrast, single-cell eQTL-based MR analysis indicated that *KCNAB1* expression in oligodendrocytes conferred an increased risk of RLS. For *KCNMA1*, both bulk and microglia-specific single-cell eQTL MR analyses consistently supported that higher expression was associated with an increased risk of RLS. These findings highlight the complexity of potassium channel regulation in RLS. The opposing results for *KCNAB1* suggest that its effect is cell-type dependent: protective associations observed in bulk tissue may be driven by other cell populations, whereas oligodendrocyte-specific signals showed a deleterious role. In contrast, the concordant results for *KCNMA1* across bulk and microglial eQTL data strengthen the evidence that increased *KCNMA1* expression contributes to RLS risk, particularly through microglia-mediated mechanisms. Together, these observations emphasize the importance of incorporating single-cell eQTL data into causal inference frameworks, as they uncover cell-type-specific regulatory mechanisms that are masked in bulk analyses. They also suggest that distinct glial cell populations—oligodendrocytes for *KCNAB1* and microglia for *KCNMA1*—may represent critical cellular contexts for the pathogenic involvement of potassium channels in RLS.

However, it is worth noting that the RLS dataset 2 has stronger heritability and more significant susceptibility genes, suggesting a more concentrated upstream etiology and leading to more significant results in the replication enrichment stage, and the same trend was also observed in cis-MR analysis. The latest large-scale GWAS meta-analysis by Schormair et al.^20^, includes and extends the RLS dataset 2. In this expanded dataset (GCST90432061, 116,647 cases and 1,546,466 controls; N = 1,663,113), genome-wide significant associations were observed for *KCNAB1*, *KCNMA1* and *KCNK13* (**Supplementary S5**). Although not independent of RLS dataset 2, these findings reinforce the MR and enrichment results by demonstrating that potassium channel genes are directly implicated at the population level in the largest available sample, which is not an artefact of weak instruments or small-sample effects.

In addition, the analysis result from the differential expression of GSE271674 indicates that activation of *MEIS1* gene can cause upregulation of *KCNH3* gene expression. Recent studies have shown that *KCNH3* and *KCNH8* play crucial roles in regulating the nighttime repetitive firing rate of the suprachiasmatic nucleus neurons and controlling their circadian rhythm^47^. Therefore, knocking out the *MEIS1* gene may lead to a decrease in the regulatory ability of circadian rhythm mediated by the inability to activate the *KCNH3* gene expression. This dynamic regulatory pattern may lead to an increase in false negative results, as it is impossible to distinguish the differential expression of *KCNH3* gene between *MEIS1* knockout and resting state. Interestingly, an important paralog of this gene is *KCNH8* which also was negative correlation trend with RLS in cis-MR (RLS dataset 2: OR=0.81, 95% CI: 0.659-0.998, P=0.48; RLS dataset 3: OR=0.783, 95% CI: 0.652-0.941, P=0.009), suggesting potential protective effects (**Fig. 6 C and E**).

The analysis results of this study support the complex causal relationship between the regulation of multiple gene subtypes related to potassium channels and RLS. Given the key role of potassium channels in cell membrane potential regulation, a novel potential mechanism, “Membrane Potential Disturbance”, was proposed, which could be a link between diverse upstream etiologies and downstream similar mechanisms/symptoms, that suggests a potential unified explanation for RLS:

### Dopaminergic system changes and addiction

CSF testing, neuroimaging and post-mortem studies in RLS patients consistently support dopaminergic system hyperactivity^3,4^, and the latter two also support secondary dopamine receptor downregulation. Previous studies have shown that changes in excitability and dopamine secretion of dopaminergic neurons can be regulated by different types of potassium ion channels through membrane potential^48,49^. Specifically, dopaminergic neurons, as self-initiated pacemaker cells, are sensitive to the transmembrane “background depolarization drive” in terms of pacing rate^50,51^: moderately pushing the membrane potential towards depolarization will increase the discharge frequency and change the discharge mode^48,49^. Conversely, activated dopaminergic neurons maintain homeostasis by compensatory increase in potassium channel currents^52,53^. *KCNQ* potentiation via retigabine administration can normalize hyperexcitability in ventral tegmental area (VTA) dopaminergic neurons to the nucleus accumbens, resulting in significant alleviation of thermal hypersensitivity in neuropathic pain models^54^.

Increased dopamine also may reduce presynaptic glutamate release through dopamine D2 receptor-mediated inhibition of the indirect pathway, thereby indirectly suppressing synaptic excitability via GABAergic interneuron activity in the striatum^55^. And the D2 receptor mediates the reward circuit from VTA to the central amygdala may involve in pain relief^56^, and use pain relief as a reward to change behavior^57^.

Although the hyperactive dopaminergic system can maintain homeostasis and analgesia through negative feedback and reward circuits, chronic dopamine stimulation may induce D2 receptor downregulation and synaptic plasticity changes, contributing to addiction vulnerability^55^, and may create a sustained state of “dopamine hyperactivity-hypoallergenic”. The probability of developing RLS during opioid withdrawal is higher than during alcohol withdrawal^58,59^. KEGG enrichment analysis showed involvement of morphine addiction mechanism and was replicated in replication analysis, consistent with clinical observation. Different subtypes of potassium ion channels can participate in the addiction mechanisms of alcohol, cocaine, methamphetamine, and opioid drugs by regulating neuronal excitability, dopamine release, and neuroadaptive changes^60^, which may also cause RLS. The *KCNK13* gene is a key target for ethanol induced VTA and is differentially expressed in acute and chronic ethanol exposure and withdrawal reactions, with upregulation at 24 hours and downregulation at 72 hours in withdrawal^61^. In this study, the *KCNK13* eQTL cis-MR results across different RLS datasets supported that upregulation of the gene expression led to an increased risk of RLS. These findings suggest a complex relationship between potassium channels, addiction, and RLS, where addiction and RLS may mutually promote development.

### Iron Deficiency

Iron deficiency plays an important role in the occurrence and development of RLS, and iron supplementation is preferred over other treatment methods in RLS patients with concomitant iron deficiency^2,62^. In animal models, iron deficiency increases acute and chronic pain responses in mice and may cause the increased pain sensitivity^63,64^. However, brain iron deficiency has been proven to occur even if peripheral iron is normal^62^, suggesting the presence of iron transport disorders. Iron concentration in neurons and glial cells recently determined in adult rat brains showed that 0.57 mM was the average intracellular iron concentration in cortical neurons, and that astrocytes and microglia cells presented greater iron levels, with averages of 1.29 mM and 1.76 mM, respectively, however, the iron concentration found in mature oligodendrocytes was 3.05 mM, which are the cell type with the highest iron content in the brain and double the iron content of astrocytes and microglia cells and quintuples the average in neurons^65^. Iron metabolism in oligodendrocytes is closely related to myelination and remyelination^66,67^. A review included 12 studies indicated that reduced oligodendrocyte numbers and decreased myelin thickness may play a crucial role in the development or maintenance of pain^68^. Postmortem and imaging in RLS individuals showed a decrease in myelin similar to that reported in animal models of iron deficiency, with decreased ferritin and transferrin in the myelin fractions^69^. In this study, the cis-MR results based on sc-eQTL support that the upregulation of *KCNAB1* from oligodendrocytes as a risk factor across different RLS datasets, providing genetic support for the involvement of oligodendrocytes in RLS. Moreover, the hypothesis based on abnormal membrane potential suggests a new pathway to link iron deficiency with changes in the dopaminergic system.

### Periodic Limb Movement Disorder and Neural Electrophysiology

At the genetic, epidemiological, electrophysiological, and pharmacological response levels, RLS and periodic limb movement disorder (PLMD) demonstrate remarkable similarities^70–72^. The electrophysiological studies showed that the depolarization current of peripheral motor nerve axons in RLS increases, reduces the threshold for action potential release, promotes spontaneous discharge, and has stronger adaptability to hyperpolarization^73^. And there are similar manifestations in the spinal cord and brain, both with increased excitability^74,75^. The theory of this study unifies these two conditions, proposing that they represent different manifestations of membrane potential abnormalities in the nervous system: sensory symptoms and motor symptoms. RLS may be mapped to increased Type I neuronal excitability (sensory, continuous firing), while PLMD corresponds to increased Type II excitability (motor, burst firing).

### Genetic Susceptibility

CSF analysis and autopsy findings in RLS patients show deficiencies in endogenous opioid. The LDSC analysis supported genetic correlation between the two conditions, while reverse MR analysis supported that RLS was a risk factor for opioids use. These findings suggested that the opioids use risk in RLS patients may be driven by genetic susceptibility. In other words, deficiency in endogenous opioid may predispose individuals to dependency, which can be either physiological or pathological. Physiological dependency can be viewed as a form of drug replacement therapy with sustained and stable therapeutic effects. This provides potential theoretical support for the sustained and stable therapeutic effects of opioid drugs observed in long-term follow-up studies^13,14^. However, the mechanisms underlying membrane potential abnormalities caused by secondary RLS may be more complex, potentially leading to the development of pathological dependency and addiction due to continuous dose escalation when opioids fail to address the underlying problem. This underscored the critical importance of effective assessment and optimal dose management in opioids use among patients with RLS.

In summary, RLS is not a disease per se, but rather a description of clinical manifestations. In fact, this may be a membrane potential disturbance caused by various etiologies. In the central nervous system, neurons and synapses occur hyperexcitability, activating dopaminergic neuron for the regulation negative feedback and analgesia, the deficiency of endogenous opioids may exacerbate this compensatory mechanism, which leads to increased dopamine secretion causes reinforcement of addiction. Although low-threshold ion channels are affected, the human body’s perceptual ability for low-threshold ion channels in sensory nervous system may be superior to other locations, leading to the high proportion of RLS development. Considering the compensatory mechanisms of the dopaminergic system in the brain, it can be further classified into an asymptomatic compensated phase and a symptomatic decompensated phase. As is well known, sensory symptoms include not only discomfort but also an irresistible urge to move the lower limbs, fast relief after activity, and circadian rhythm changes. The fast relief after activity may be due to repolarization and hyperpolarization that occur after action potentials, that is reasonable on a time scale, while simultaneously, symptom relief reduces the dopaminergic burden of abnormally activated reward mechanisms, or symptom relief-dopamine secretion coupling phenomena occur. The strong desire for activity may be driven by addiction mechanism. The circadian rhythm changes may be related to the insufficient expression of *KCNH3/KCNH8*, which are closely related to the regulation of circadian rhythm^47^. In addition, the abnormal baseline of synaptic plasticity and neurotransmitters in long-term pathological states makes understanding this process more complex.

#### Limitations

Several caveats merit mention. First, The GWAS data were predominantly of European ancestry, limiting generalizability; effect sizes and LD may differ in other populations. Second, the temporal mediation model is conceptual and relies on theoretical assumptions, since cross-sectional GWAS cannot capture dynamic changes; longitudinal metabolomic/clinical data would be needed to validate the time course. Third, as exploratory analyses used to uncover clues, I used a single SNP (rs2618039) as the opioid instrument (though it is strong, F=30.6), and a relaxed significance threshold for metabolites. The metabolite-RLS MR results were not multiple testing corrected (which could inflate type I error). Fourth, the opioid-use phenotype encompassed all prescription opioids (ATC N02A) without detail on dose, duration or specific drug, potentially diluting drug-specific genetic effects. Fifth, although there are 19942 OpenGWAS eQTLs, only 50 *KCN*-eQTLs were found, and the 40 effective instrumental variables obtained after screening did not fully cover all *KCN*-subtype-genes. Sixth, “Membrane Potential Disturbance” remains a hypothesis; we lack direct experimental validation. However, its explanatory power is notable, coherently unifying the various unexplainable phenomena of RLS. The convergent evidence from genetic correlation, bidirectional MR, mediation analysis, enrichment and multiple validation streams lends credibility to this model even absent bench experiments.

## Conclusion

This study provides the genetic support and preliminary multi method evaluation for understanding the relationship between potassium channel regulation and RLS, and propose “Membrane Potential Disturbance” hypothesis as the potential pathophysiological mechanism. Potassium channel regulation dysfunction as the potential driver of neuronal hyperexcitability, secondary dopaminergic adaptations, that may offer explanatory value for biological observations. The changes in membrane potential also suggest a new pathway on the connection between iron deficiency caused myelin changes, and dopaminergic system changes. These findings may suggest a new perspective and framework for understanding the pathophysiology of RLS.

## Data Availability

All data produced in the present study are available upon reasonable request to the authors.

## Data availability

All data of the paper can be found in Supplementary Materials. The resources were described in the methods section.

## References

1. Song, P., Wu, J., Cao, J., Sun, W., Li, X., Zhou, T., Shen, Y., Tan, X., Ye, X., Yuan, C., Zhu, Y., Rudan, I., & Global Health Epidemiology Research Group (GHERG). (2024). The global and regional prevalence of restless legs syndrome among adults: A systematic review and modelling analysis. Journal of Global Health, 14. 10.7189/jogh.14.04113

2. Khachatryan, S. G., Ferri, R., Fulda, S., Garcia-Borreguero, D., Manconi, M., Muntean, M., & Stefani, A. (2022). Restless legs syndrome: Over 50 years of European contribution. Journal of Sleep Research, 31(4). 10.1111/jsr.13632

3. Allen, R. P., Connor, J. R., Hyland, K., & Earley, C. J. (2009). Abnormally increased CSF 3-ortho-methyldopa (3-OMD) in untreated restless legs syndrome (RLS) patients indicates more severe disease and possibly abnormally increased dopamine synthesis. Sleep Medicine, 10(1), 123–128. 10.1016/j.sleep.2007.11.012

4. Walters, A. S., Li, Y., Koo, B. B., Ondo, W. G., Weinstock, L. B., Champion, D., Afrin, L. B., Karroum, E. G., Bagai, K., & Spruyt, K. (2024). Review of the role of the endogenous opioid and melanocortin systems in the restless legs syndrome. Brain, 147(1), 26–38. 10.1093/brain/awad283

5. Cosme, D., Estevinho, M. M., Rieder, F., & Magro, F. (2021). Potassium channels in intestinal epithelial cells and their pharmacological modulation: A systematic review. American Journal of Physiology Cell Physiology, 320(4), C520–C546. 10.1152/ajpcell.00393.2020

6. Alam, K. A., Svalastoga, P., Martinez, A., Glennon, J. C., & Haavik, J. (2023). Potassium channels in behavioral brain disorders. Molecular mechanisms and therapeutic potential: A narrative review. Neuroscience & Biobehavioral Reviews, 152, 105301. 10.1016/j.neubiorev.2023.105301

7. Luo, H., Marron Fernandez De Velasco, E., &Wickman, K. (2022). Neuronal G protein-gated K^+^ channels. American Journal of Physiology Cell Physiology, 323(2), C439–C460. 10.1152/ajpcell.00102.2022

8. Drion, G., O’Leary, T., & Marder, E. (2015). Ion channel degeneracy enables robust and tunable neuronal firing rates. Proceedings of the National Academy of Sciences, 112(38). 10.1073/pnas.1516400112

9. Koo, B. B., Abdelfattah, A., Eysa, A., & Lu, L. (2024). The melanocortin and endorphin neuropeptides in patients with restless legs syndrome. Annals of Neurology, 95(4), 688–699. 10.1002/ana.26876

10. Walters, A. S., Ondo, W. G., Zhu, W., & Le, W. (2009). Does the endogenous opiate system play a role in the restless legs syndrome?: A pilot post-mortem study. Journal of the Neurological Sciences, 279(1–2), 62–65. 10.1016/j.jns.2008.12.022

11. Lyu, S., DeAndrade, M. P., Mueller, S., Oksche, A., Walters, A. S., & Li, Y. (2019). Hyperactivity, dopaminergic abnormalities, iron deficiency and anemia in an in vivo opioid receptors knockout mouse: Implications for the restless legs syndrome. Behavioural Brain Research, 374, 112123. 10.1016/j.bbr.2019.112123

12. Trenkwalder, C., Beneš, H., Grote, L., García-Borreguero, D., Högl, B., Hopp, M., Bosse, B., Oksche, A., Reimer, K., Winkelmann, J., Allen, R. P., & Kohnen, R. (2013). Prolonged release oxycodone–naloxone for treatment of severe restless legs syndrome after failure of previous treatment: A double-blind, randomised, placebo-controlled trial with an open-label extension. Lancet Neurology, 12(12), 1141–1150. 10.1016/s1474-4422(13)70239-4

13. Silver, N., Allen, R. P., Senerth, J., & Earley, C. J. (2011). A 10-year, longitudinal assessment of dopamine agonists and methadone in the treatment of restless legs syndrome. Sleep Medicine, 12(5), 440–444. 10.1016/j.sleep.2010.11.002

14. Winkelman, J. W., Wipper, B., & Zackon, J. (2023). Long-term safety, dose stability, and efficacy of opioids for patients with restless legs syndrome in the national RLS opioid registry. Neurology, 100(14). 10.1212/wnl.0000000000206855

15. Reeves, K. C., Shah, N., Muñoz, B., & Atwood, B. K. (2022). Opioid receptor-mediated regulation of neurotransmission in the brain. Frontiers in Molecular Neuroscience, 15, 919773. 10.3389/fnmol.2022.919773

16. Richmond, R. C., & Davey Smith, G. (2022). Mendelian randomization: Concepts and scope. Cold Spring Harbor Perspectives in Medicine, 12(1), a040501. 10.1101/cshperspect.a040501

17. Gkatzionis, A., Burgess, S., & Newcombe, P. J. (2023). Statistical methods for cis -mendelian randomization with two-sample summary-level data. Genetic Epidemiology, 47(1), 3–25. 10.1002/gepi.22506

18. Wu, Y., Byrne, E. M., Zheng, Z., Kemper, K. E., Yengo, L., Mallett, A. J., Yang, J., Visscher, P. M., & Wray, N. R. (2019). Genome-wide association study of medication-use and associated disease in the UK biobank. Nature Communications, 10(1). 10.1038/s41467-019-09572-5

19. Akçimen, F., Chia, R., Saez-Atienzar, S., Ruffo, P., Rasheed, M., Ross, J. P., Liao, C., Ray, A., Dion, P. A., Scholz, S. W., Rouleau, G. A., & Traynor, B. J. (2024). Genomic analysis identifies risk factors in restless legs syndrome. Annals of Neurology, 96(5), 994–1005. 10.1002/ana.27040

20. Schormair, B., Zhao, C., Bell, S., Didriksen, M., Nawaz, M. S., Schandra, N., Stefani, A., Högl, B., Dauvilliers, Y., Bachmann, C. G., Kemlink, D., Sonka, K., Paulus, W., Trenkwalder, C., Oertel, W. H., Hornyak, M., Teder-Laving, M., Metspalu, A., Hadjigeorgiou, G. M., … Winkelmann, J. (2024). Genome-wide meta-analyses of restless legs syndrome yield insights into genetic architecture, disease biology and risk prediction. Nature Genetics, 56(6), 1090–1099. 10.1038/s41588-024-01763-1

21. Kurki, M. I., Karjalainen, J., Palta, P., Sipilä, T. P., Kristiansson, K., Donner, K. M., Reeve, M. P., Laivuori, H., Aavikko, M., Kaunisto, M. A., Loukola, A., Lahtela, E., Mattsson, H., Laiho, P., Della Briotta Parolo, P., Lehisto, A. A., Kanai, M., Mars, N., Rämö, J., … Palotie, A. (2023). FinnGen provides genetic insights from a well-phenotyped isolated population. Nature, 613(7944), 508–518. 10.1038/s41586-022-05473-8

22. Wang, C., Yang, C., Western, D., Ali, M., Wang, Y., Phuah, C.-L., Budde, J., Wang, L., Gorijala, P., Timsina, J., Ruiz, A., Pastor, P., Fernandez, M. V., Dominantly Inherited Alzheimer Network (DIAN), Perrin, R., The Alzheimer’s Disease Neuroimaging Initiative (ADNI), Panyard, D. J., Engelman, C. D., Deming, Y., … Cruchaga, C. (2024). Genetic architecture of cerebrospinal fluid and brain metabolite levels and the genetic colocalization of metabolites with human traits. Nature Genetics, 56(12), 2685–2695. 10.1038/s41588-024-01973-7

23. Ding, R., Wang, Q., Gong, L., Zhang, T., Zou, X., Xiong, K., Liao, Q., Plass, M., & Li, L. (2024). scQTLbase: An integrated human single-cell eQTL database. Nucleic Acids Research, 52(D1), D1010–D1017. 10.1093/nar/gkad781

24. Bryois, J., Calini, D., Macnair, W., Foo, L., Urich, E., Ortmann, W., Iglesias, V. A., Selvaraj, S., Nutma, E., Marzin, M., Amor, S., Williams, A., Castelo-Branco, G., Menon, V., De Jager, P., & Malhotra, D. (2022). Cell-type-specific cis-eQTLs in eight human brain cell types identify novel risk genes for psychiatric and neurological disorders. Nature Neuroscience, 25(8), 1104–1112. 10.1038/s41593-022-01128-z

25. Kittke, V., Zhao, C., Lam, D. D., Harrer, P., Krezel, W., Schormair, B., Oexle, K., & Winkelmann, J. (2024). RLS-associated MEIS transcription factors control distinct processes in human neural stem cells. Scientific Reports, 14(1), 28986. 10.1038/s41598-024-80266-9

26. Schizophrenia Working Group of the Psychiatric Genomics Consortium, Bulik-Sullivan, B. K., Loh, P.-R., Finucane, H. K., Ripke, S., Yang, J., Patterson, N., Daly, M. J., Price, A. L., & Neale, B. M. (2015). LD score regression distinguishes confounding from polygenicity in genome-wide association studies. Nature Genetics, 47(3), 291–295. 10.1038/ng.3211

27. Verbanck, M., Chen, C.-Y., Neale, B., & Do, R. (2018). Detection of widespread horizontal pleiotropy in causal relationships inferred from mendelian randomization between complex traits and diseases. Nature Genetics, 50(5), 693–698. 10.1038/s41588-018-0099-7

28. Cohen, J. F., Chalumeau, M., Cohen, R., Korevaar, D. A., Khoshnood, B., & Bossuyt, P. M. M. (2015). Cochran’s Q test was useful to assess heterogeneity in likelihood ratios in studies of diagnostic accuracy. Journal of Clinical Epidemiology, 68(3), 299–306. 10.1016/j.jclinepi.2014.09.005

29. Bowden, J., Davey Smith, G., & Burgess, S. (2015). Mendelian randomization with invalid instruments: Effect estimation and bias detection through egger regression. International Journal of Epidemiology, 44(2), 512–525. 10.1093/ije/dyv080

30. Zuber, V., Grinberg, N. F., Gill, D., Manipur, I., Slob, E. A. W., Patel, A., Wallace, C., & Burgess, S. (2022). Combining evidence from mendelian randomization and colocalization: Review and comparison of approaches. American Journal of Human Genetics, 109(5), 767–782. 10.1016/j.ajhg.2022.04.001

31. Zhu, Z., Zhang, F., Hu, H., Bakshi, A., Robinson, M. R., Powell, J. E., Montgomery, G. W., Goddard, M. E., Wray, N. R., Visscher, P. M., & Yang, J. (2016). Integration of summary data from GWAS and eQTL studies predicts complex trait gene targets. Nature Genetics, 48(5), 481–487. 10.1038/ng.3538

32. Bender, R., & Lange, S. (2001). Adjusting for multiple testing—When and how? Journal of Clinical Epidemiology, 54(4), 343–349. 10.1016/S0895-4356(00)00314-0

33. De Vries, F., Bruin, M., Lobatto, D. J., Dekkers, O. M., Schoones, J. W., Van Furth, W. R., Pereira, A. M., Karavitaki, N., Biermasz, N. R., & Zamanipoor Najafabadi, A. H. (2020). Opioids and their endocrine effects: A systematic review and meta-analysis. Journal of Clinical Endocrinology and Metabolism, 105(4), 1020–1029. 10.1210/clinem/dgz022

34. Patel, E., & Ben-Shlomo, A. (2024). Opioid-induced adrenal insufficiency: Diagnostic and management considerations. Frontiers in Endocrinology, 14, 1280603. 10.3389/fendo.2023.1280603

35. Mikasova, L., Xiong, H., Kerkhofs, A., Bouchet, D., Krugers, H. J., & Groc, L. (2017). Stress hormone rapidly tunes synaptic NMDA receptor through membrane dynamics and mineralocorticoid signalling. Scientific Reports, 7(1), 8053. 10.1038/s41598-017-08695-3

36. Tse, Y. C., Bagot, R. C., & Wong, T. P. (2012). Dynamic regulation of NMDAR function in the adult brain by the stress hormone corticosterone. Frontiers in Cellular Neuroscience, 6. 10.3389/fncel.2012.00009

37. Schilling, C., Schredl, M., Strobl, P., & Deuschle, M. (2010). Restless legs syndrome: Evidence for nocturnal hypothalamic-pituitary-adrenal system activation. Movement Disorders, 25(8), 1047–1052. 10.1002/mds.23026

38. Parandoosh, Z., & Johanson, C. E. (1982). Ontogeny of blood-brain barrier permeability to, and cerebrospinal fluid sink action on, [14C]urea. American Journal of Physiology-Regulatory, Integrative and Comparative Physiology, 243(3), R400–R407. 10.1152/ajpregu.1982.243.3.r400

39. Franco-Pérez, J., Manjarrez-Marmolejo, J., Ballesteros-Zebadúa, P., Neri-Santos, A., Montes, S., Suarez-Rivera, N., Hernández-Cerón, M., & Pérez-Koldenkova, V. (2018). Chronic consumption of fructose induces behavioral alterations by increasing orexin and dopamine levels in the rat brain. Nutrients, 10(11), 1722. 10.3390/nu10111722

40. Flores Monar, G. V., Sanchez Cruz, C., & Calderon Martinez, E. (2025). Mindful eating: A deep insight into fructose metabolism and its effects on appetite regulation and brain function. Journal of Nutrition and Metabolism, 2025(1). 10.1155/jnme/5571686

41. Bryant, L., Doyle, T., Chen, Z., Cuzzocrea, S., Masini, E., Vinci, M. C., Esposito, E., Mazzon, E., Petrusca, D. N., Petrache, I., & Salvemini, D. (2009). Spinal ceramide and neuronal apoptosis in morphine antinociceptive tolerance. Neuroscience Letters, 463(1), 49–53. 10.1016/j.neulet.2009.07.051

42. Brodowicz, J., Przegaliński, E., Müller, C. P., & Filip, M. (2018). Ceramide and its related neurochemical networks as targets for some brain disorder therapies. Neurotoxicity Research, 33(2), 474–484. 10.1007/s12640-017-9798-6

43. Pulst, S., & Otis, T. S. (2012). Repolarization matters: Mutations in the Kv4.3 potassium channel cause SCA19/22. Annals of Neurology, 72(6), 829–831. 10.1002/ana.23803

44. Giudicessi, J. R., Ye, D., Tester, D. J., Crotti, L., Mugione, A., Nesterenko, V. V., Albertson, R. M., Antzelevitch, C., Schwartz, P. J., & Ackerman, M. J. (2011). Transient outward current (ito) gain-of-function mutations in the KCND3-encoded Kv4.3 potassium channel and brugada syndrome. Heart Rhythm, 8(7), 1024–1032. 10.1016/j.hrthm.2011.02.021

45. Teumer, A., Trenkwalder, T., Kessler, T., Jamshidi, Y., Van Den Berg, M. E., Kaess, B., Nelson, C. P., Bastiaenen, R., De Bortoli, M., Rossini, A., Deisenhofer, I., Stark, K., Assa, S., Braund, P. S., Cabrera, C., Dominiczak, A. F., Gögele, M., Hall, L. M., Ikram, M. A., … Reinhard, W. (2019). KCND3 potassium channel gene variant confers susceptibility to electrocardiographic early repolarization pattern. JCI Insight, 4(23). 10.1172/jci.insight.131156

46. Alexander, S. P. H., Mathie, A., Peters, J. A., Veale, E. L., Striessnig, J., Kelly, E., Armstrong, J. F., Faccenda, E., Harding, S. D., Pawson, A. J., Sharman, J. L., Southan, C., Davies, J. A., & CGTP Collaborators. (2019). THE CONCISE GUIDE TO PHARMACOLOGY 2019/20: Ion channels. British Journal of Pharmacology, 176(S1). 10.1111/bph.14749

47. Hermanstyne, T. O., Yang, N.-D., Granados-Fuentes, D., Li, X., Mellor, R. L., Jegla, T., Herzog, E. D., & Nerbonne, J. M. (2023). Kv12-encoded K+ channels drive the day–night switch in the repetitive firing rates of SCN neurons. Journal of General Physiology, 155(9), e202213310. 10.1085/jgp.202213310

48. Iyer, R., Ungless, M. A., & Faisal, A. A. (2017). Calcium-activated SK channels control firing regularity by modulating sodium channel availability in midbrain dopamine neurons. Scientific Reports, 7(1), 5248. 10.1038/s41598-017-05578-5

49. Canavier, C. C., & Landry, R. S. (2006). An increase in AMPA and a decrease in SK conductance increase burst firing by different mechanisms in a model of a dopamine neuron in vivo. Journal of Neurophysiology, 96(5), 2549–2563. 10.1152/jn.00704.2006

50. Guzman, J. N., Sánchez-Padilla, J., Chan, C. S., & Surmeier, D. J. (2009). Robust pacemaking in substantia nigra dopaminergic neurons. Journal of Neuroscience, 29(35), 11011–11019. 10.1523/JNEUROSCI.2519-09.2009

51. Khaliq, Z. M., & Bean, B. P. (2010). Pacemaking in dopaminergic ventral tegmental area neurons: Depolarizing drive from background and voltage-dependent sodium conductances. Journal of Neuroscience, 30(21), 7401–7413. 10.1523/JNEUROSCI.0143-10.2010

52. Friedman, A. K., Walsh, J. J., Juarez, B., Ku, S. M., Chaudhury, D., Wang, J., Li, X., Dietz, D. M., Pan, N., Vialou, V. F., Neve, R. L., Yue, Z., & Han, M.-H. (2014). Enhancing depression mechanisms in midbrain dopamine neurons achieves homeostatic resilience. Science, 344(6181), 313–319. 10.1126/science.1249240

53. Liu, E., Pang, K., Liu, M., Tan, X., Hang, Z., Mu, S., Han, W., Yue, Q., Comai, S., & Sun, J. (2023). Activation of Kv7 channels normalizes hyperactivity of the VTA-NAcLat circuit and attenuates methamphetamine-induced conditioned place preference and sensitization in mice. Molecular Psychiatry, 28(12), 5183–5194. 10.1038/s41380-023-02218-5

54. Wang, H.-R., Hu, S.-W., Zhang, S., Song, Y., Wang, X.-Y., Wang, L., Li, Y.-Y., Yu, Y.-M., Liu, H., Liu, D., Ding, H.-L., & Cao, J.-L. (2021). KCNQ channels in the mesolimbic reward circuit regulate nociception in chronic pain in mice. Neuroscience Bulletin, 37(5), 597–610. 10.1007/s12264-021-00668-x

55. Wise, R. A., & Jordan, C. J. (2021). Dopamine, behavior, and addiction. Journal of Biomedical Science, 28(1). 10.1186/s12929-021-00779-7

56. Huang, M., Wang, G., Lin, Y., Guo, Y., Ren, X., Shao, J., Cao, J., Zang, W., & Li, Z. (2022). Dopamine receptor D2, but not D1, mediates the reward circuit from the ventral tegmental area to the central amygdala, which is involved in pain relief. Molecular Pain, 18, 17448069221145096. 10.1177/17448069221145096

57. Taylor, A. M. W., Becker, S., Schweinhardt, P., & Cahill, C. (2016). Mesolimbic dopamine signaling in acute and chronic pain: Implications for motivation, analgesia, and addiction. Pain, 157(6), 1194–1198. 10.1097/j.pain.0000000000000494

58. McCarter, S. J., Labott, J. R., Mazumder, M. K., Gebhard, J., Cunningham, J. L., Loukianova, L. L., Gilliam, W. P., & Lipford, M. C. (2023). Emergence of restless legs syndrome during opioid discontinuation. Journal of Clinical Sleep Medicine, 19(4), 741–748. 10.5664/jcsm.10436

59. Mackie, S. E., McHugh, R. K., McDermott, K., Griffin, M. L., Winkelman, J. W., & Weiss, R. D. (2017). Prevalence of restless legs syndrome during detoxification from alcohol and opioids. Journal of Substance Abuse Treatment, 73, 35–39. 10.1016/j.jsat.2016.10.001

60. McCoy, M. T., Jayanthi, S., & Cadet, J. L. (2021). Potassium channels and their potential roles in substance use disorders. International Journal of Molecular Sciences, 22(3), 1249. 10.3390/ijms22031249

61. You, C., Vandegrift, B. J., & Brodie, M. S. (2021). KCNK13 potassium channels in the ventral tegmental area of rats are important for excitation of ventral tegmental area neurons by ethanol. Alcoholism: Clinical and Experimental Research, 45(7), 1348–1358. 10.1111/acer.14630

62. Connor, J. R., Patton, S. M., Oexle, K., & Allen, R. P. (2017). Iron and restless legs syndrome: Treatment, genetics and pathophysiology. Sleep Medicine, 31, 61–70. 10.1016/j.sleep.2016.07.028

63. Dowling, P., Klinker, F., Amaya, F., Paulus, W., & Liebetanz, D. (2009). Iron-deficiency sensitizes mice to acute pain stimuli and formalin-induced nociception ,. Journal of Nutrition, 139(11), 2087–2092. 10.3945/jn.109.112557

64. Yoo, J. J., Hayes, M., Serafin, E. K., & Baccei, M. L. (2023). Early-life iron deficiency persistently alters nociception in developing mice. Journal of Pain, 24(8), 1321–1336. 10.1016/j.jpain.2023.03.012

65. Reinert, A., Morawski, M., Seeger, J., Arendt, T., & Reinert, T. (2019). Iron concentrations in neurons and glial cells with estimates on ferritin concentrations. BMC Neuroscience, 20, 25. 10.1186/s12868-019-0507-7

66. Cheli, V. T., Correale, J., Paez, P. M., & Pasquini, J. M. (2020). Iron metabolism in oligodendrocytes and astrocytes, implications for myelination and remyelination. ASN NEURO, 12(1), 1759091420962681. 10.1177/1759091420962681

67. Wade, Q. W., & Connor, J. R. (2025). What does iron mean to an oligodendrocyte? Glia, 73(9), 1784–1804. 10.1002/glia.70043

68. Kim, W., & Angulo, M. C. (2025). Unraveling the role of oligodendrocytes and myelin in pain. Journal of Neurochemistry, 169(1), e16206. 10.1111/jnc.16206

69. Connor, J. R., Ponnuru, P., Lee, B.-Y., Podskalny, G. D., Alam, S., Allen, R. P., Earley, C. J., & Yang, Q. X. (2011). Postmortem and imaging based analyses reveal CNS decreased myelination in restless legs syndrome. Sleep Medicine, 12(6), 614–619. 10.1016/j.sleep.2010.10.009

70. Riccardi, S., Ferri, R., Garbazza, C., Miano, S., & Manconi, M. (2023). Pharmacological responsiveness of periodic limb movements in patients with restless legs syndrome: A systematic review and meta-analysis. Journal of Clinical Sleep Medicine, 19(4), 811–822. 10.5664/jcsm.10440

71. Edelson, J. L., Schneider, L. D., Amar, D., Brink-Kjaer, A., Cederberg, K. L., Kutalik, Z., Hagen, E. W., Peppard, P. E., Tempaku, P. F., Tufik, S., Evans, D. S., Stone, K., Tranah, G., Cade, B., Redline, S., Haba-Rubio, J., Heinzer, R., Marques-Vidal, P., Vollenweider, P., … Mignot, E. (2023). The genetic etiology of periodic limb movement in sleep. Sleep, 46(4). 10.1093/sleep/zsac121

72. Van Der Veen, S., Caviness, J. N., Dreissen, Y. E. M., Ganos, C., Ibrahim, A., Koelman, J. H. T. M., Stefani, A., & Tijssen, M. A. J. (2022). Myoclonus and other jerky movement disorders. Clinical Neurophysiology Practice, 7, 285–316. 10.1016/j.cnp.2022.09.003

73. Czesnik, D., Howells, J., Bartl, M., Veiz, E., Ketzler, R., Kemmet, O., Walters, A. S., Trenkwalder, C., Burke, D., & Paulus, W. (2019). I_h_ contributes to increased motoneuron excitability in restless legs syndrome. The Journal of Physiology, 597(2), 599–609. 10.1113/jp275341

74. Dafkin, C., McKinon, W., & Kerr, S. (2019). Restless legs syndrome: Clinical changes in nervous system excitability at the spinal cord level. Sleep Medicine Reviews, 47, 9–17. 10.1016/j.smrv.2019.05.005

75. Kim, T.-J., Cha, K. S., Lee, S., Yang, T.-W., Kim, K. T., Park, B.-S., Jun, J.-S., Lim, J.-A., Byun, J.-I., Sunwoo, J.-S., Shin, J.-W., Kim, K. H., Lee, S. K., & Jung, K.-Y. (2020). Brain regions associated with periodic leg movements during sleep in restless legs syndrome. Scientific Reports, 10(1). 10.1038/s41598-020-58365-0

