## Supplementary figures and images for "Potassium Channel Regulation Dysfunction as a Potential Driving Factor of Membrane Potential Disturbance in Restless Legs Syndrome"

### combined_plots.png

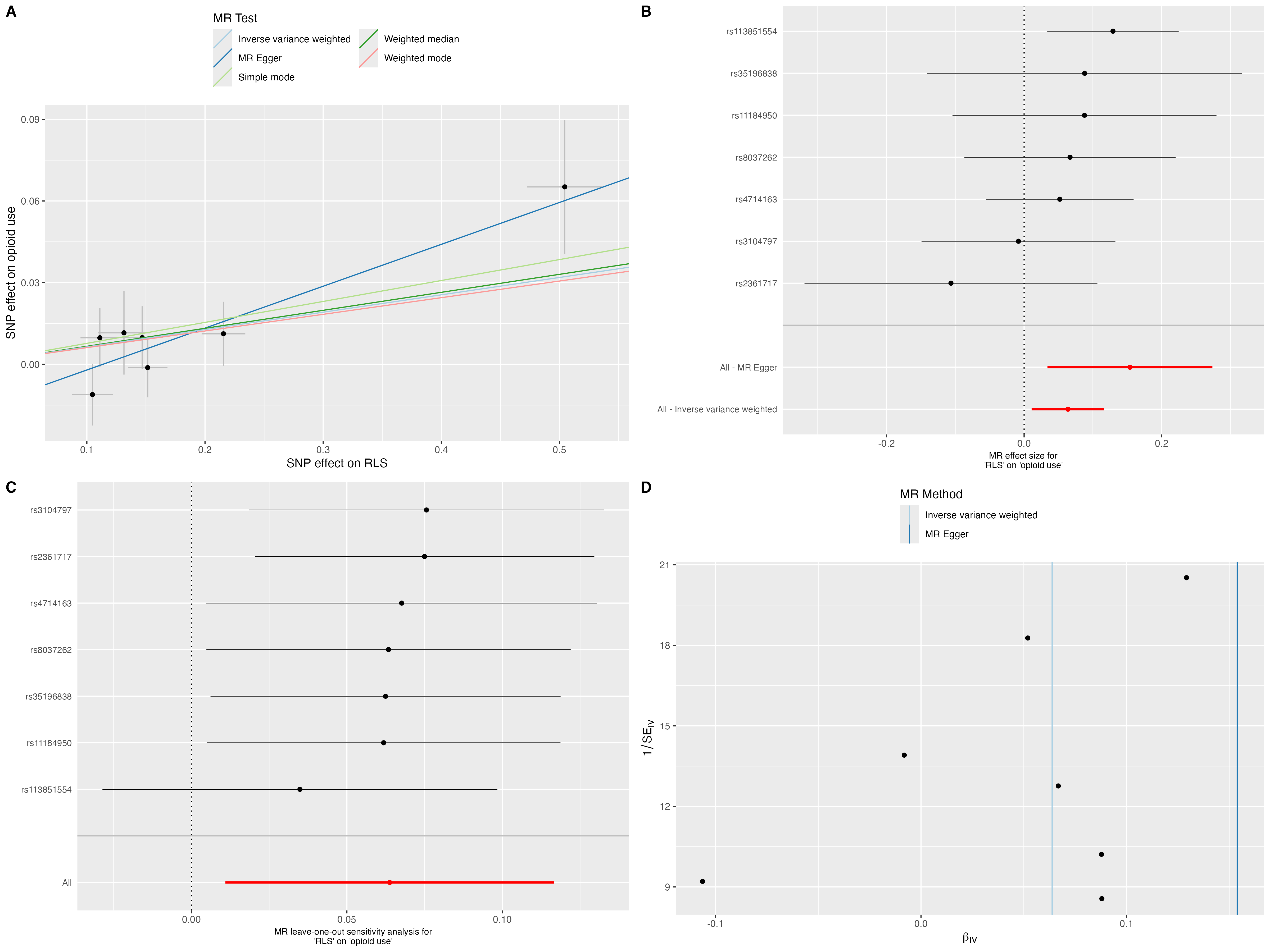

### fig1.png

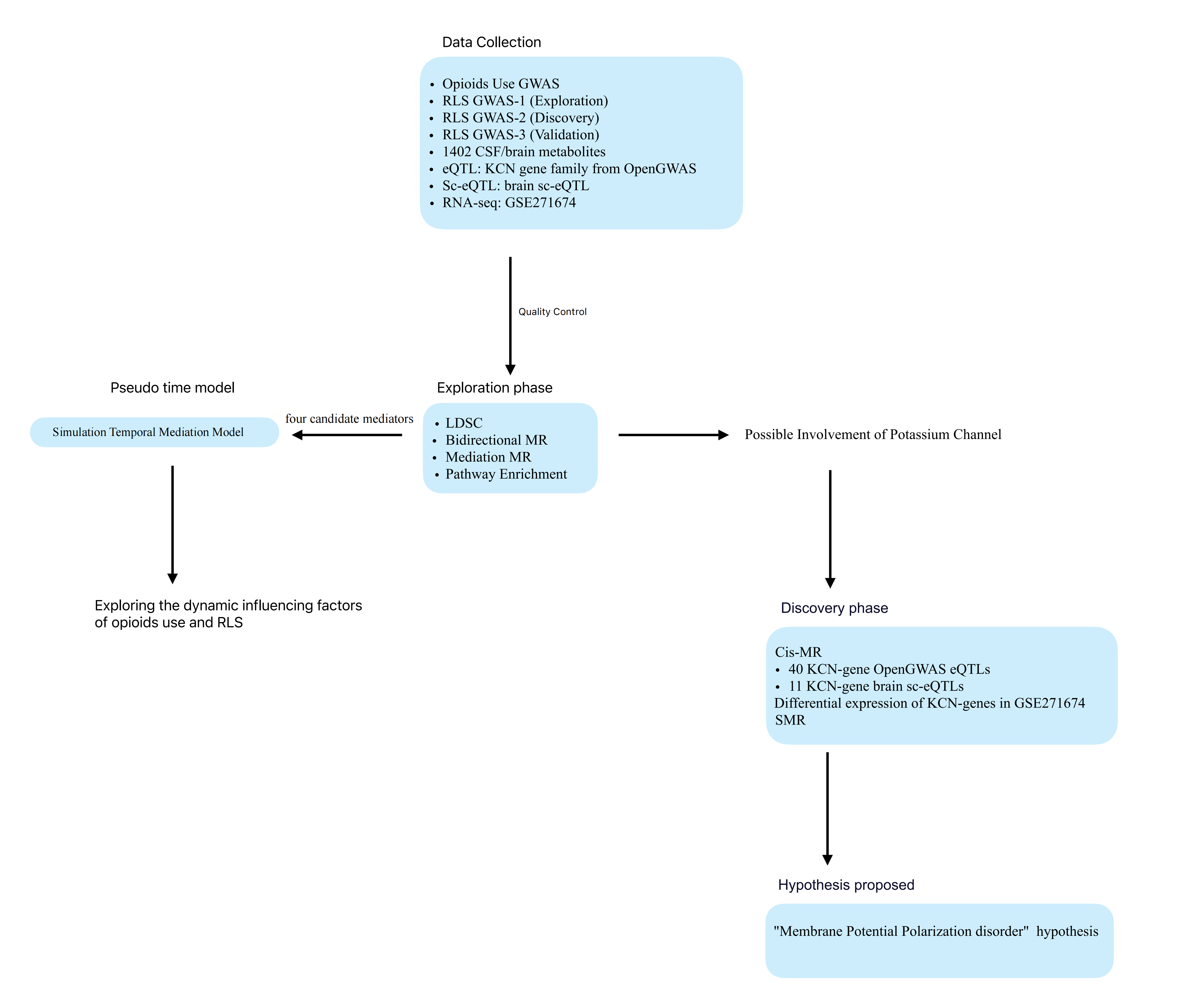

### fig2.png

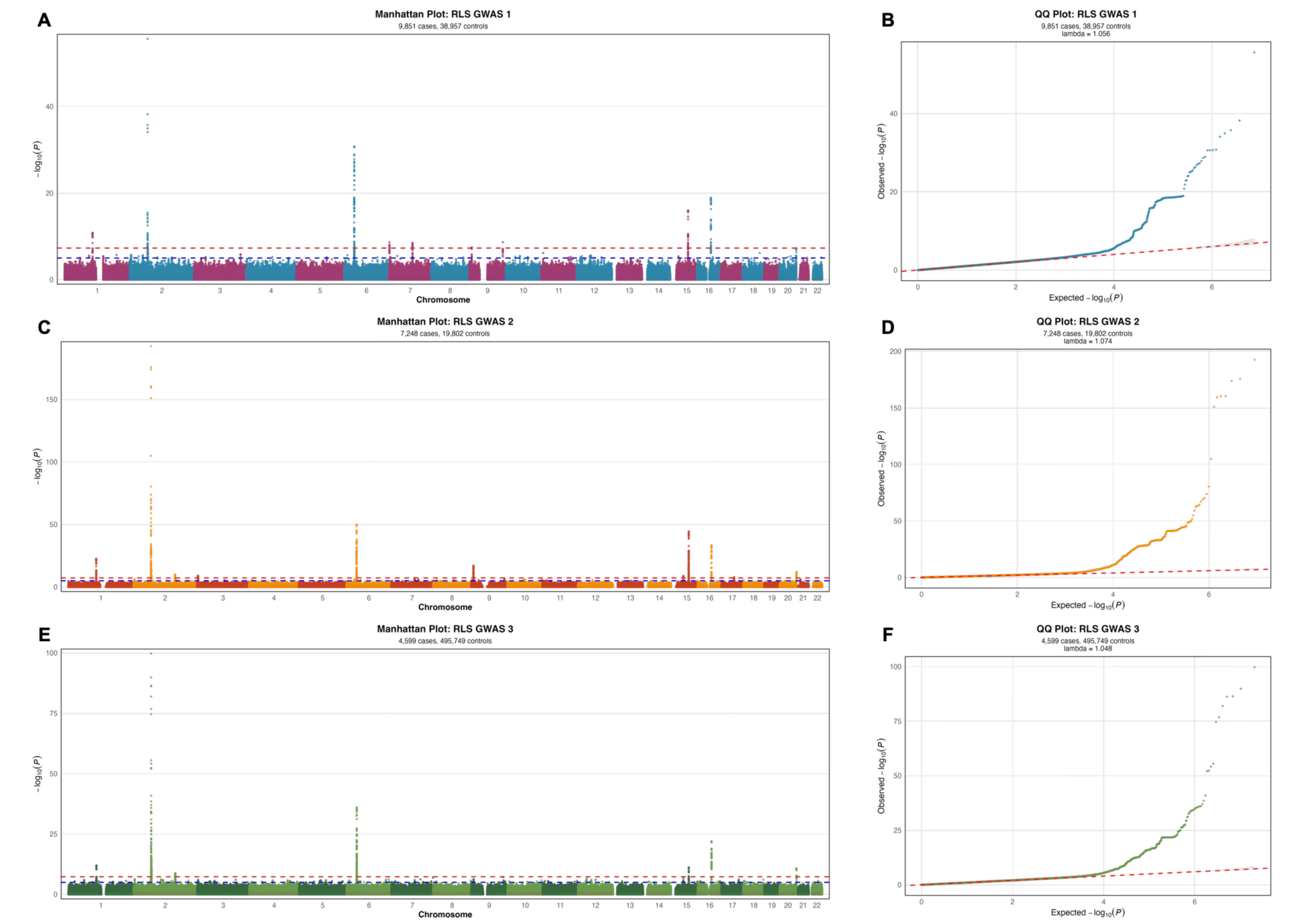

### fig3.png

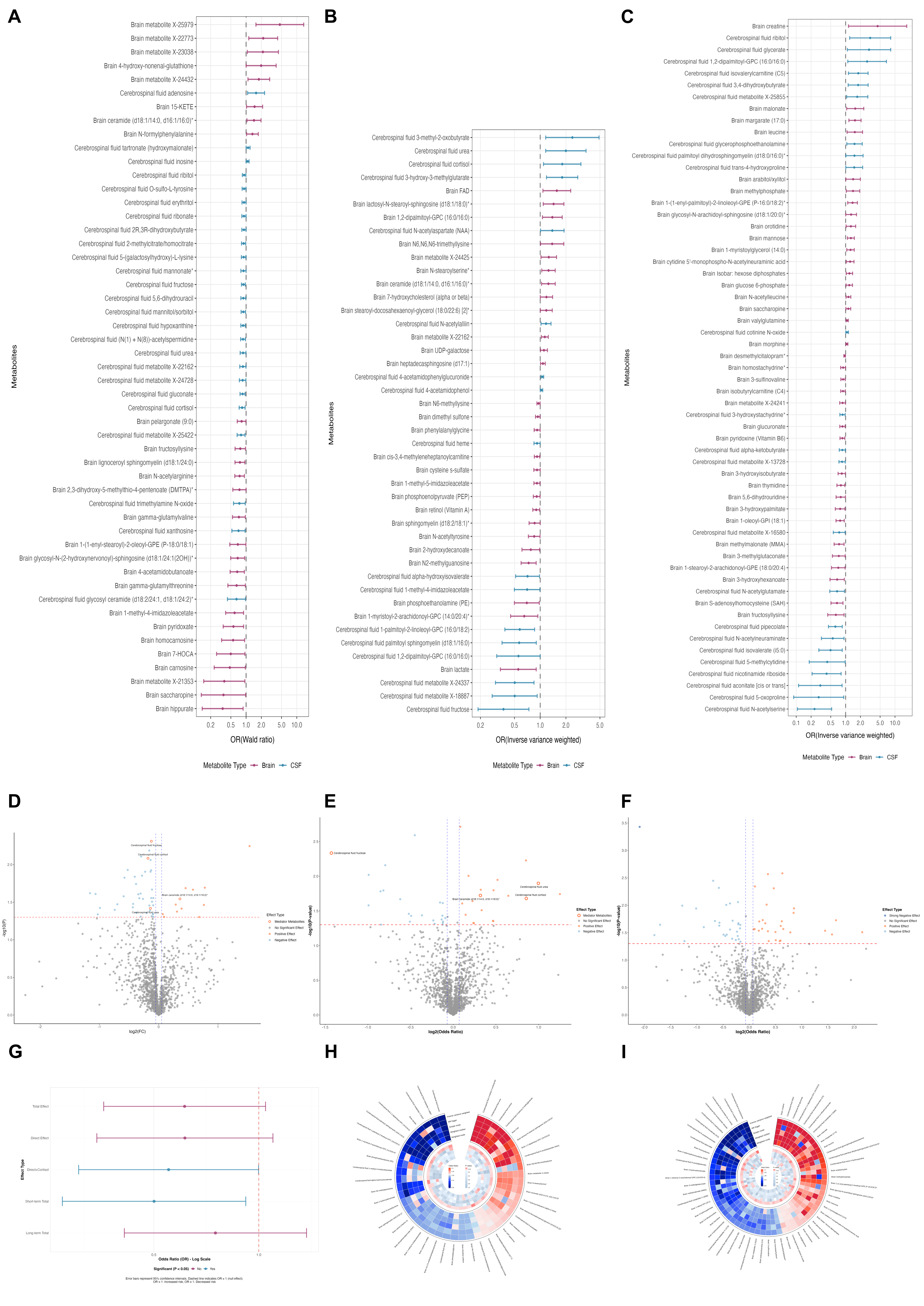

### fig4.png

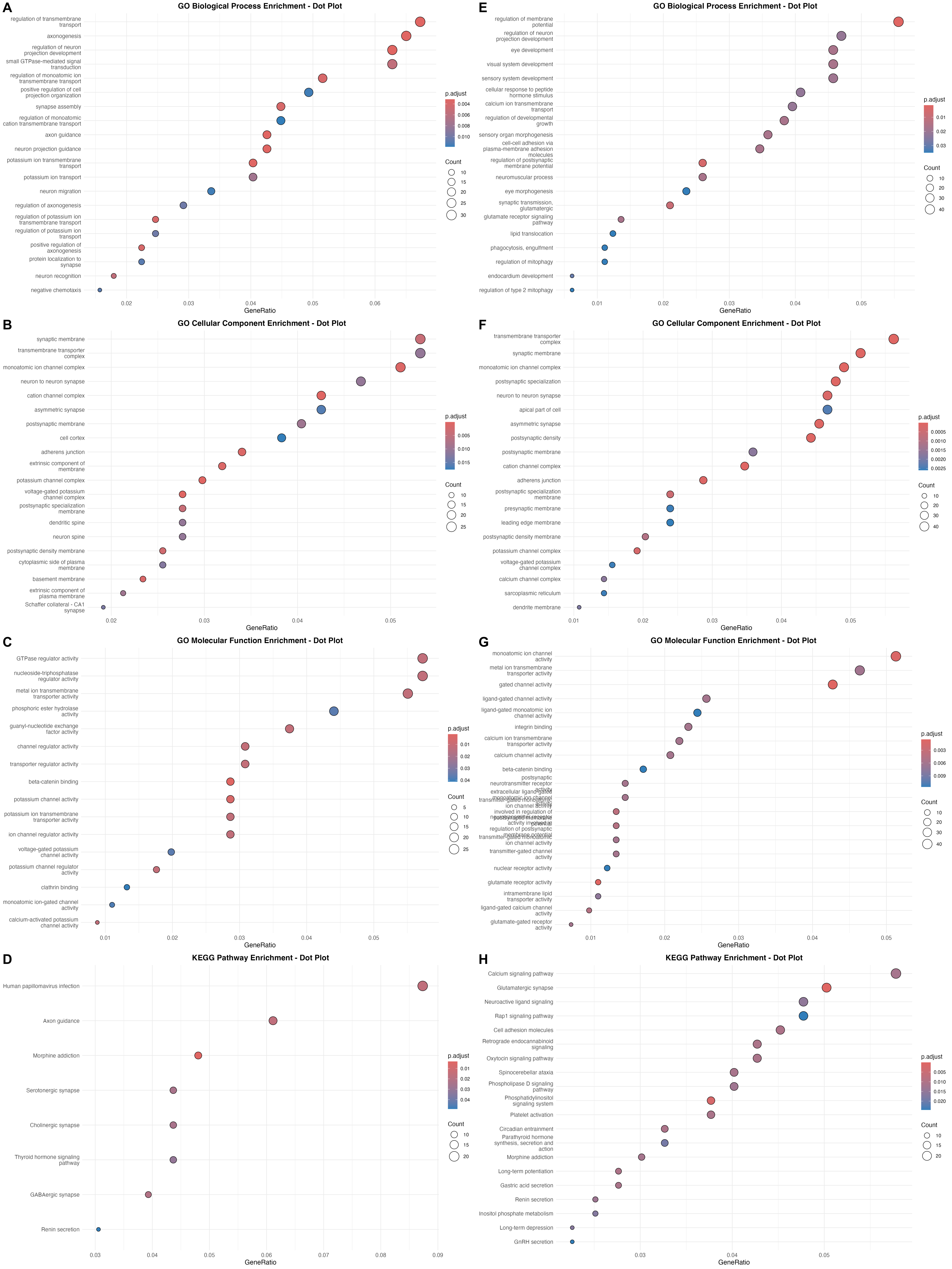

### fig5.png

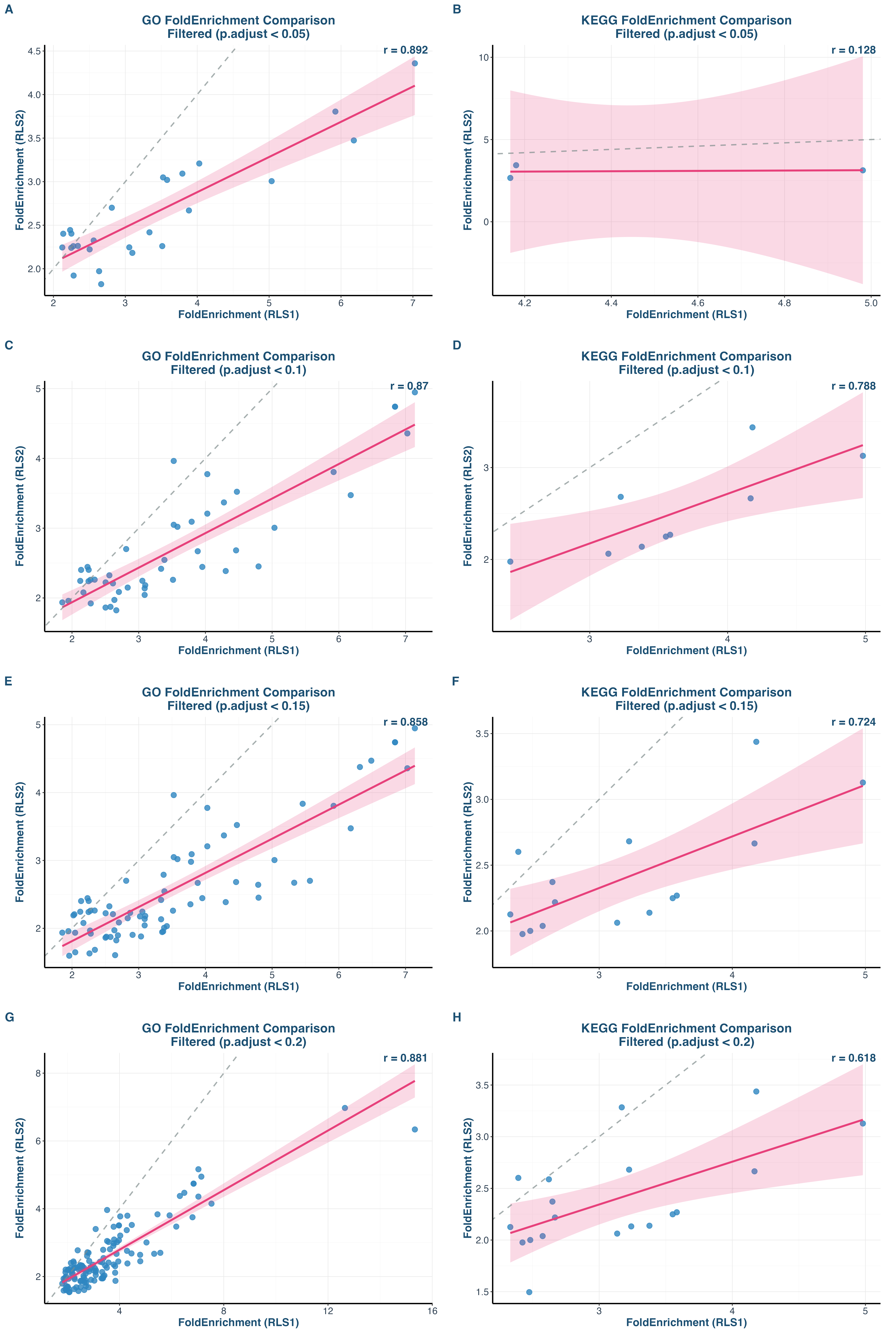

### fig6.png

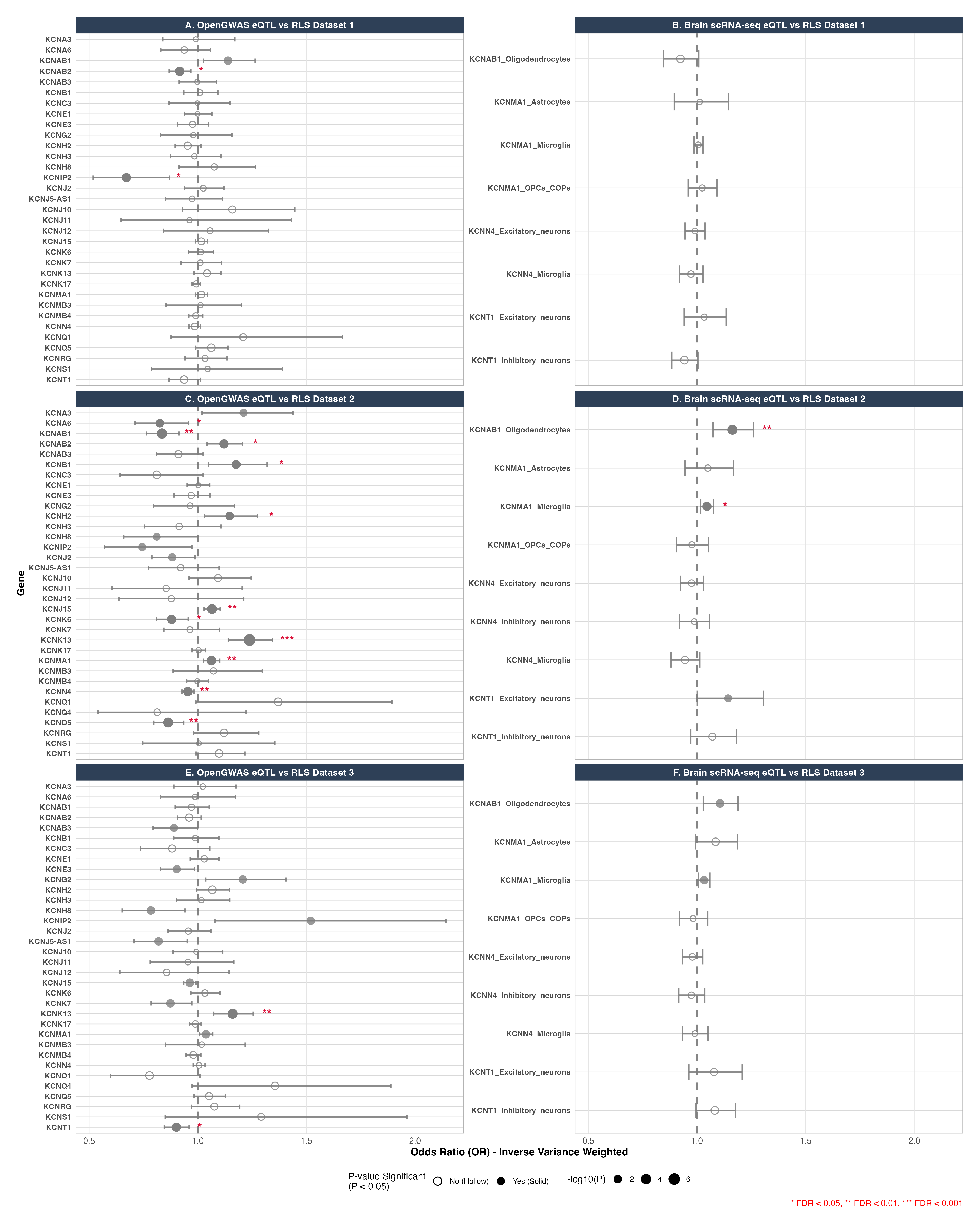

### fig7.png

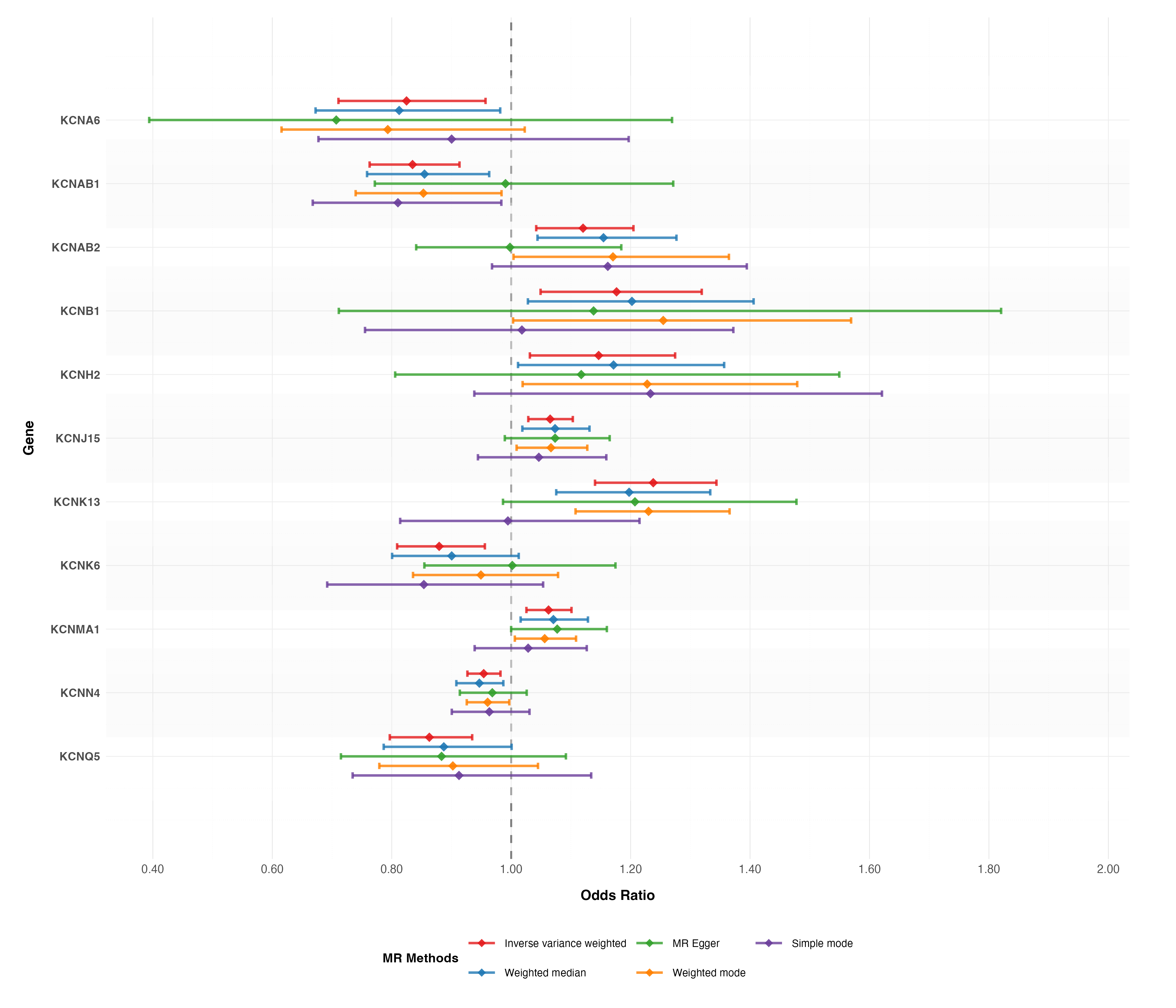

### fig8.png

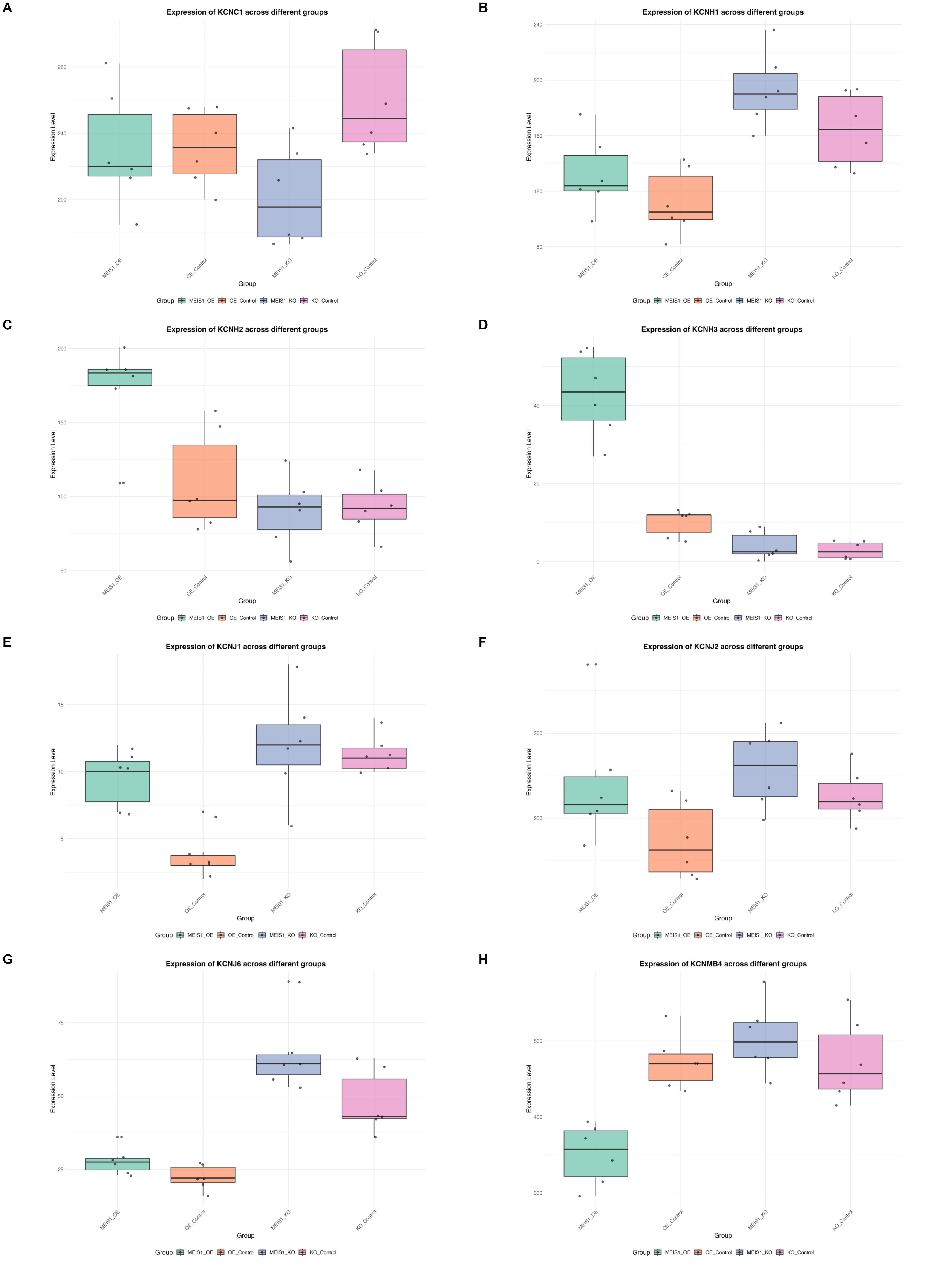

### forest_plot.png

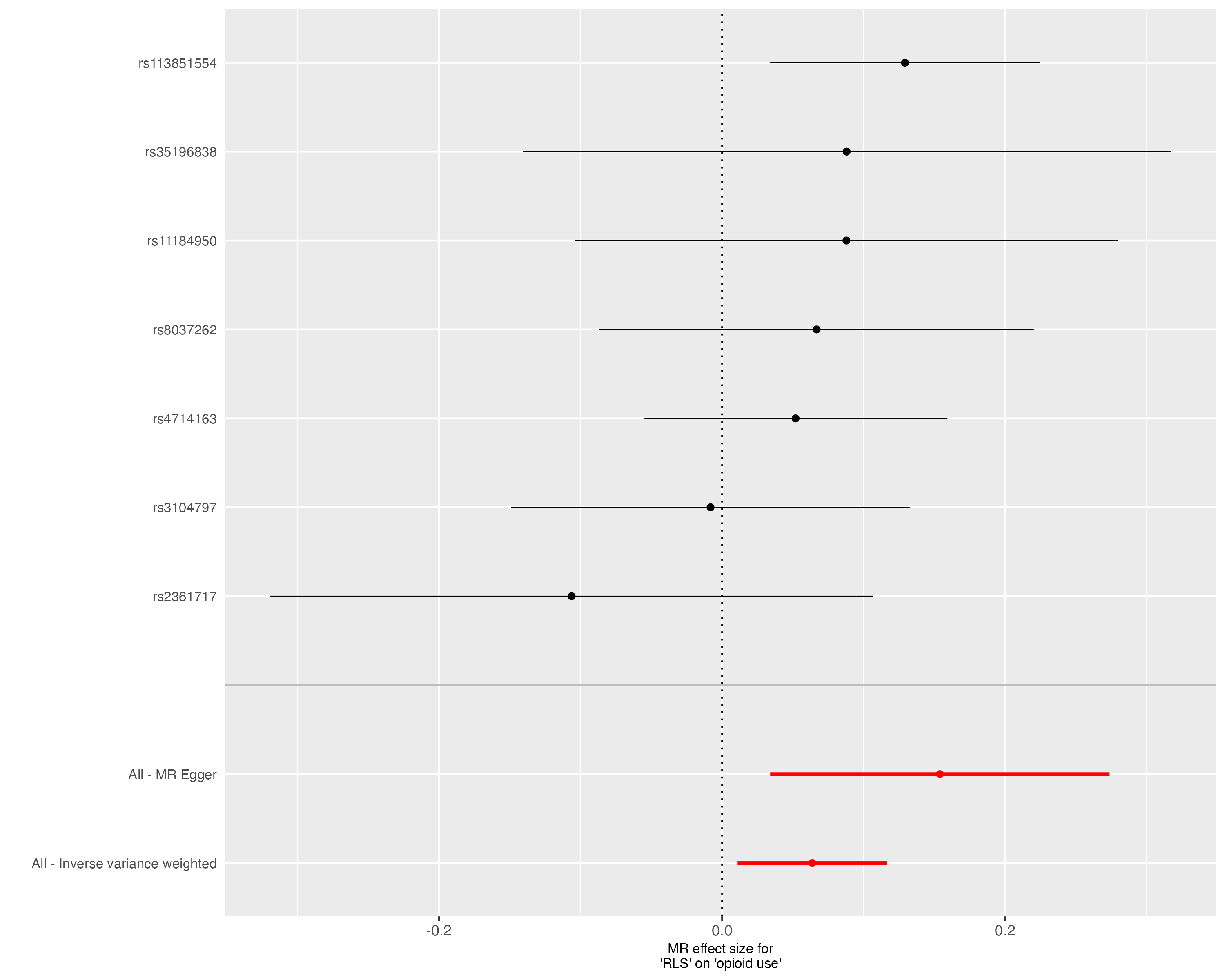

### forest_plot.png

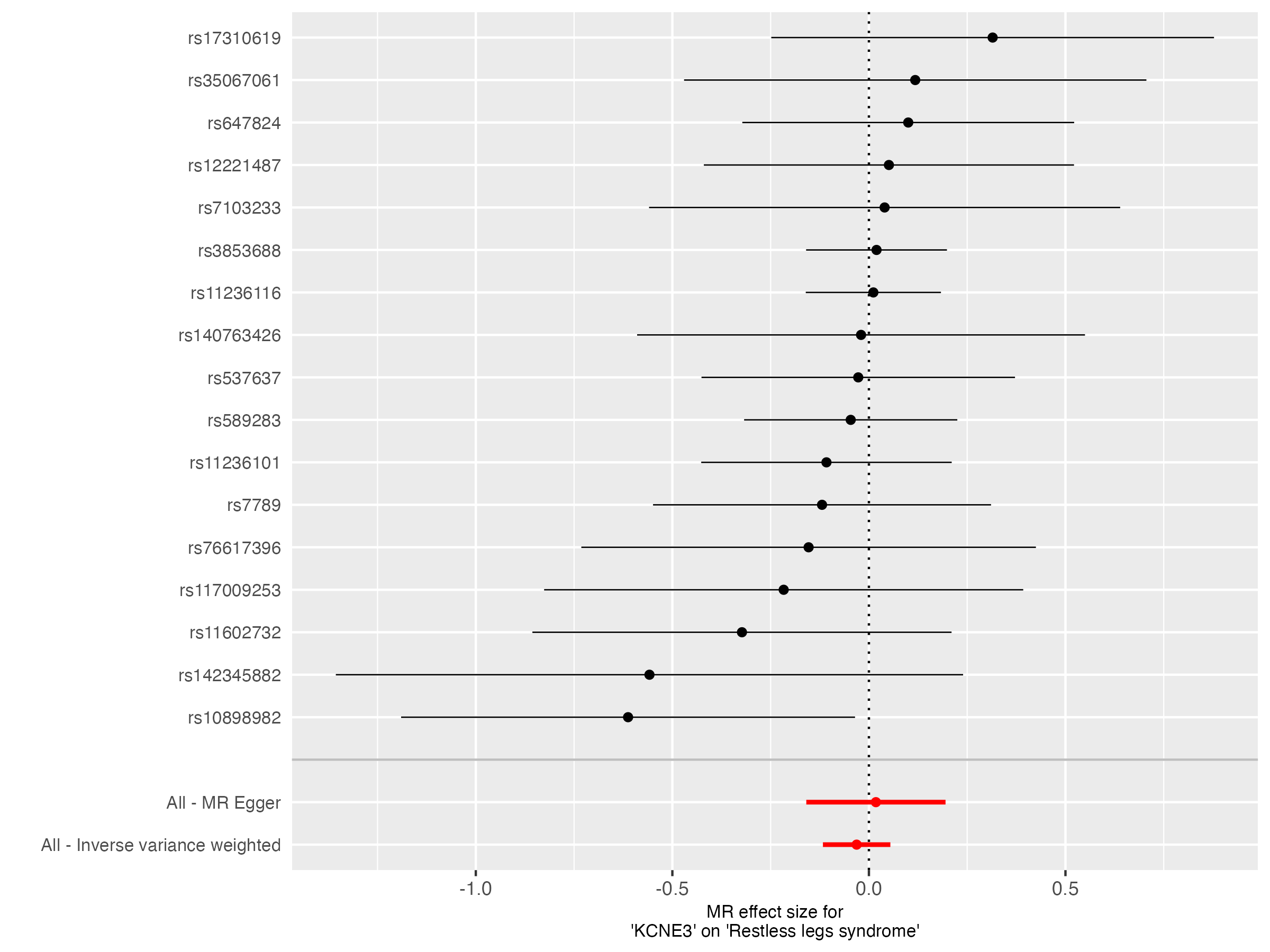

### forest_plot.png

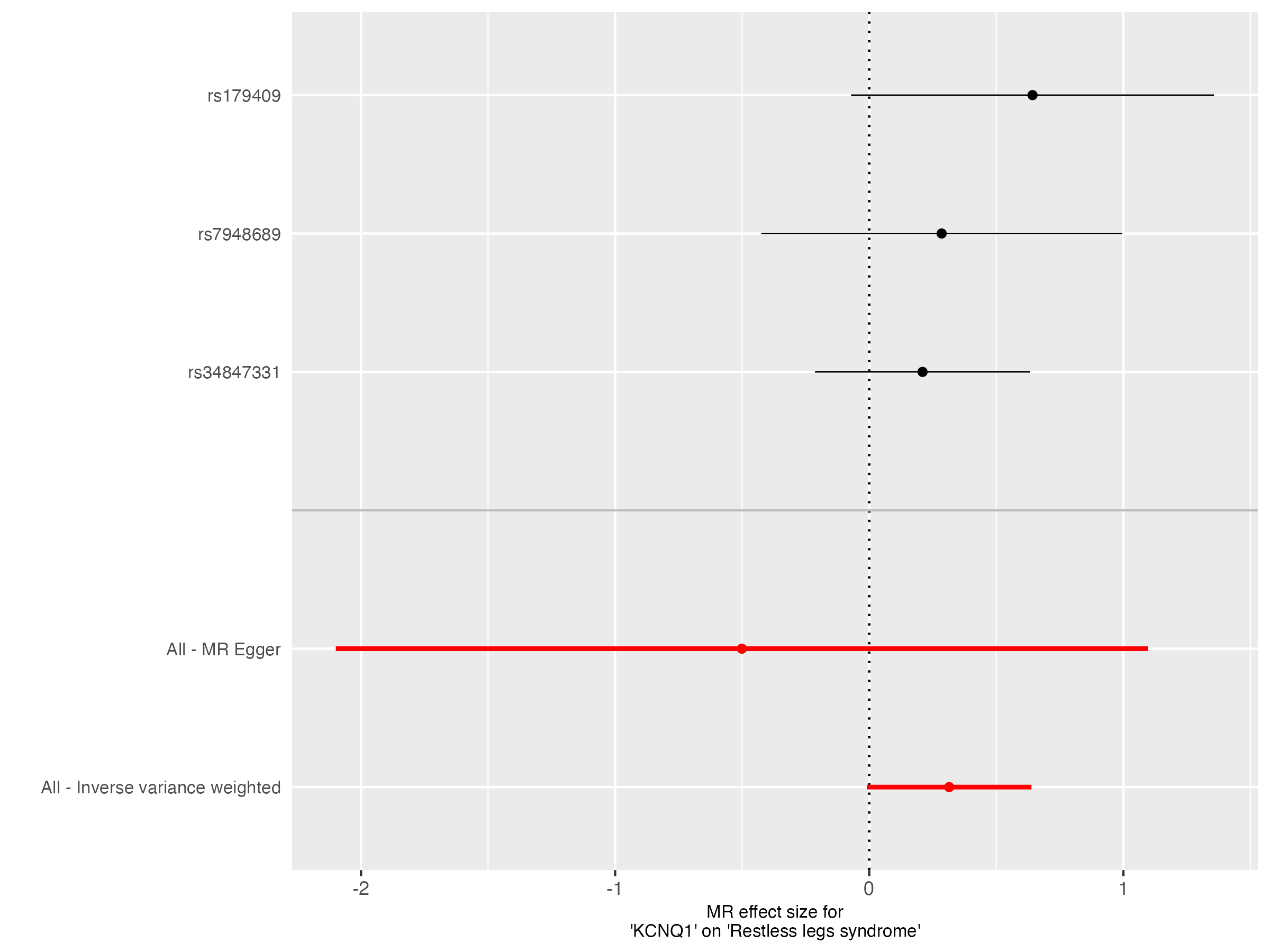

### forest_plot.png

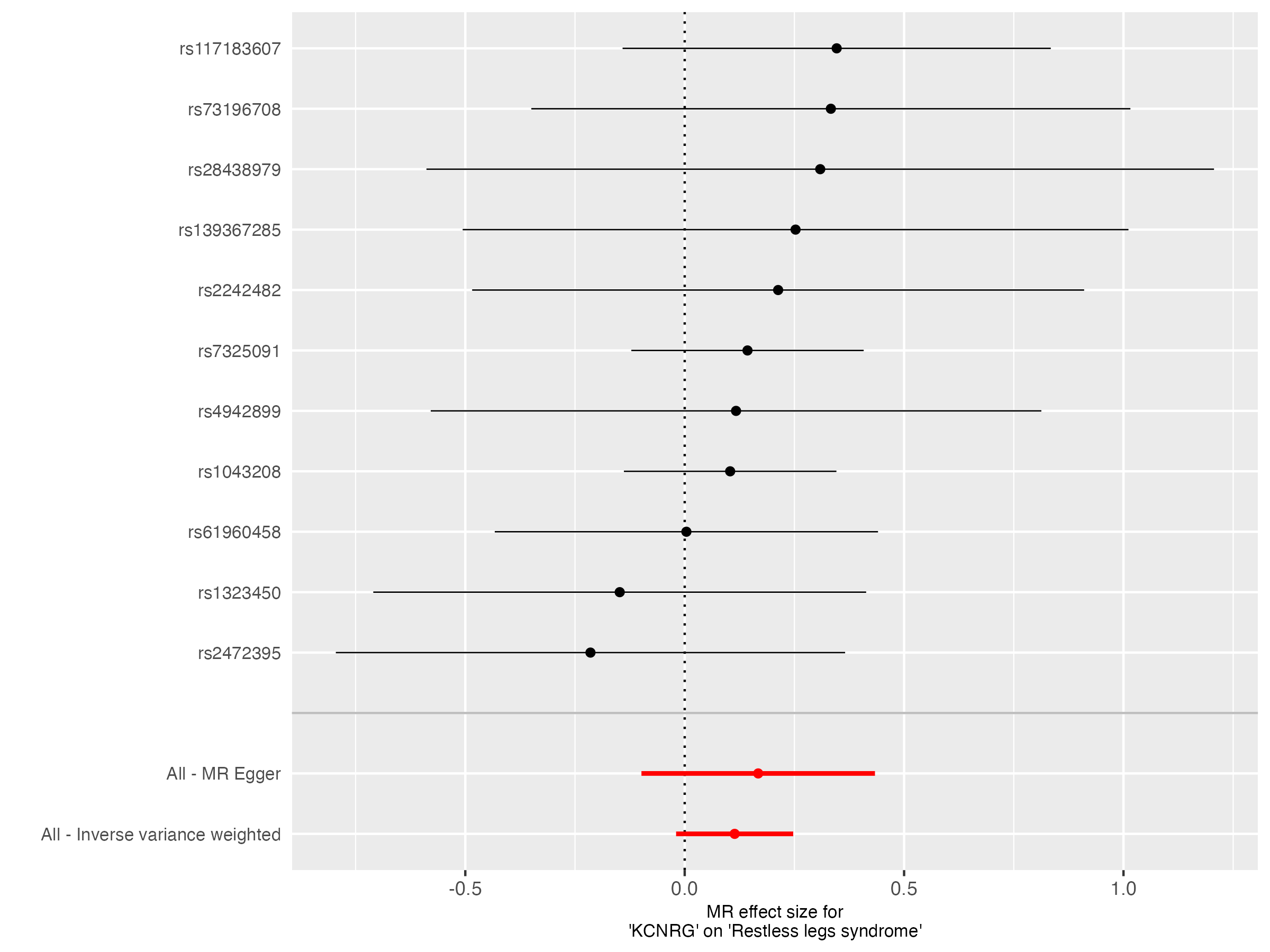

### forest_plot.png

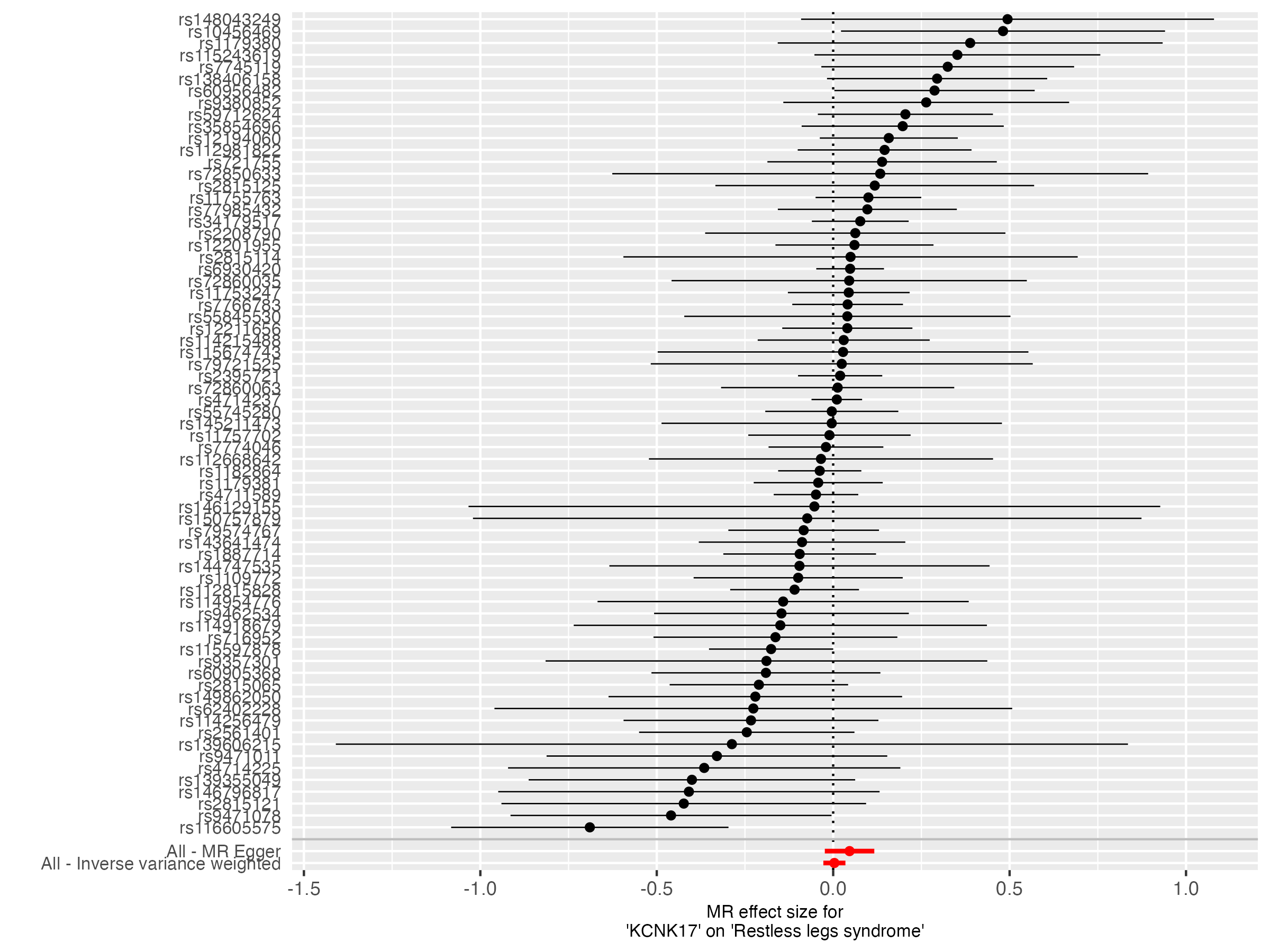

### forest_plot.png

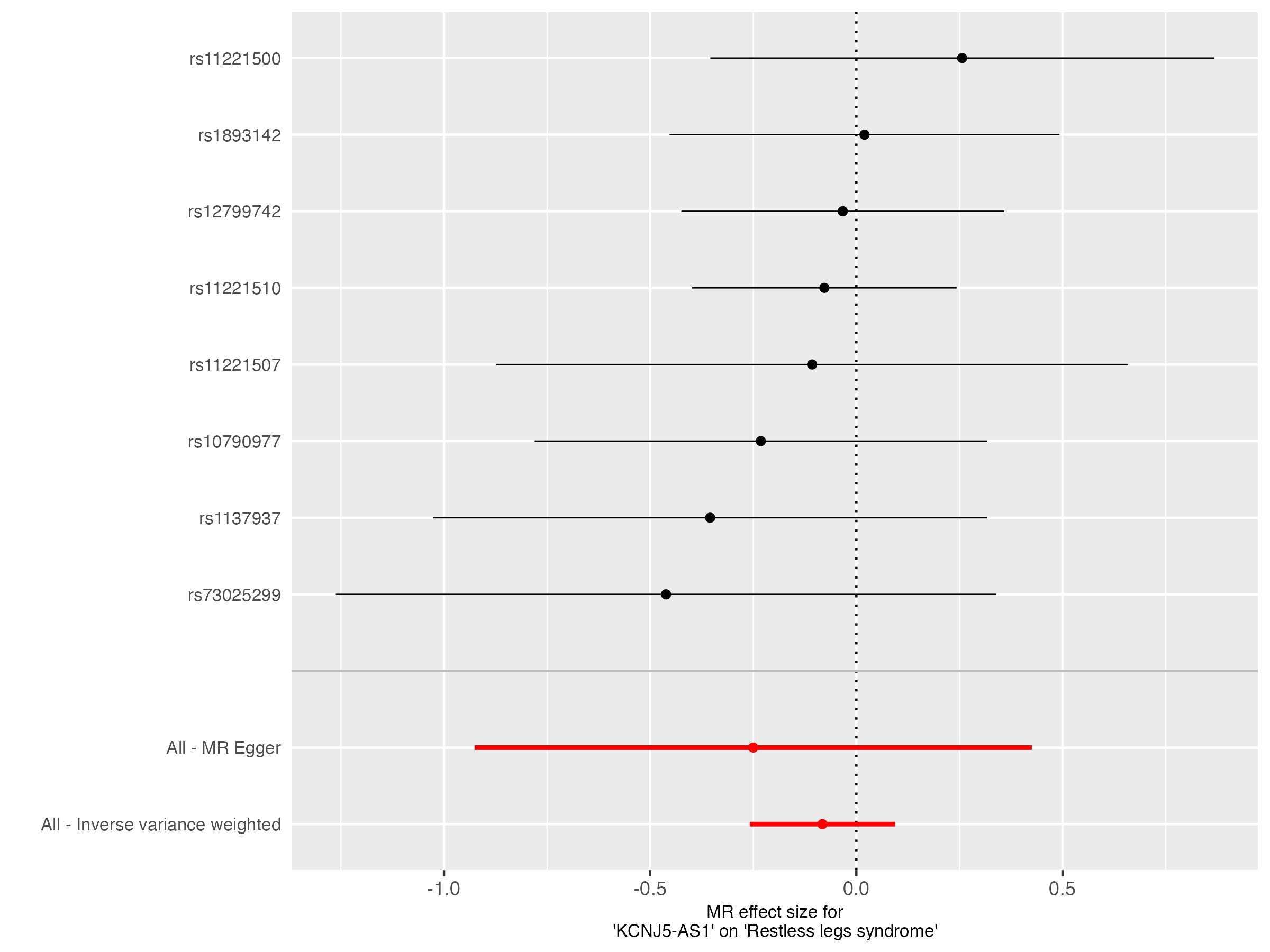

### forest_plot.png

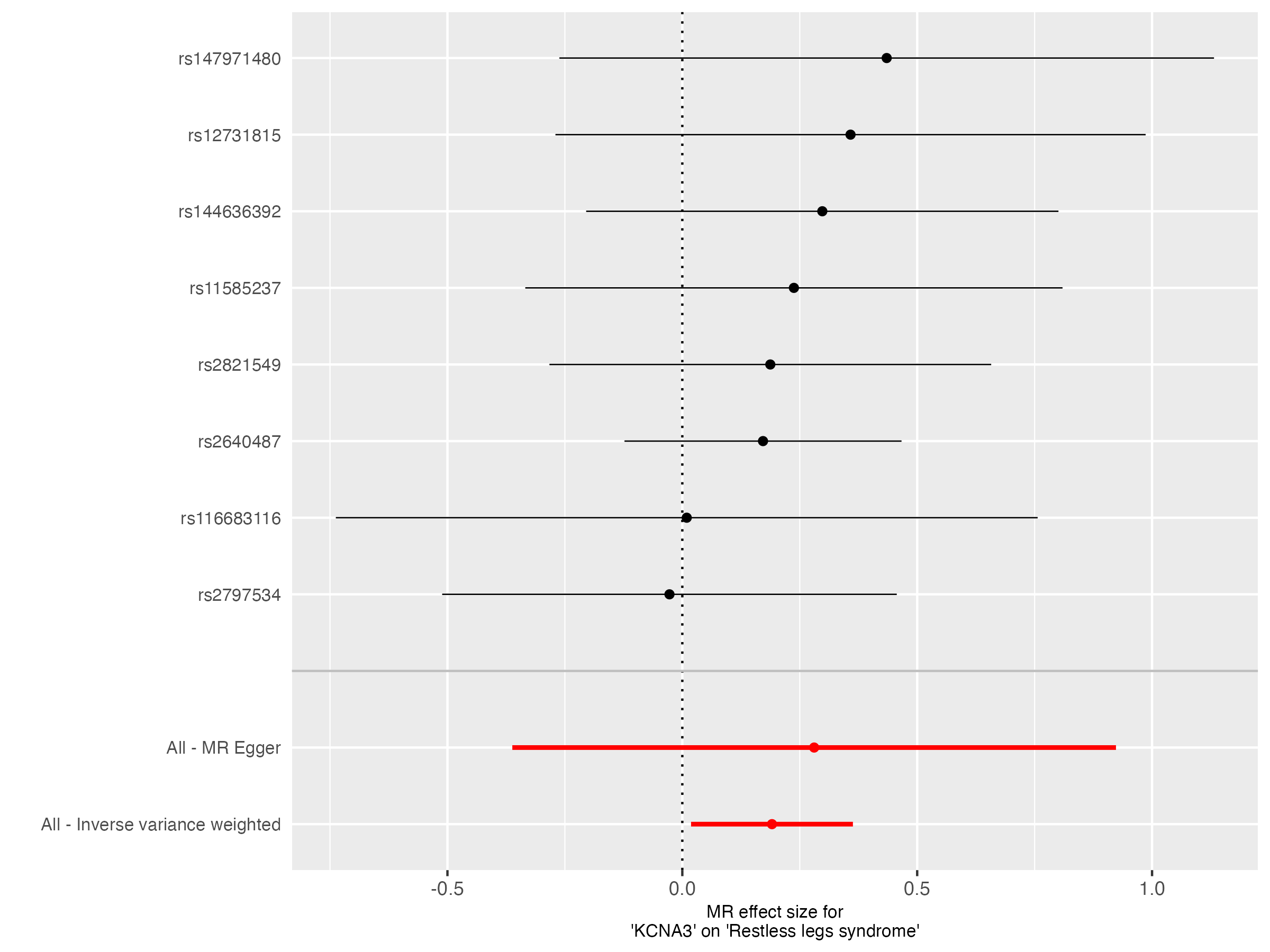

### forest_plot.png

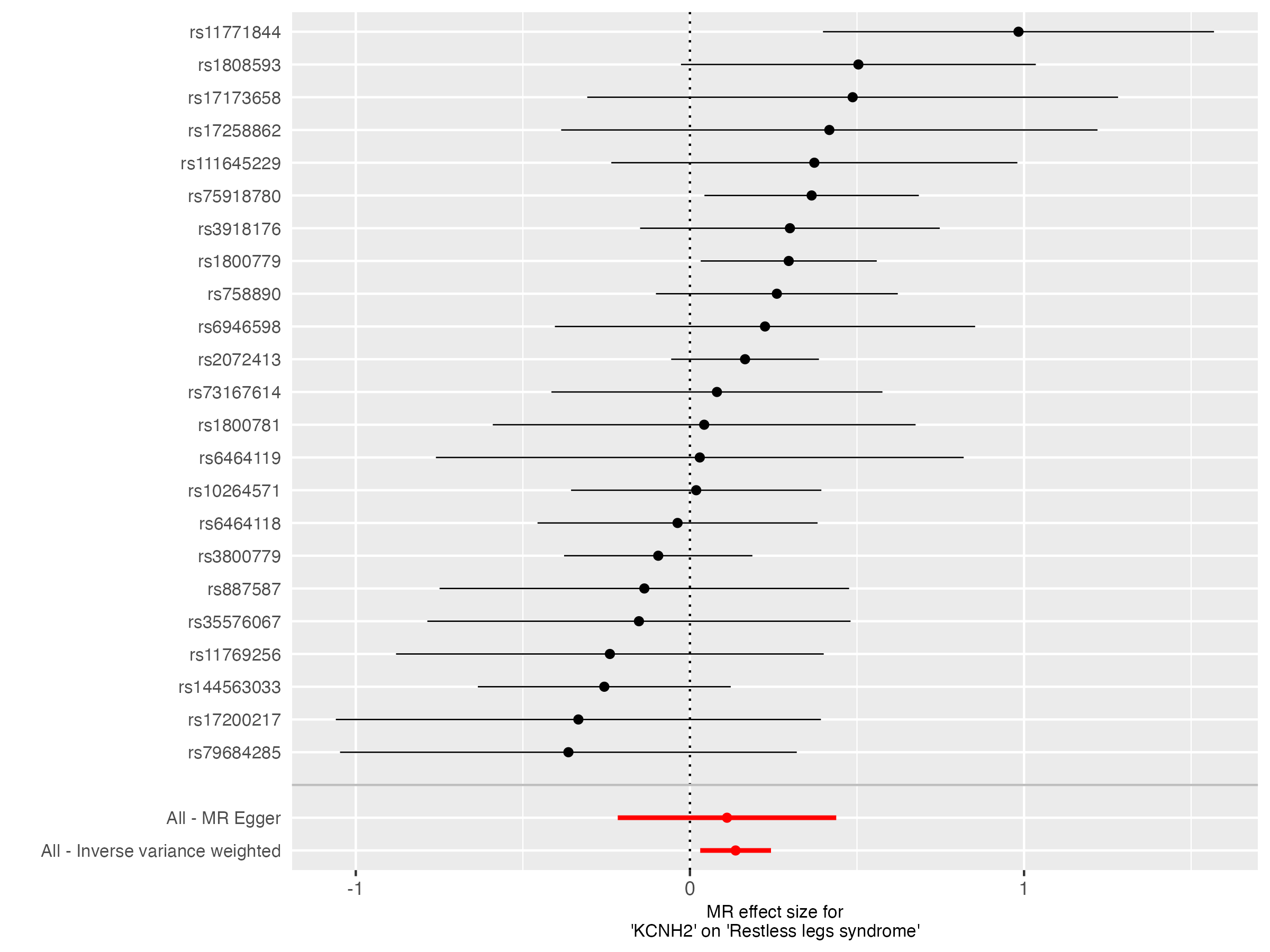

### forest_plot.png

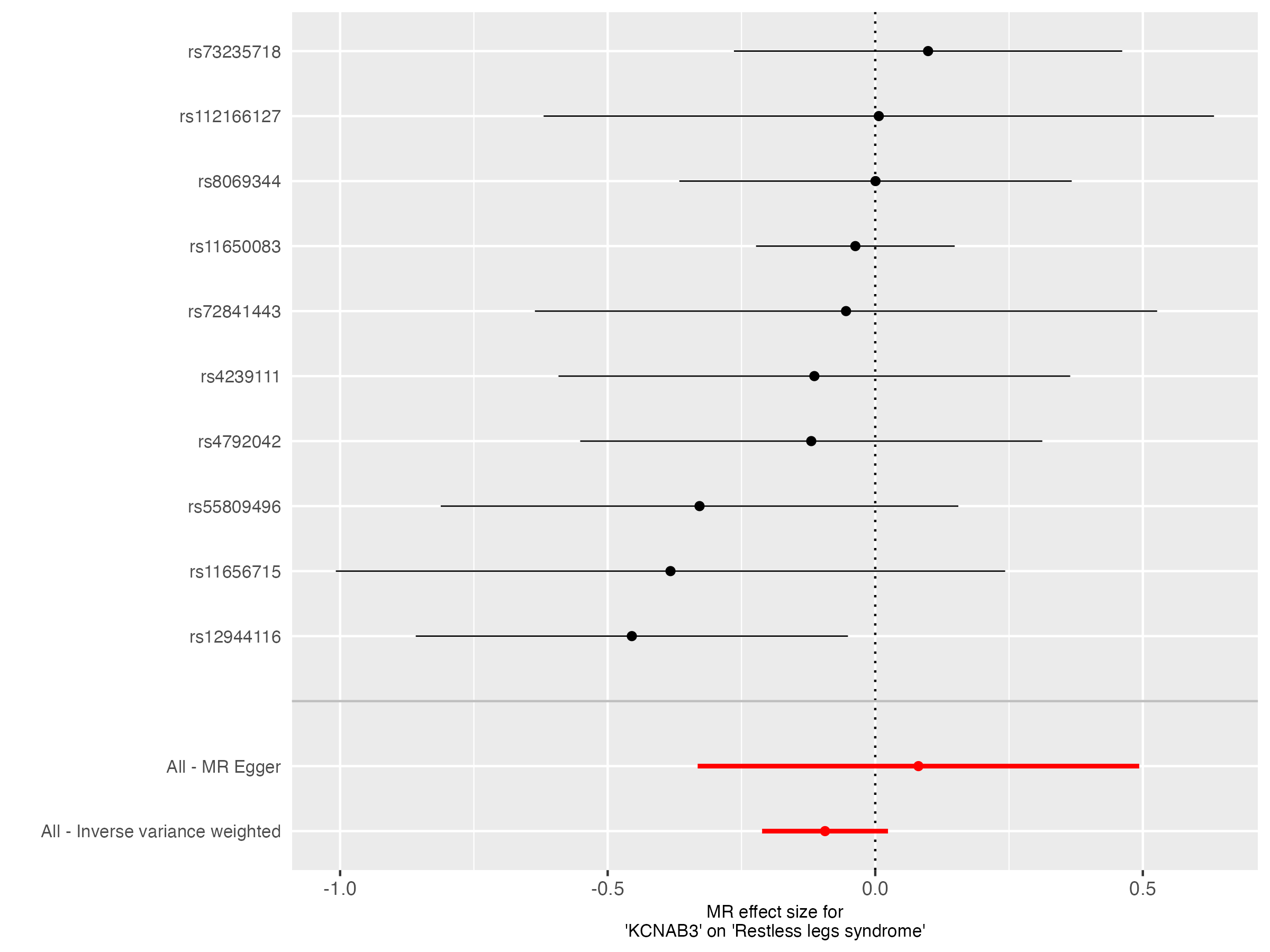

### forest_plot.png

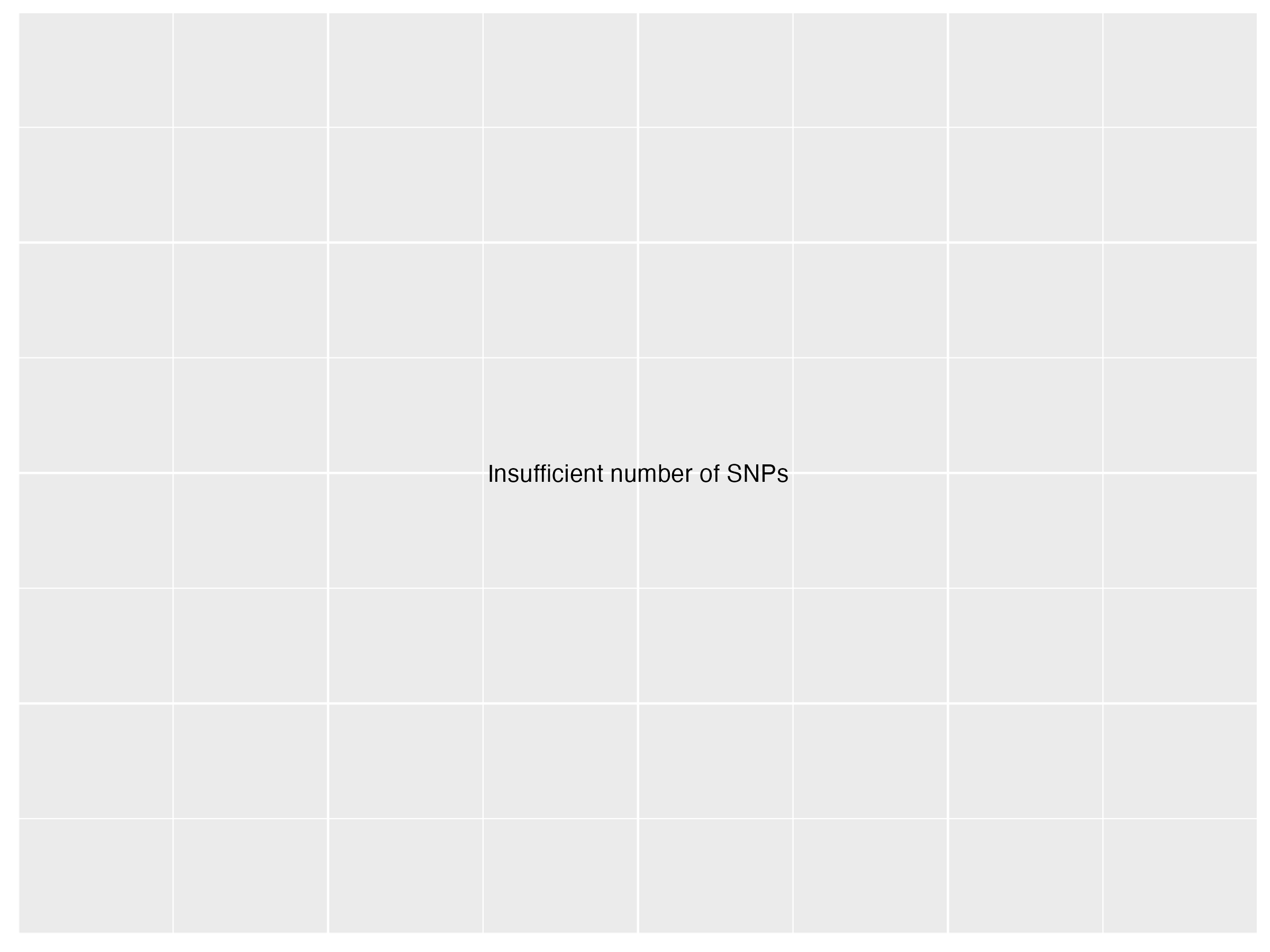

### forest_plot.png

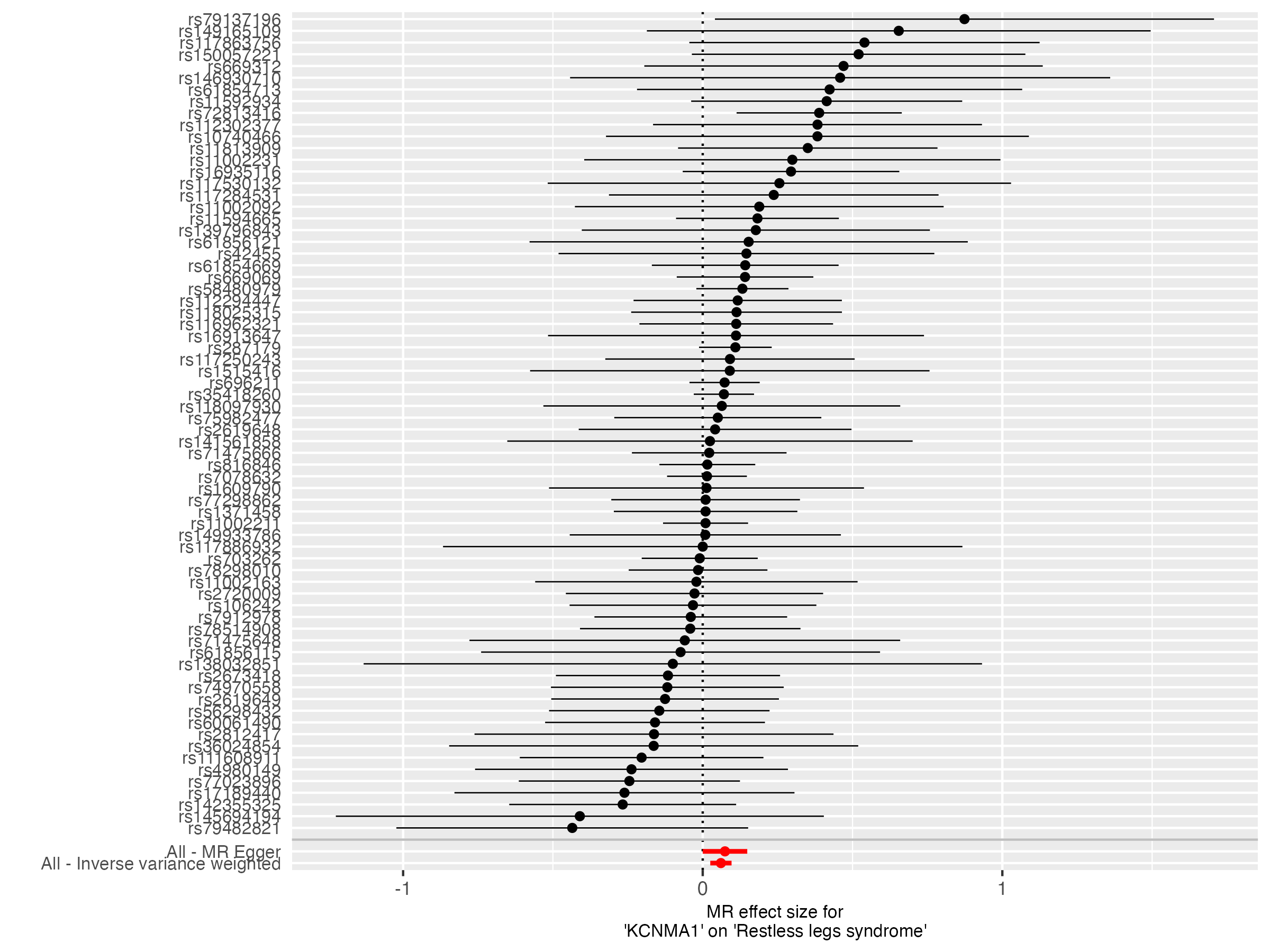

### forest_plot.png

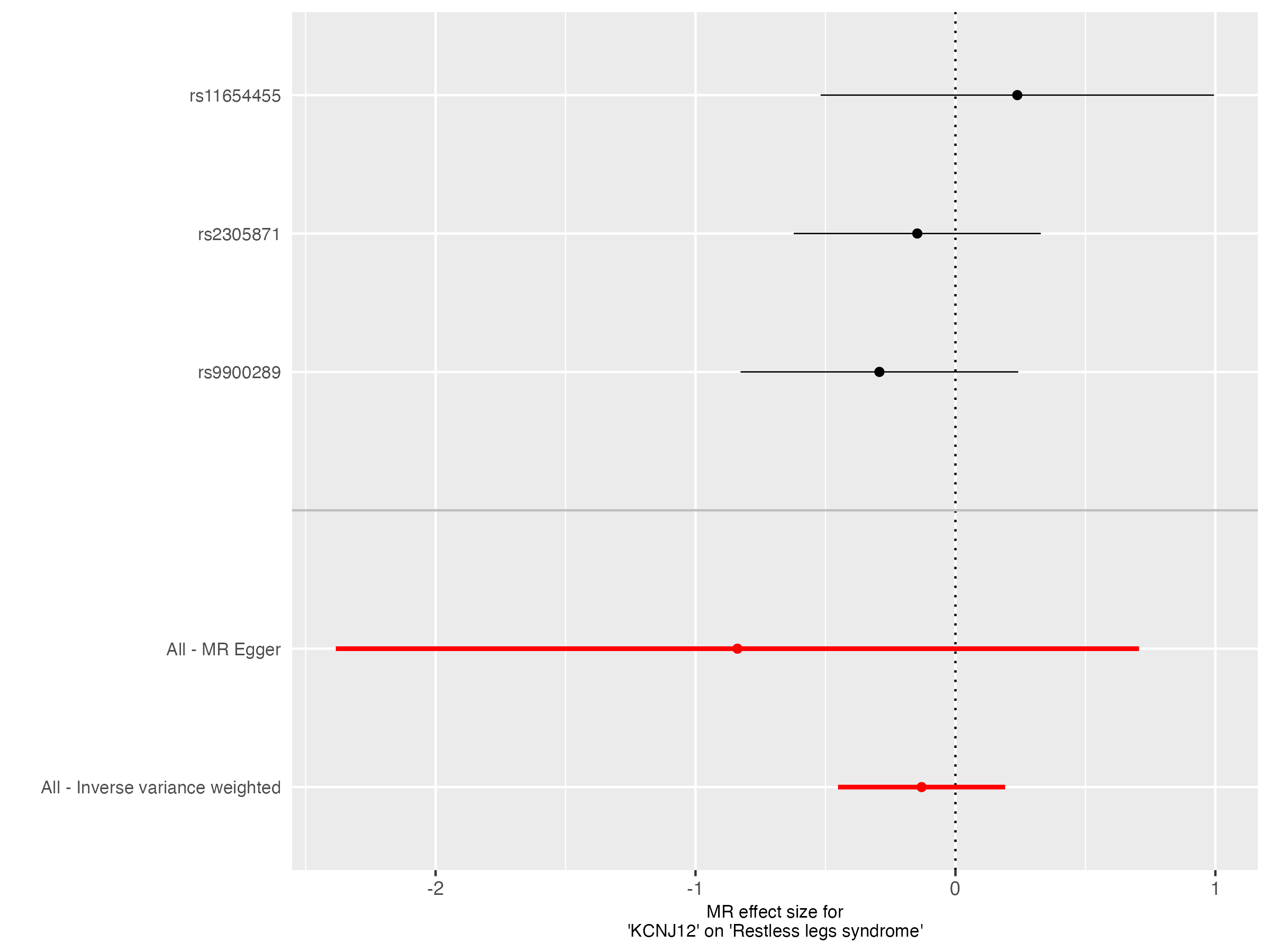

### forest_plot.png

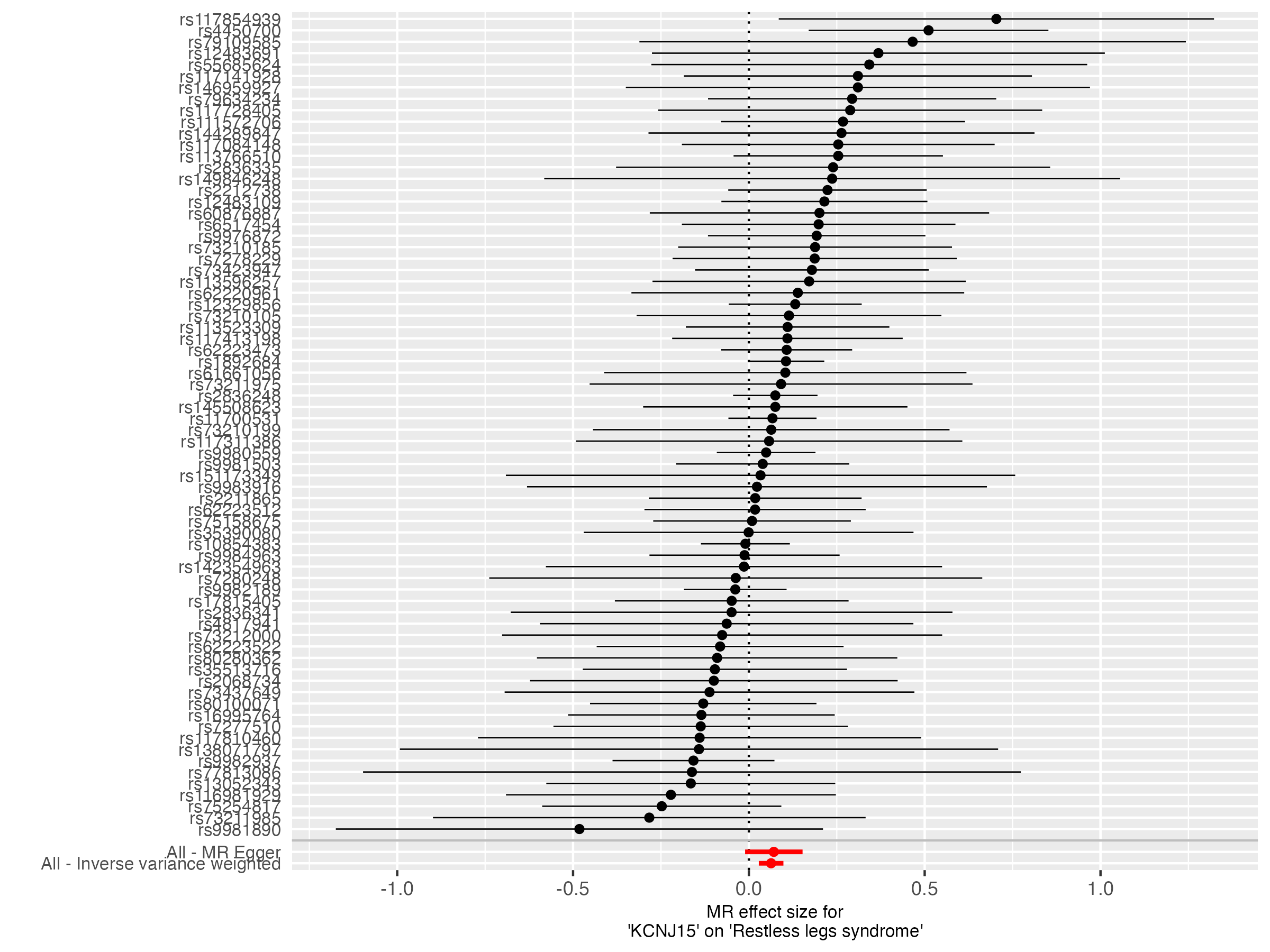

### forest_plot.png

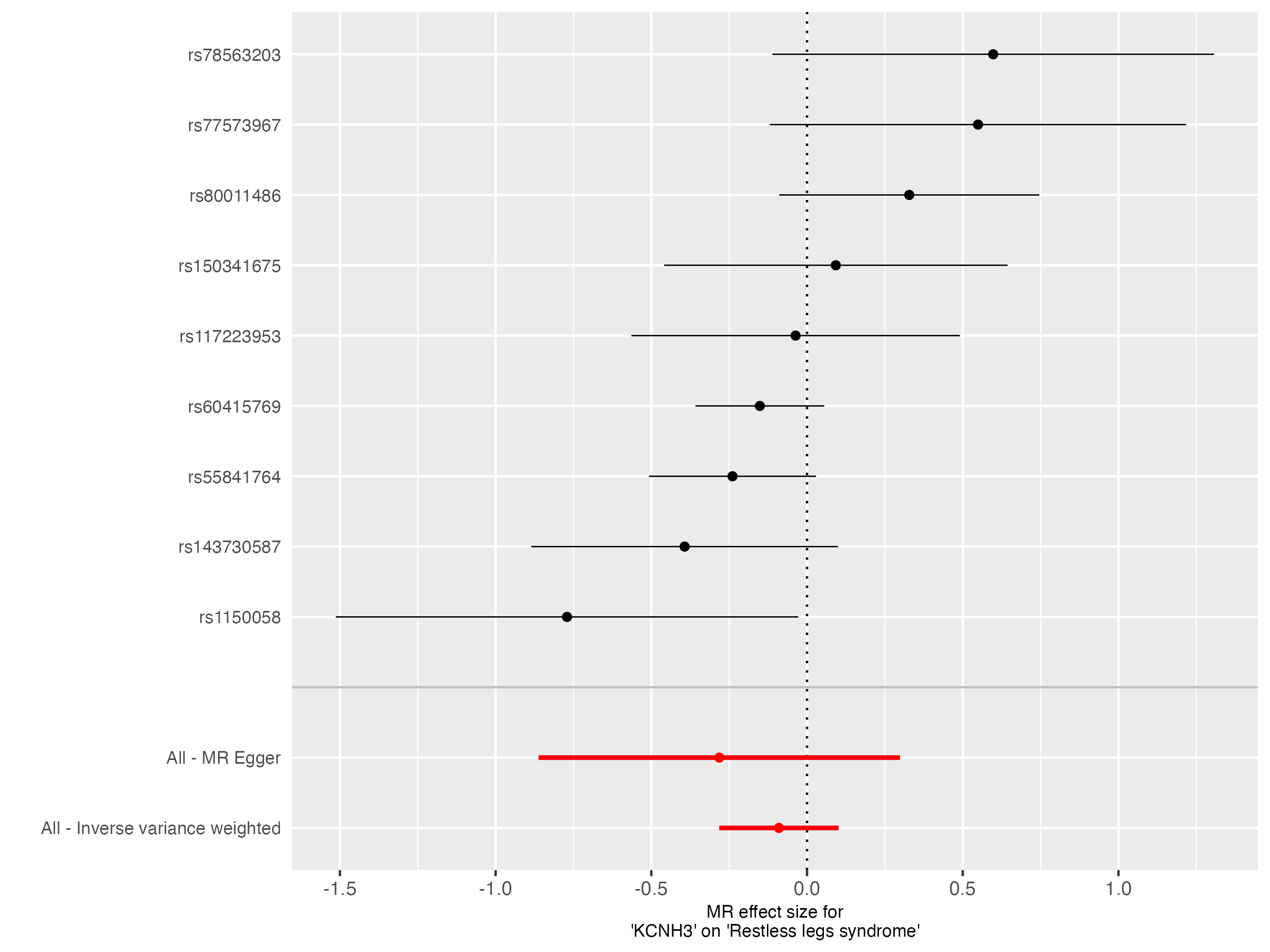

### funnel_plot.png

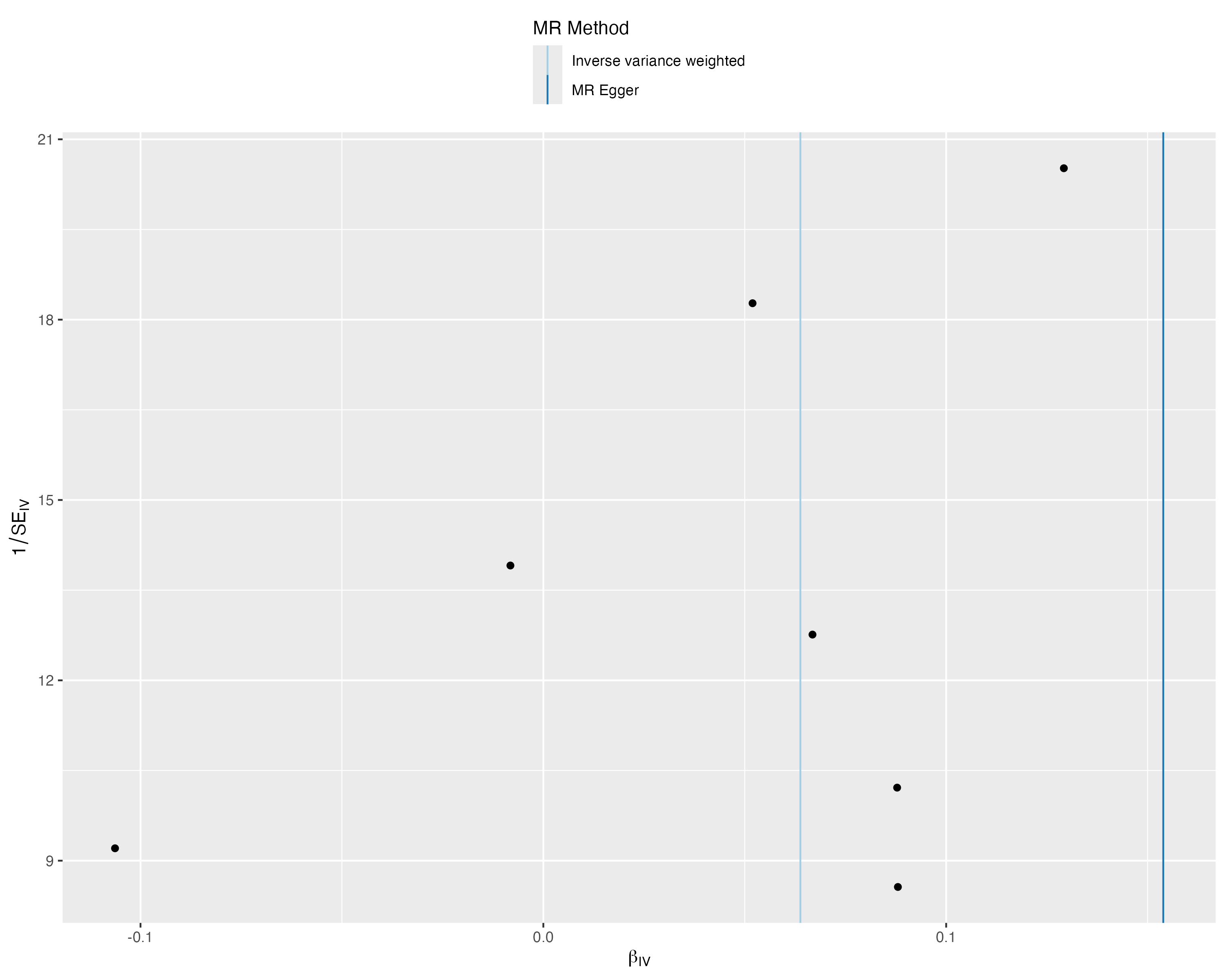

### funnel_plot_1.png

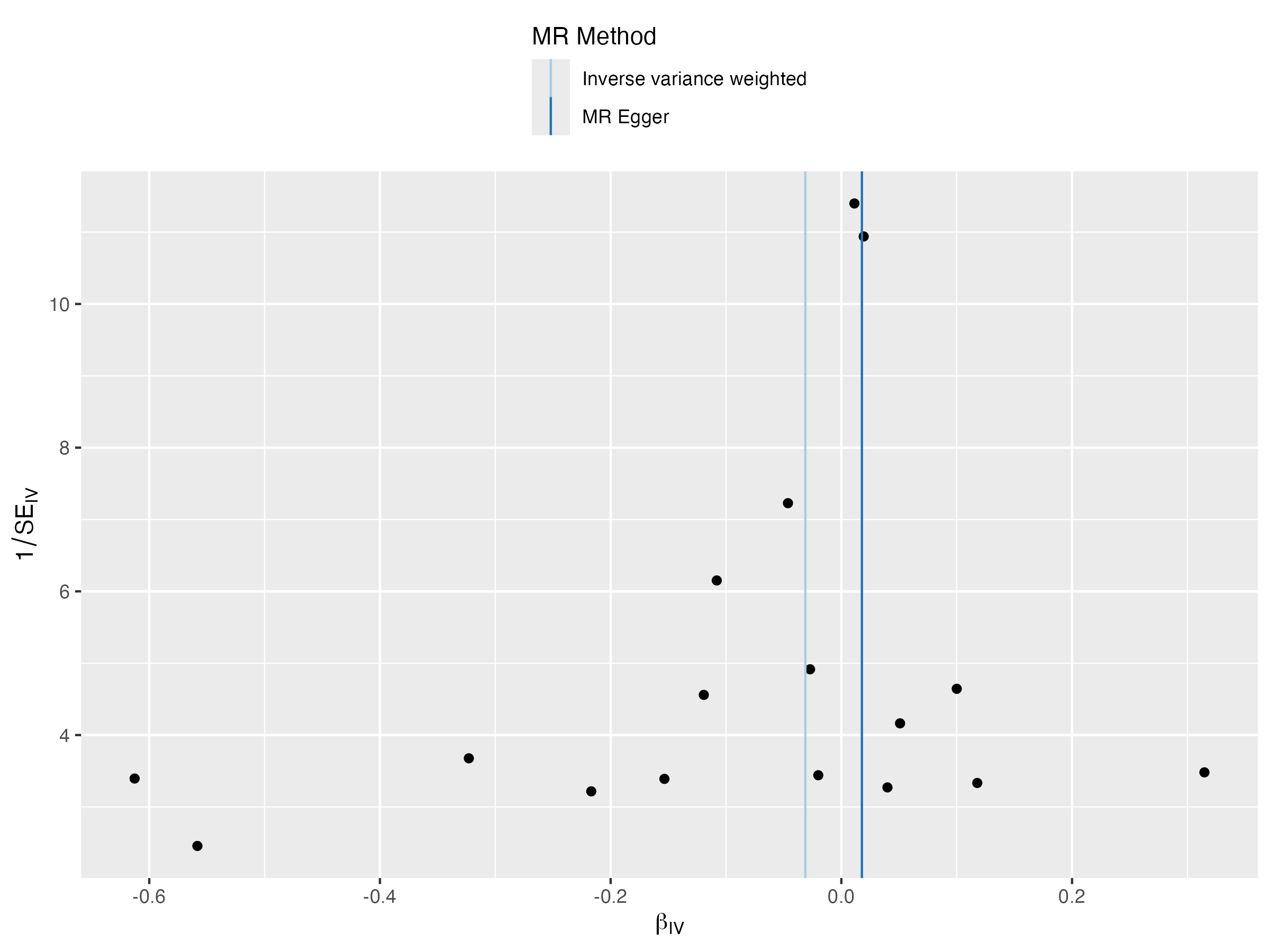

### funnel_plot_1.png

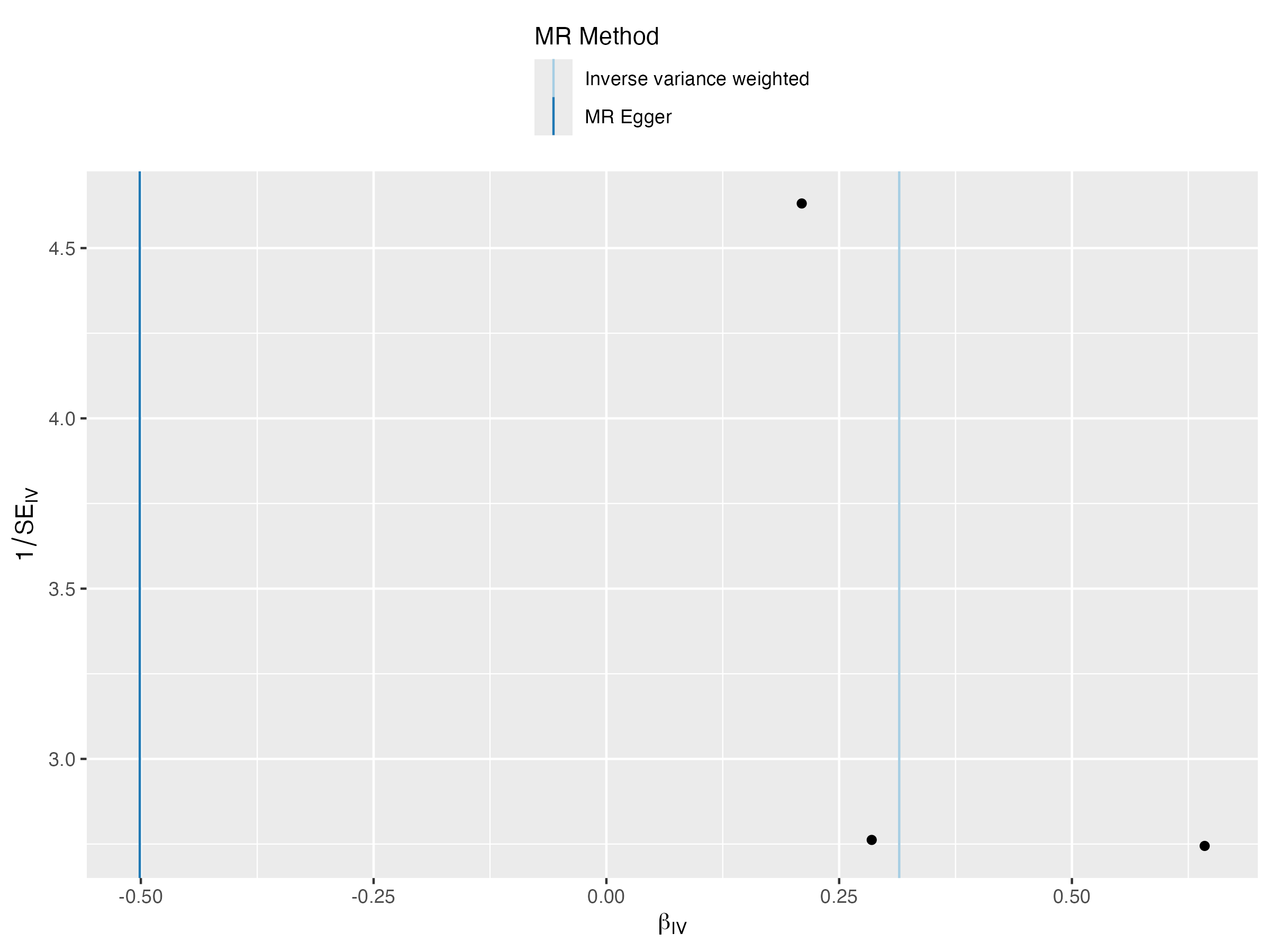

### funnel_plot_1.png

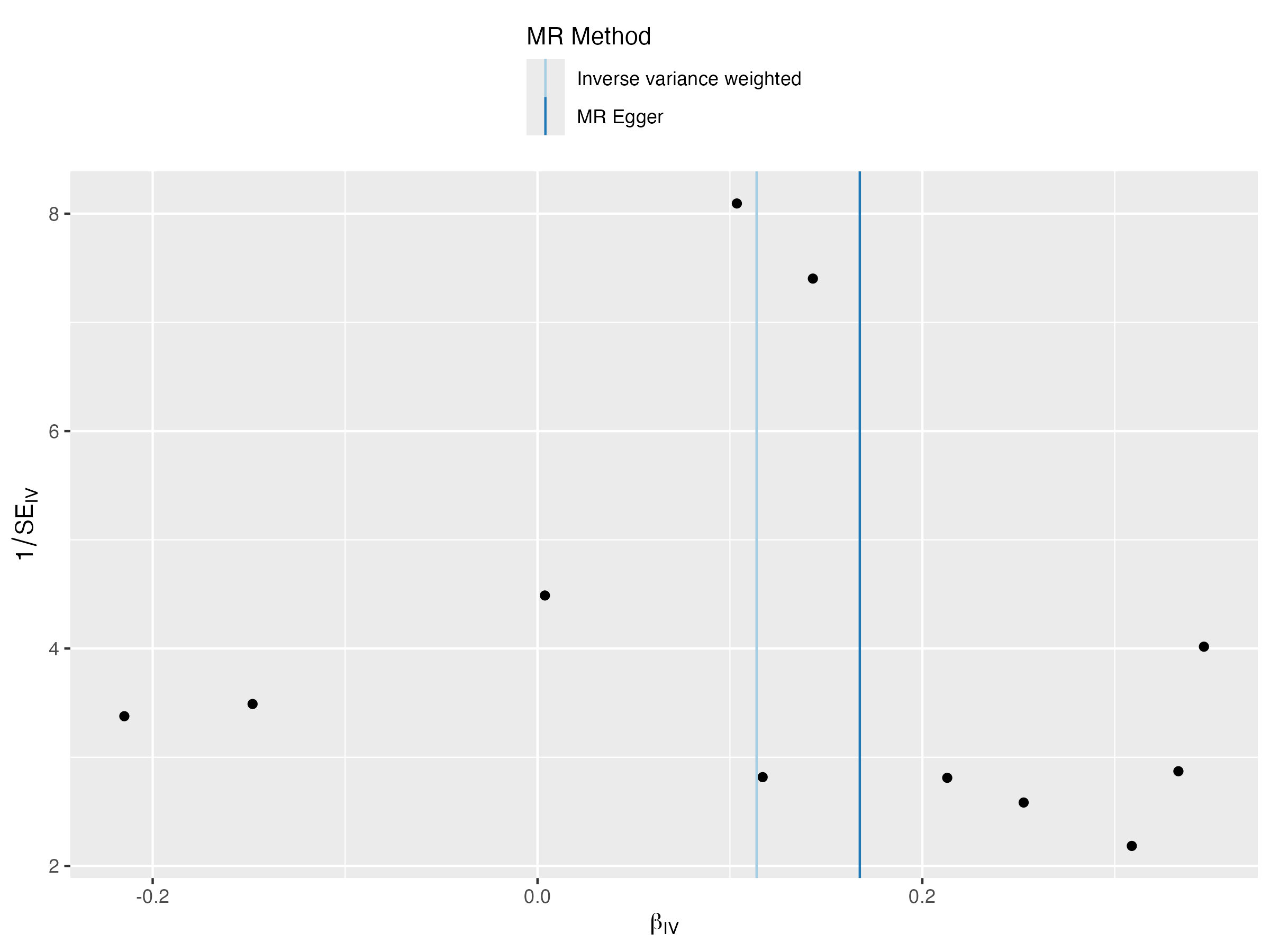

### funnel_plot_1.png

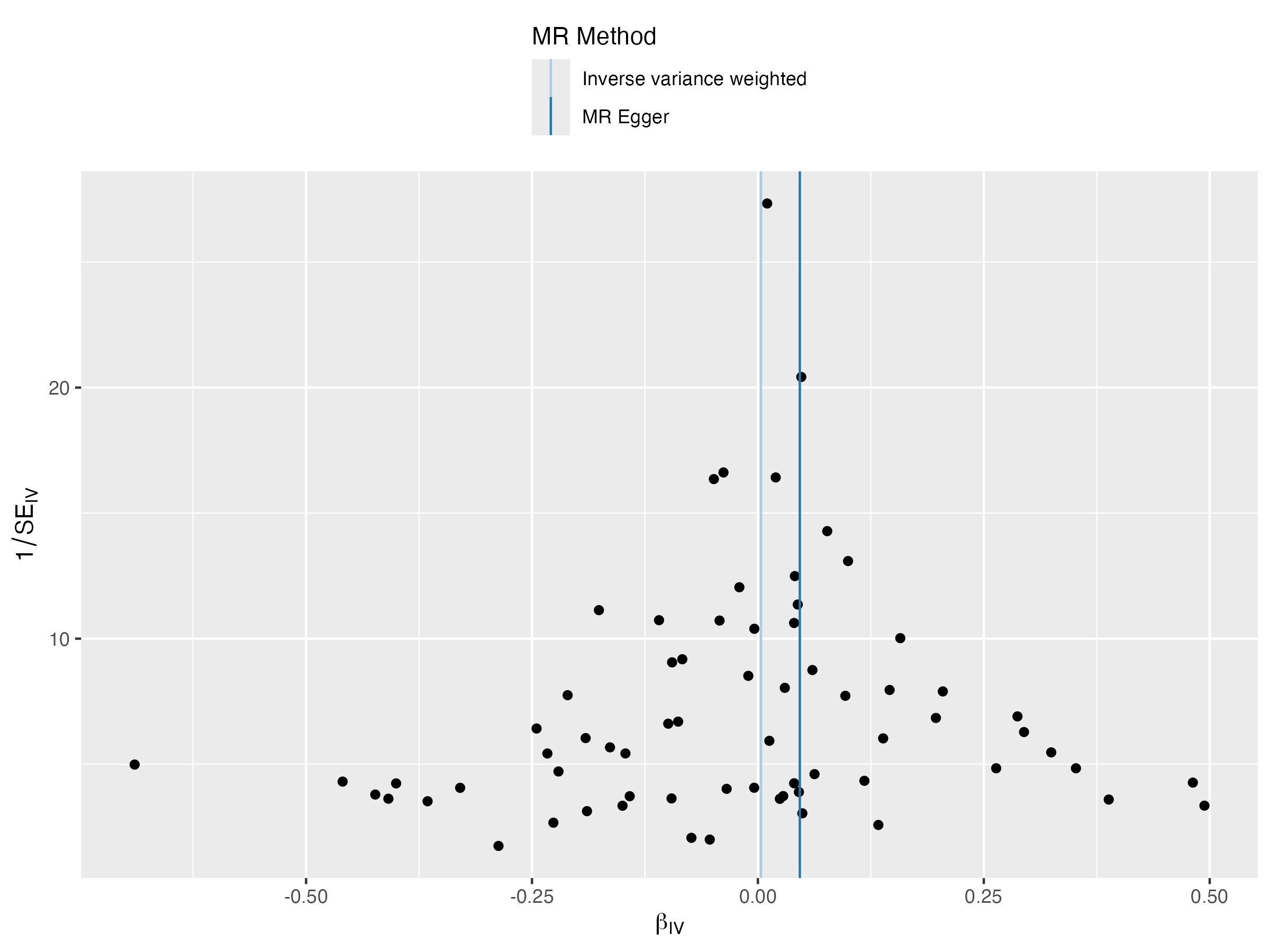

### funnel_plot_1.png

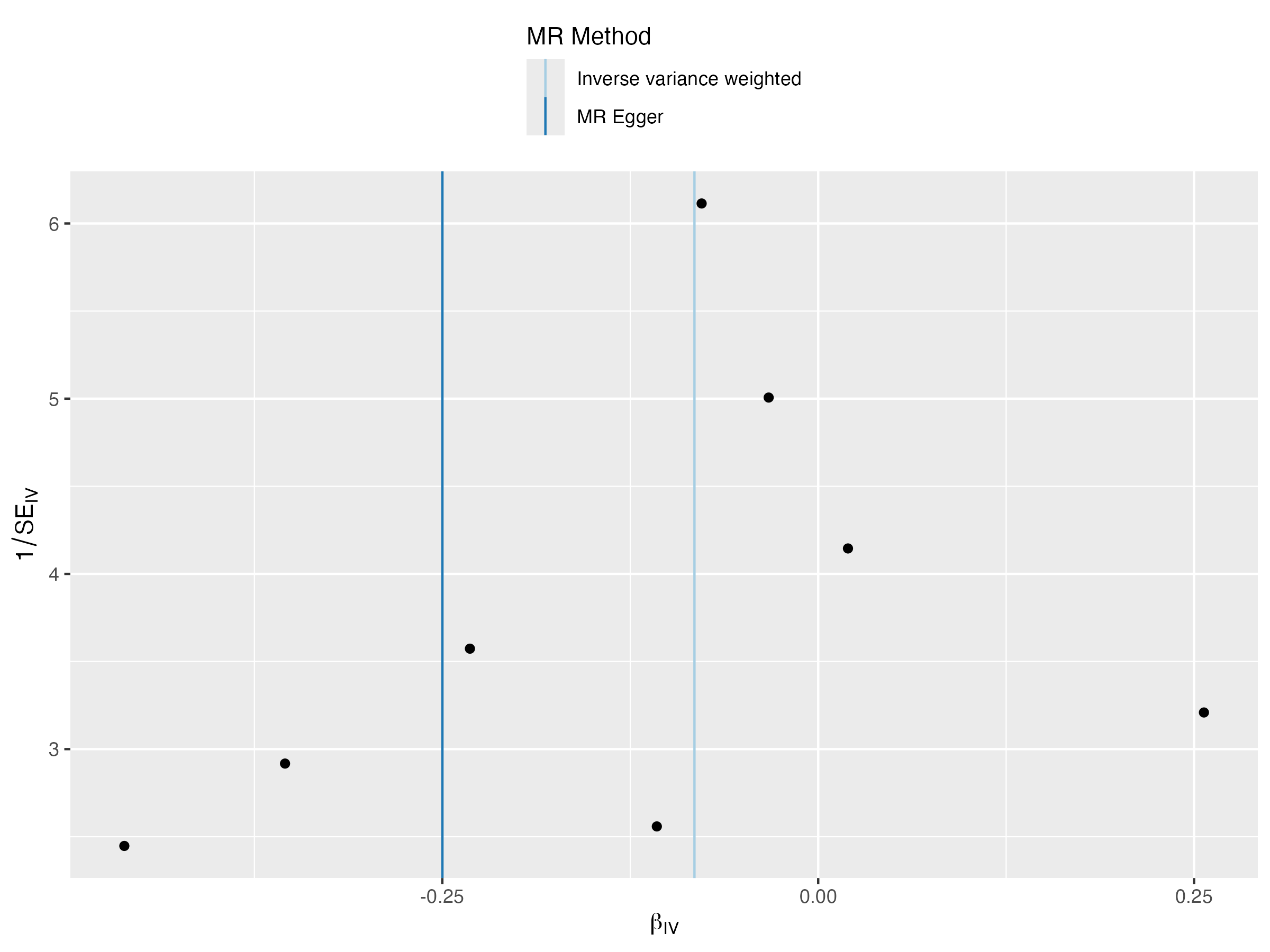

### funnel_plot_1.png

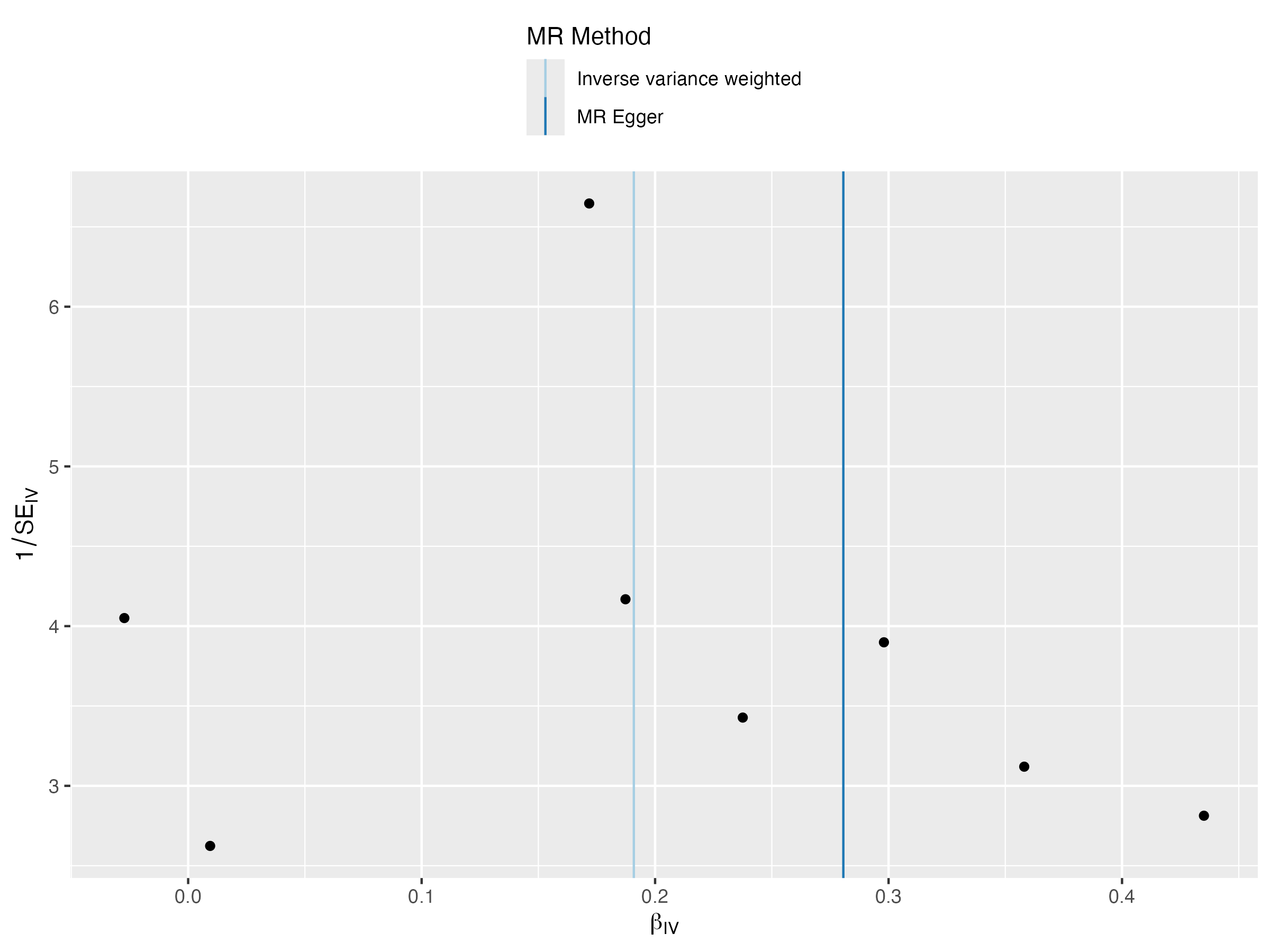

### funnel_plot_1.png

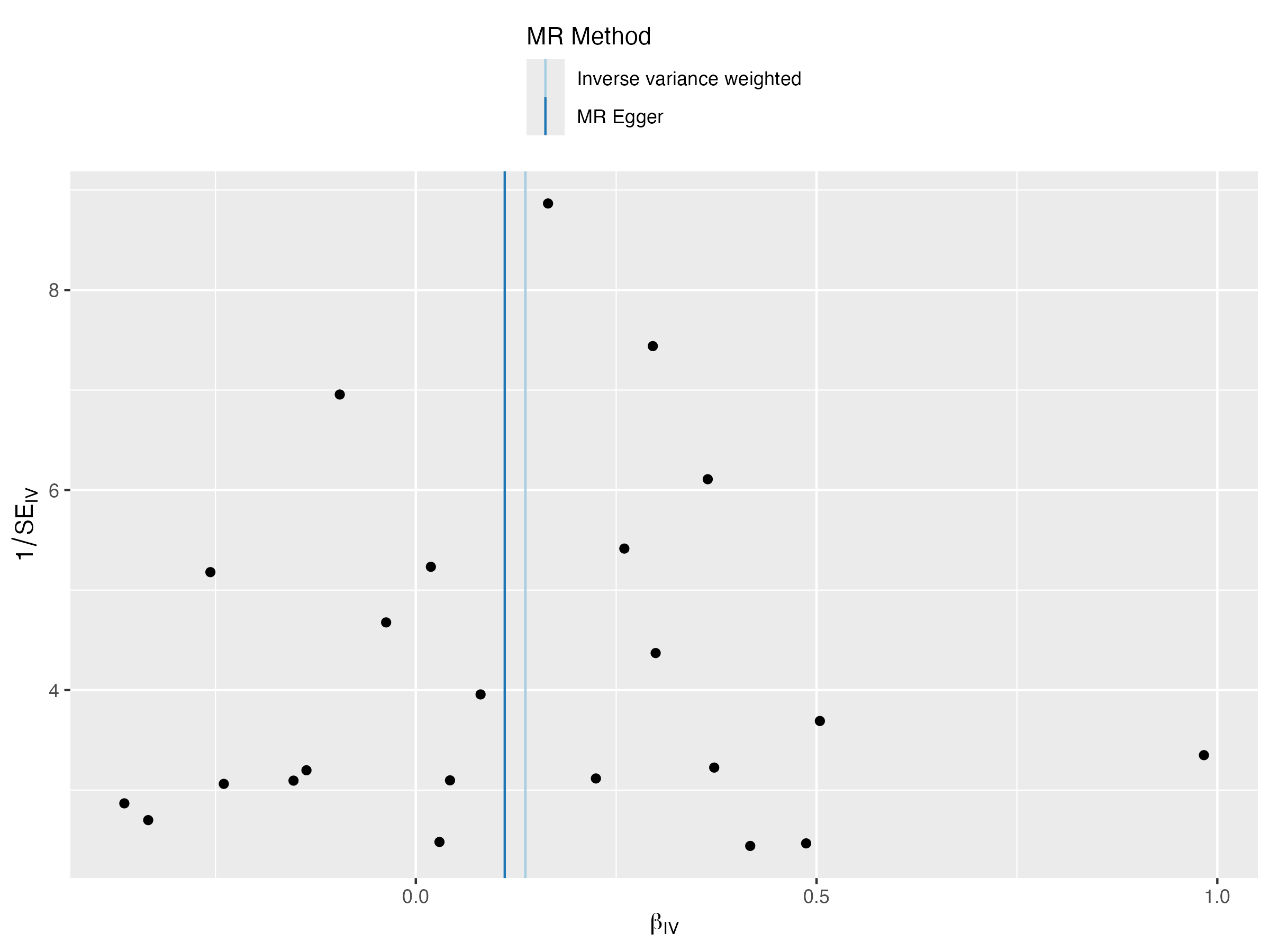

### funnel_plot_1.png

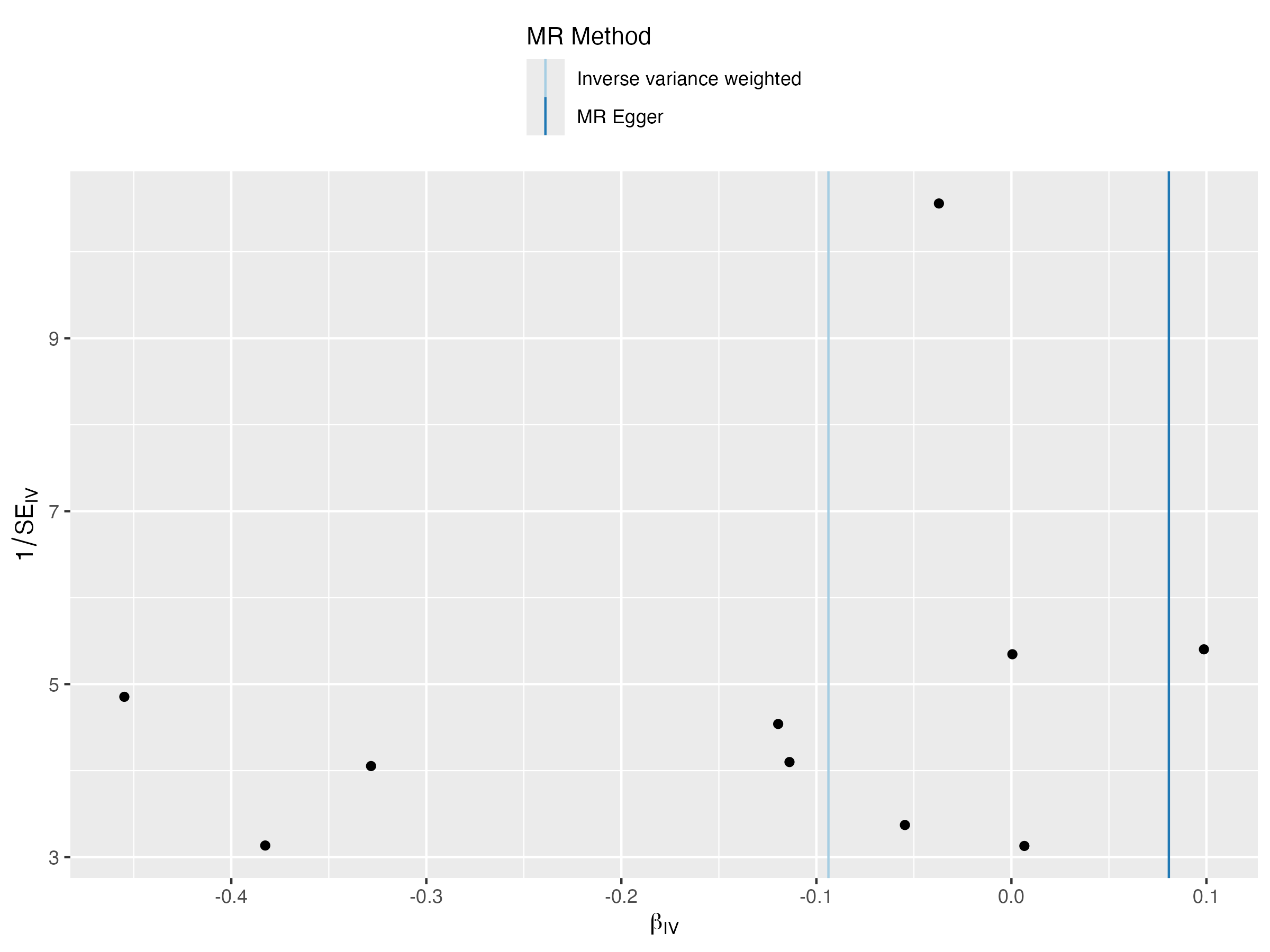

### funnel_plot_1.png

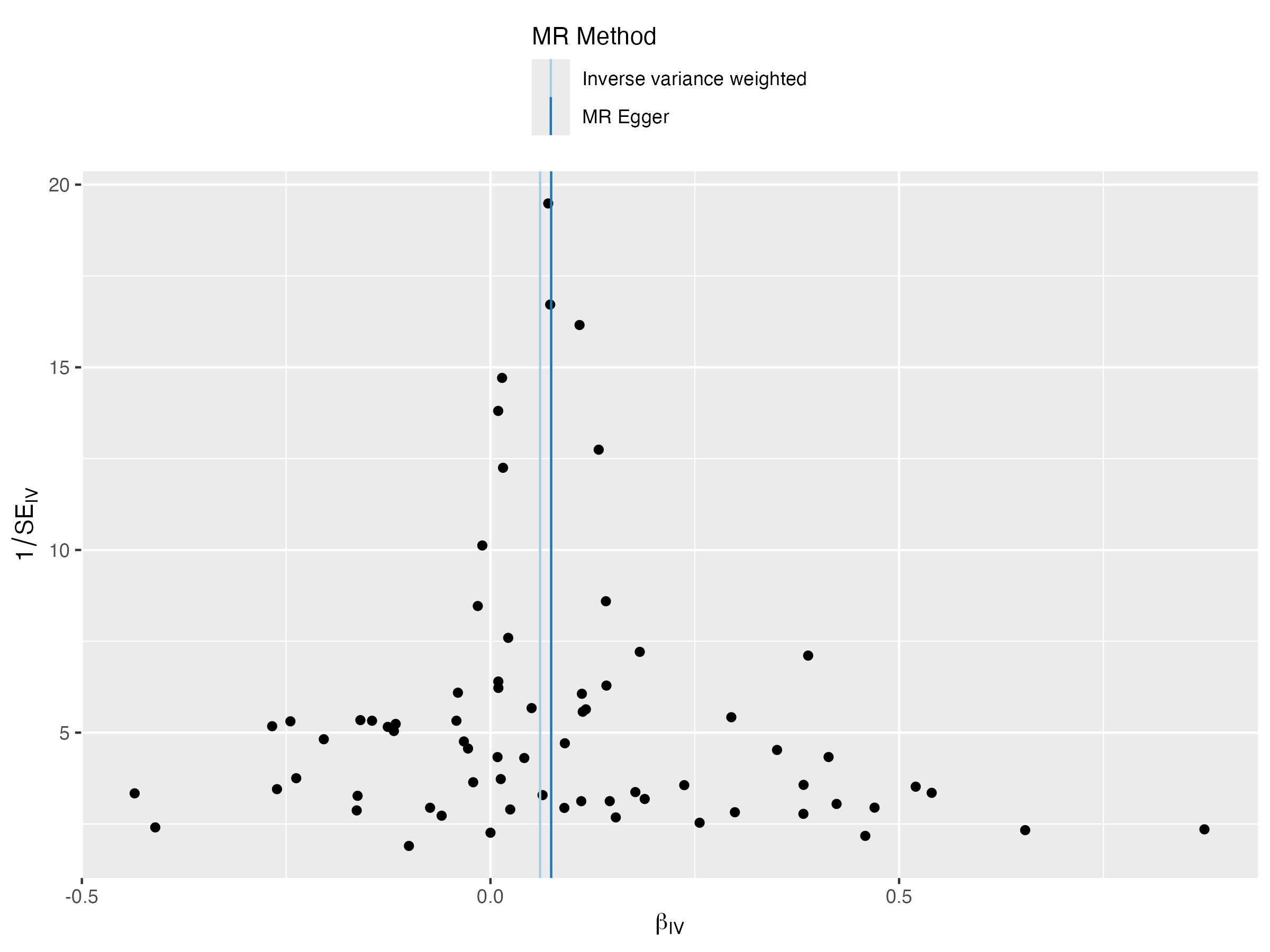

### funnel_plot_1.png

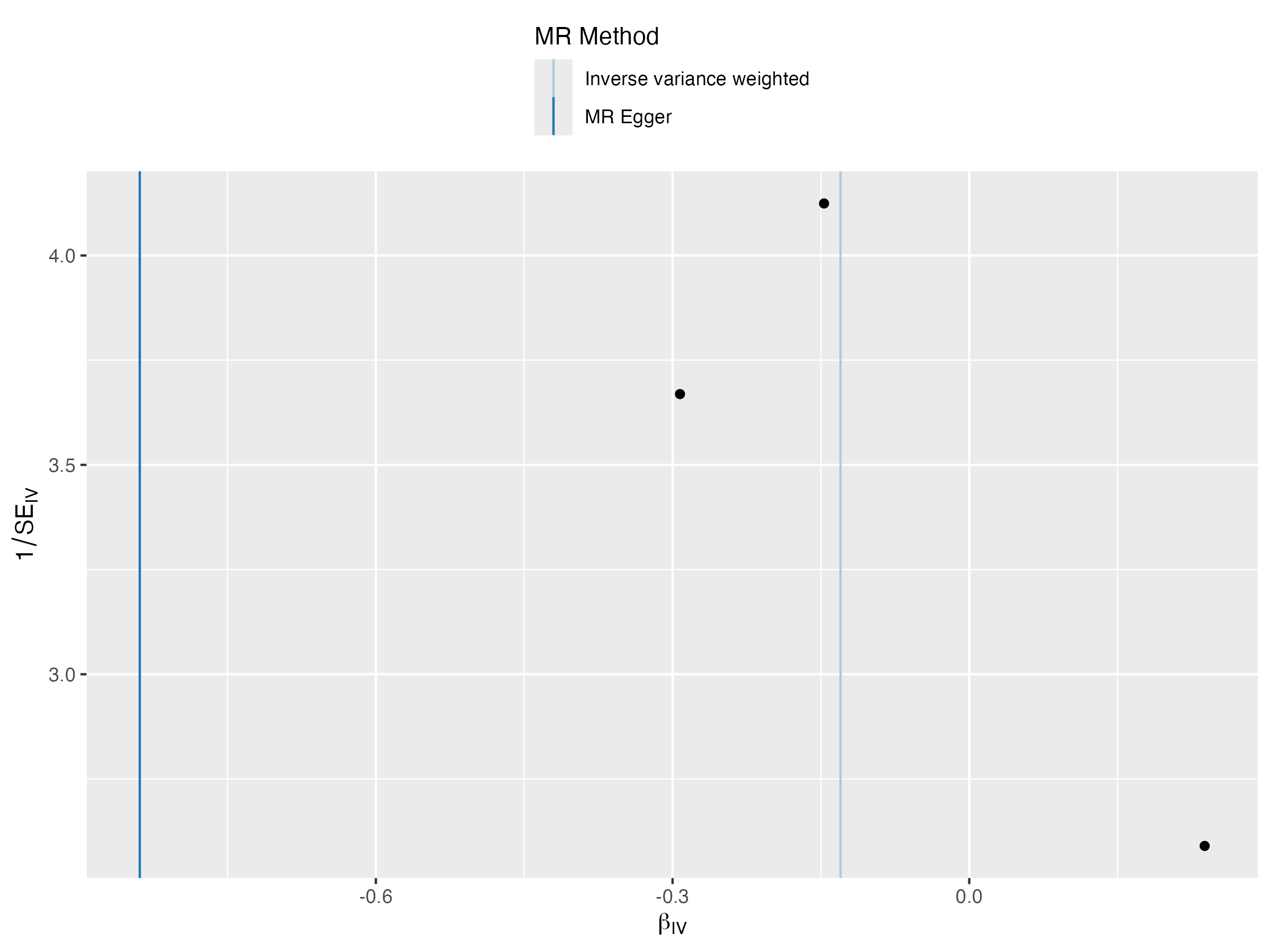

### funnel_plot_1.png

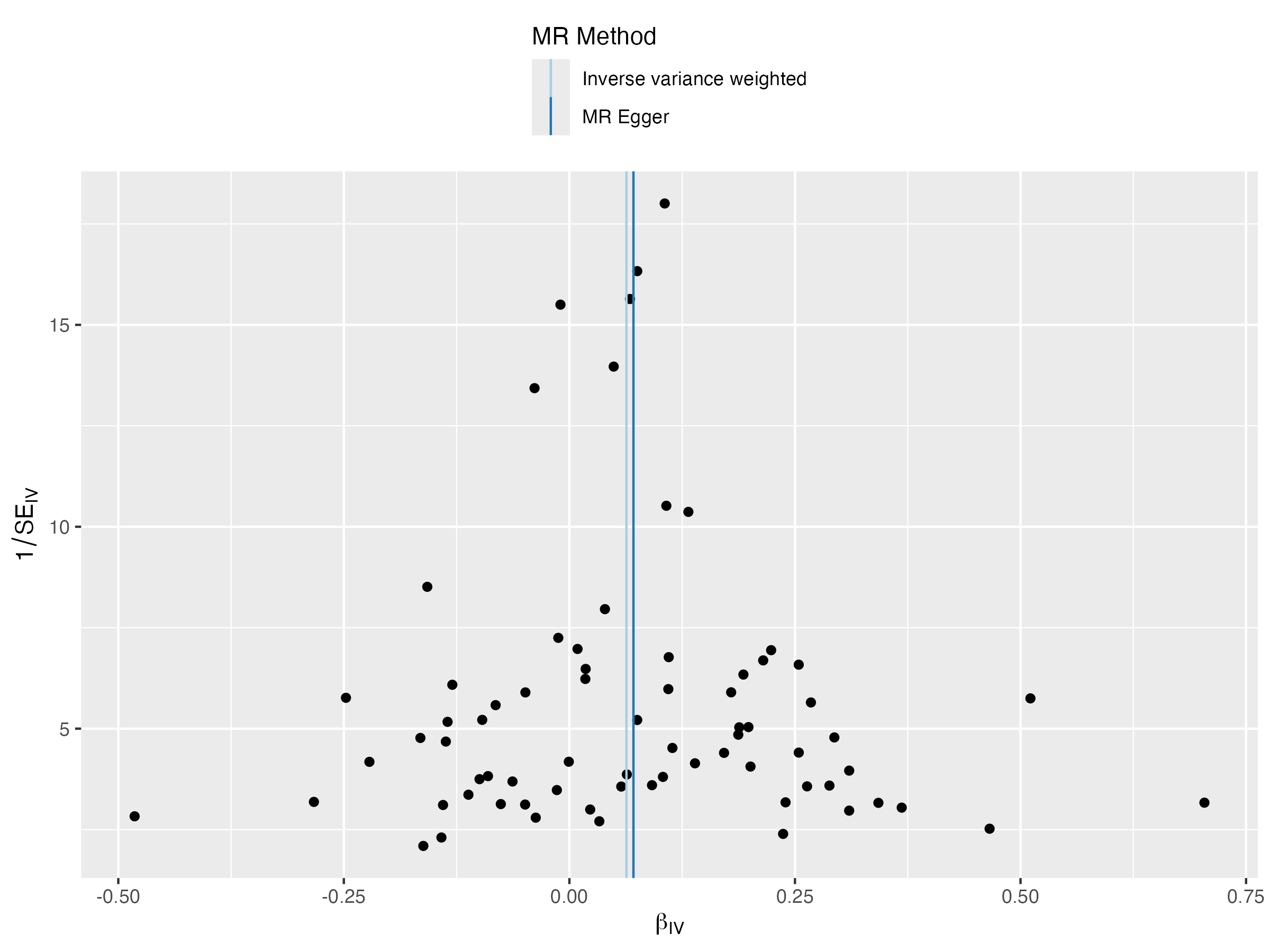

### leave_one_out_plot.png

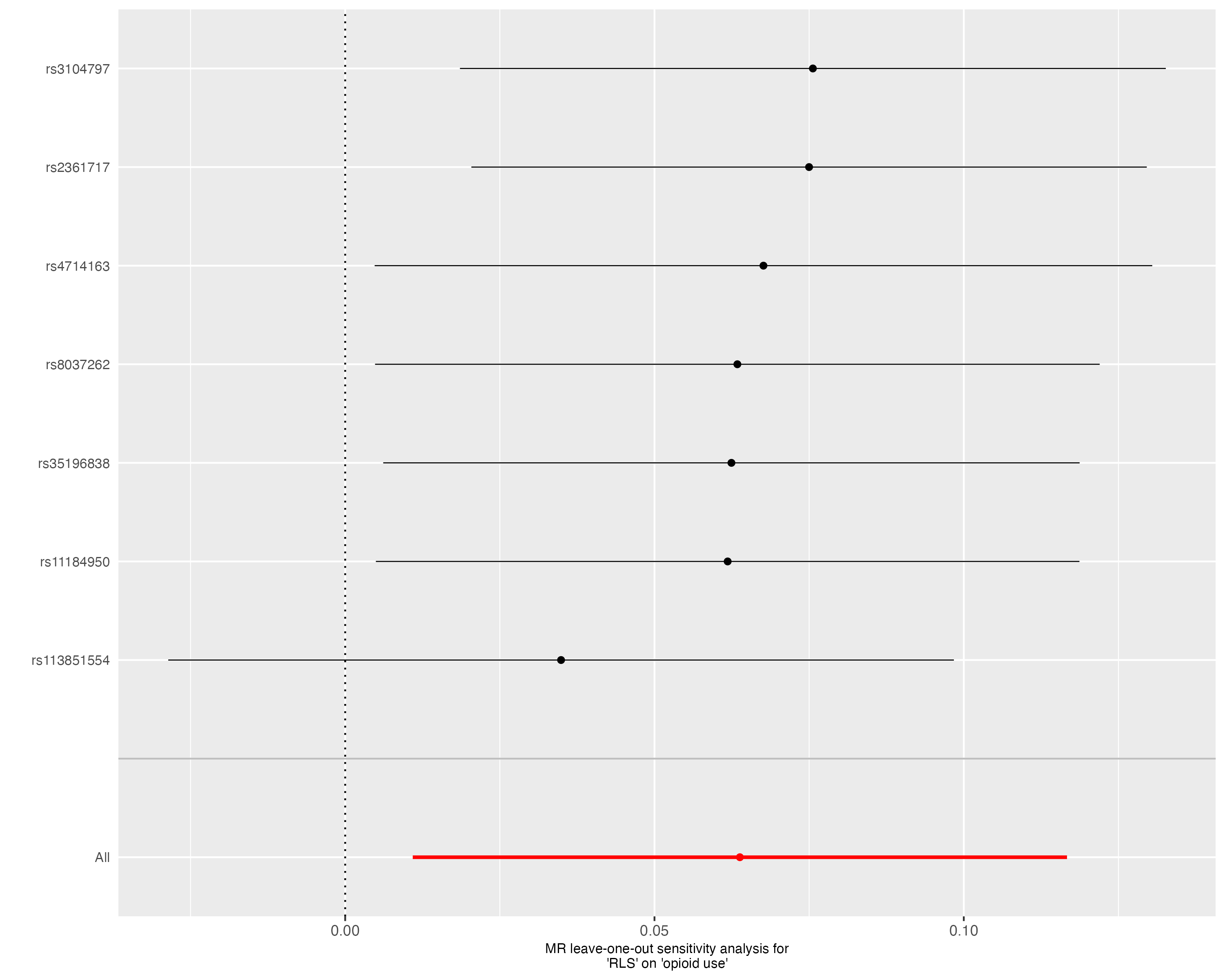

### leaveoneout_plot.png

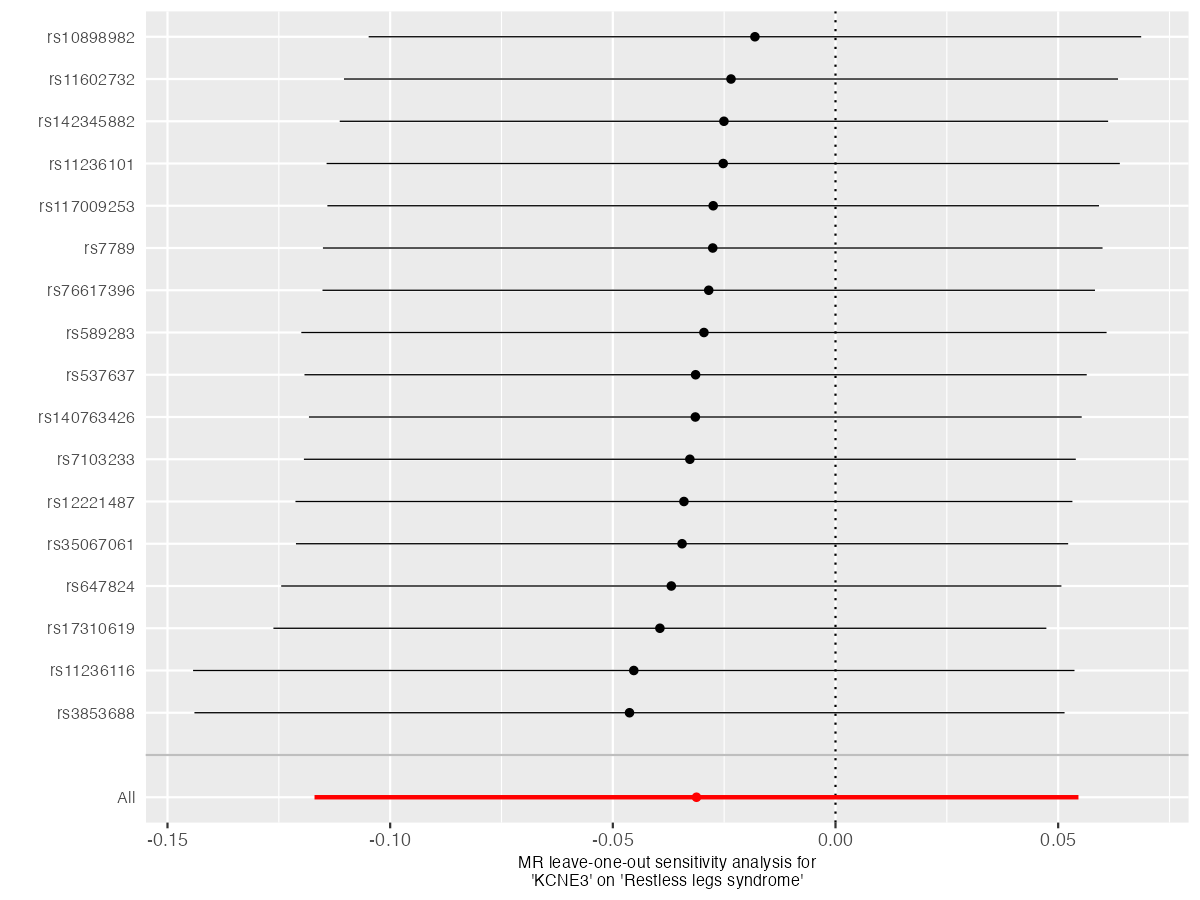

### leaveoneout_plot.png

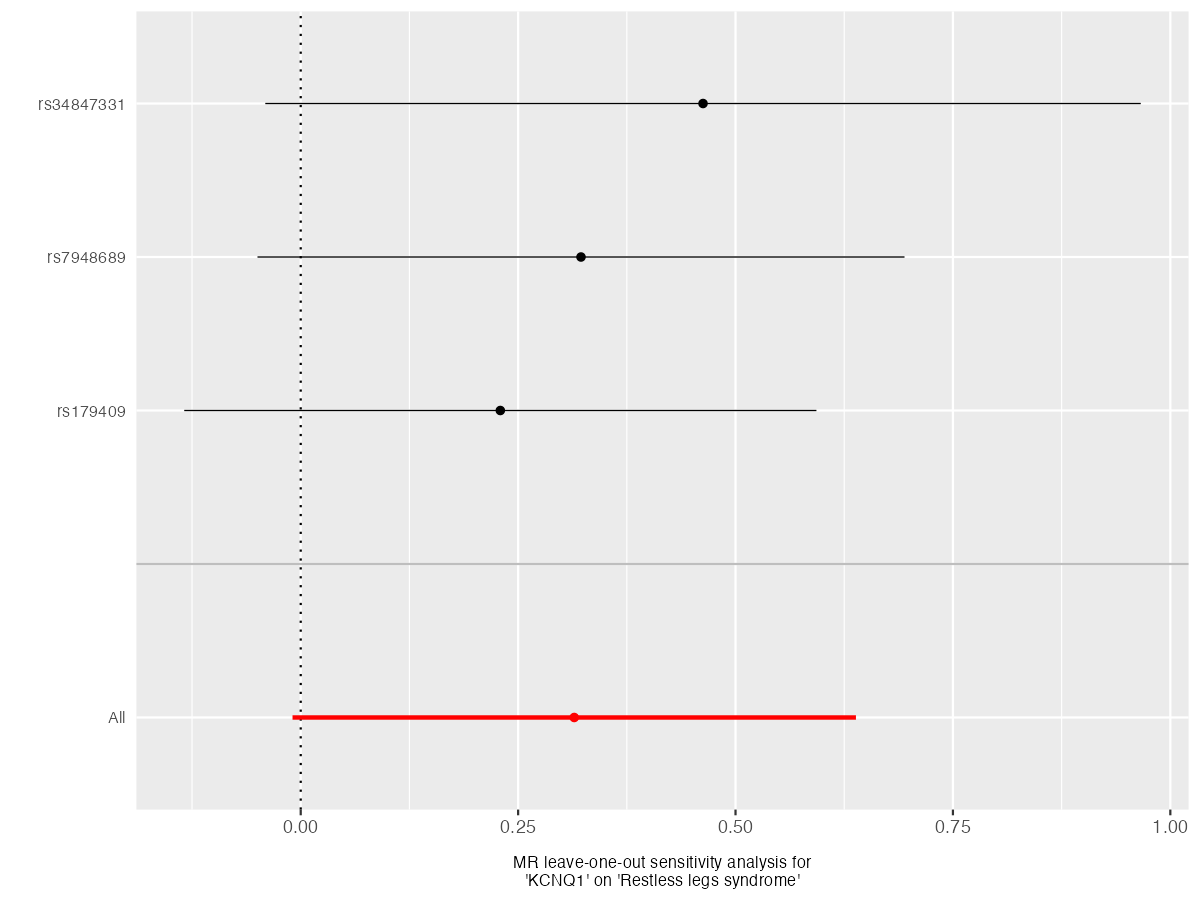

### leaveoneout_plot.png

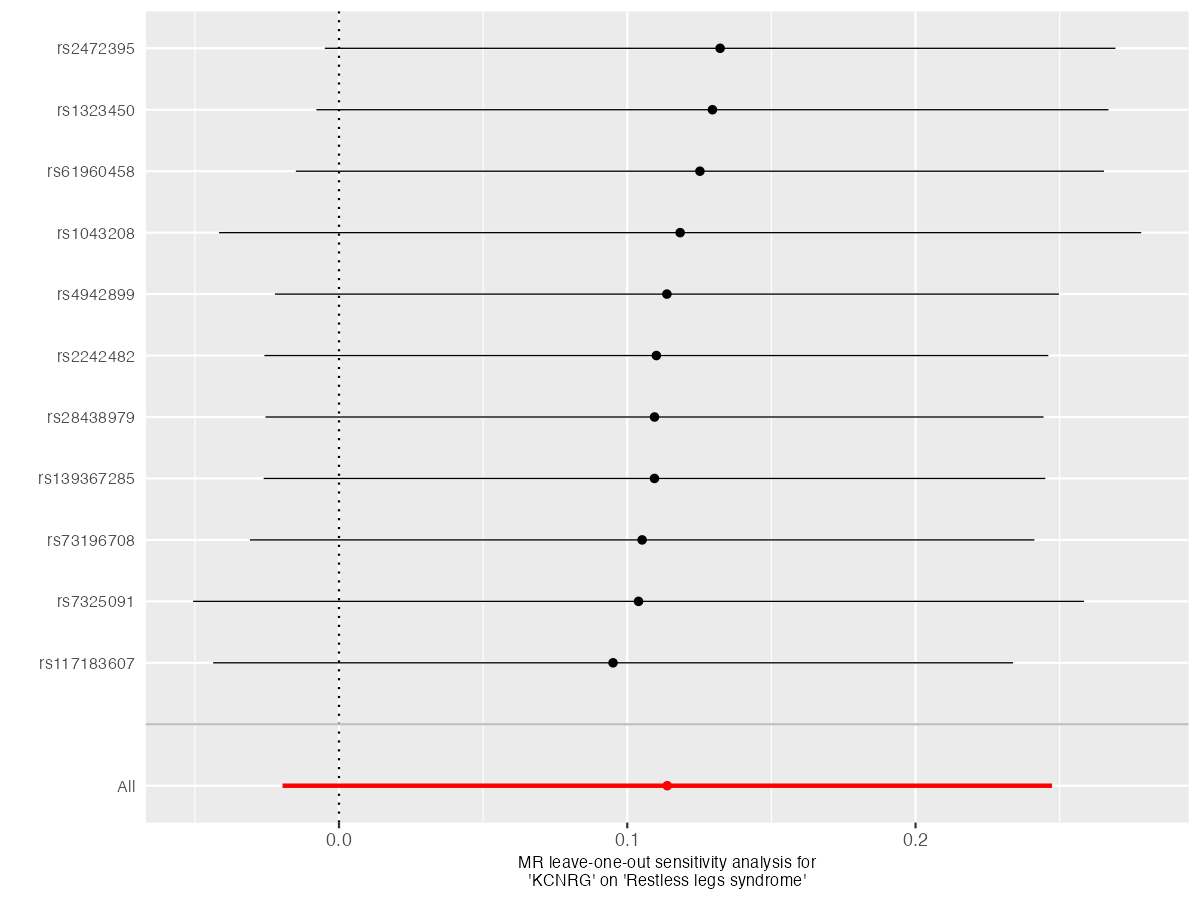

### leaveoneout_plot.png

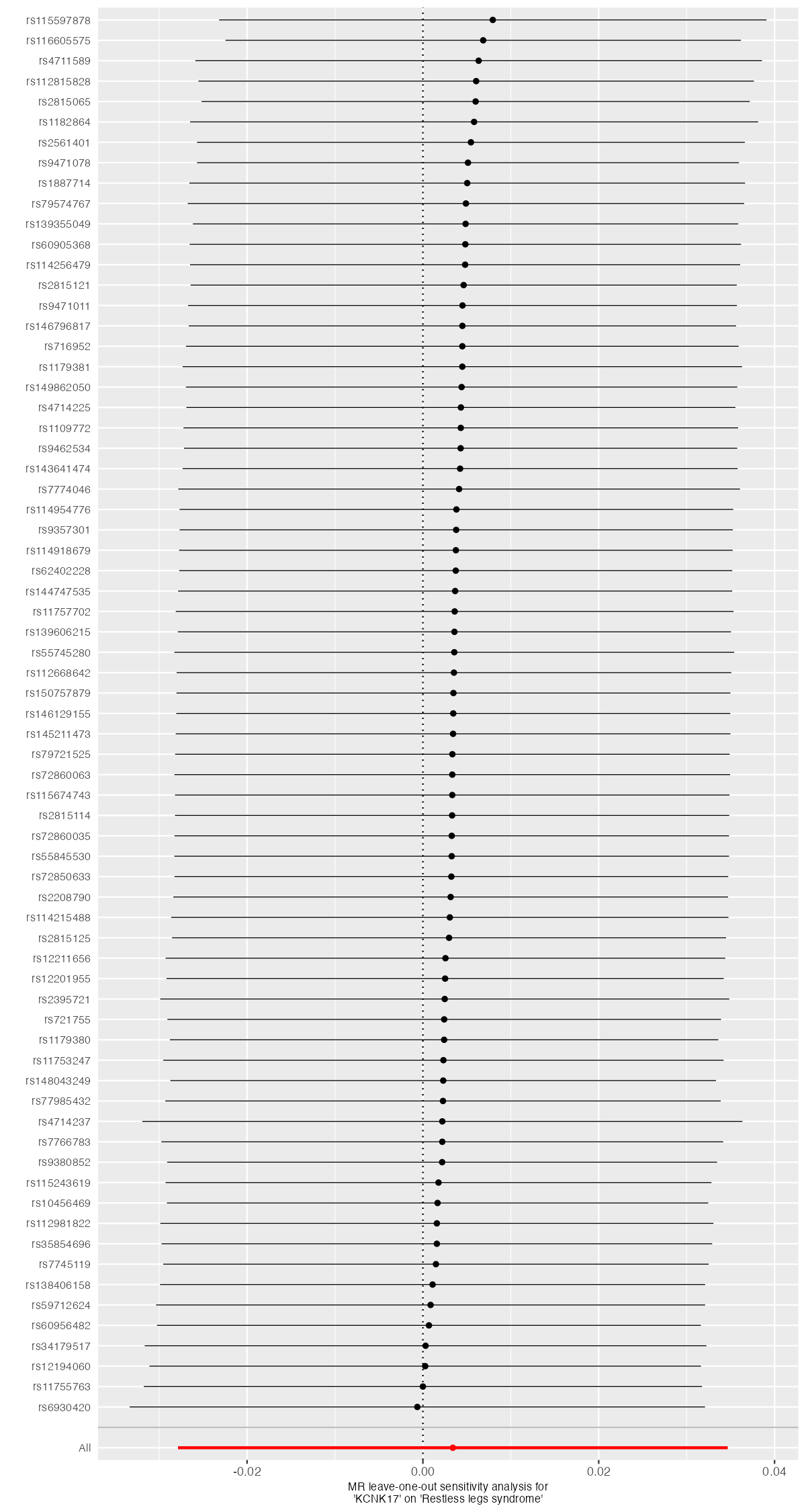

### leaveoneout_plot.png

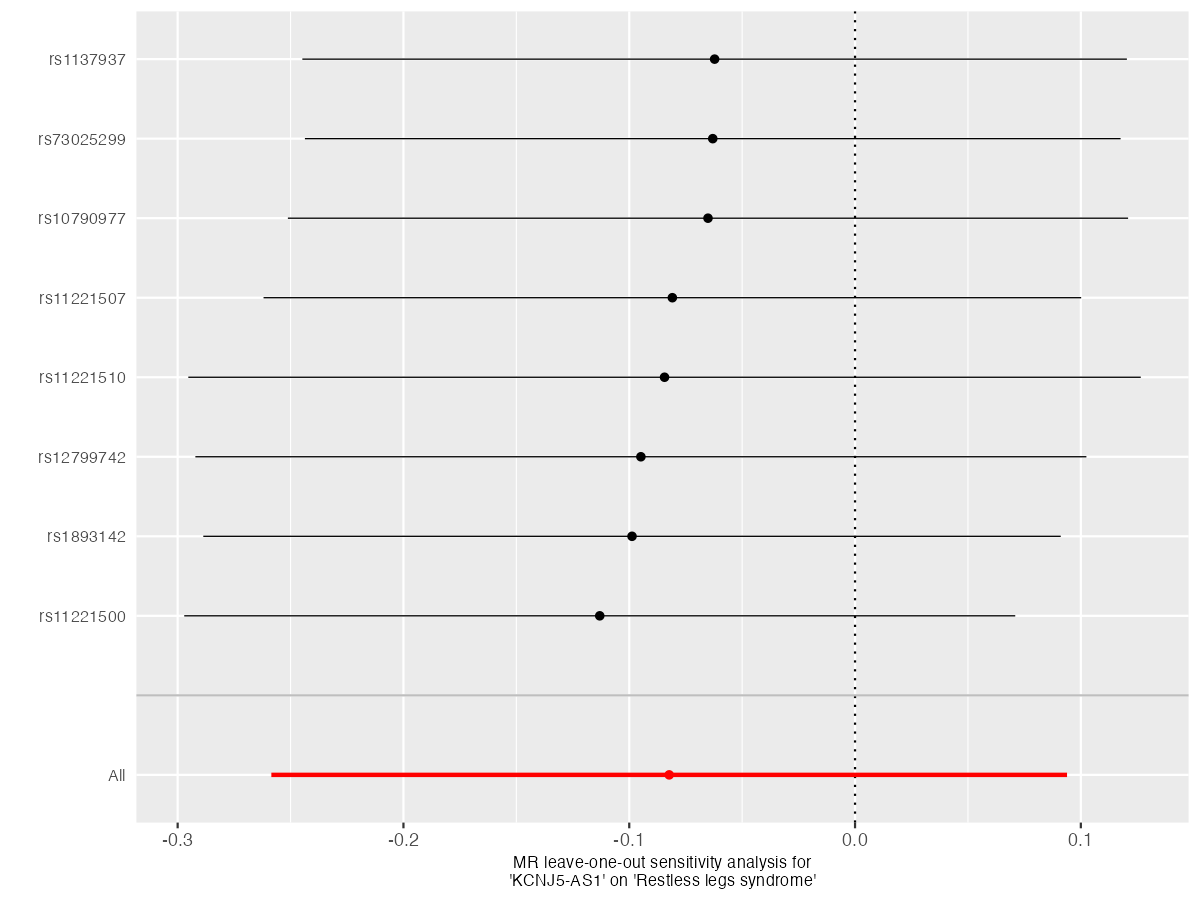

### leaveoneout_plot.png

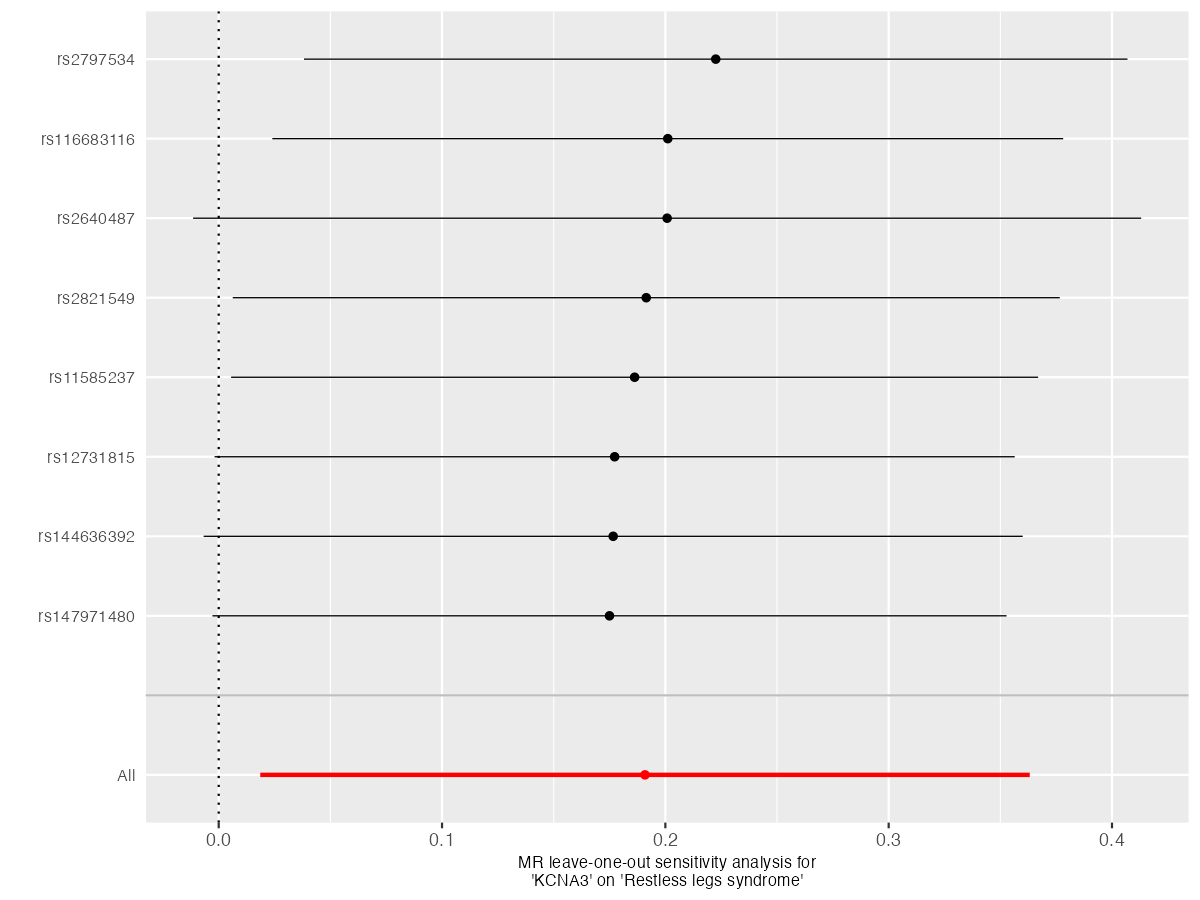

### leaveoneout_plot.png

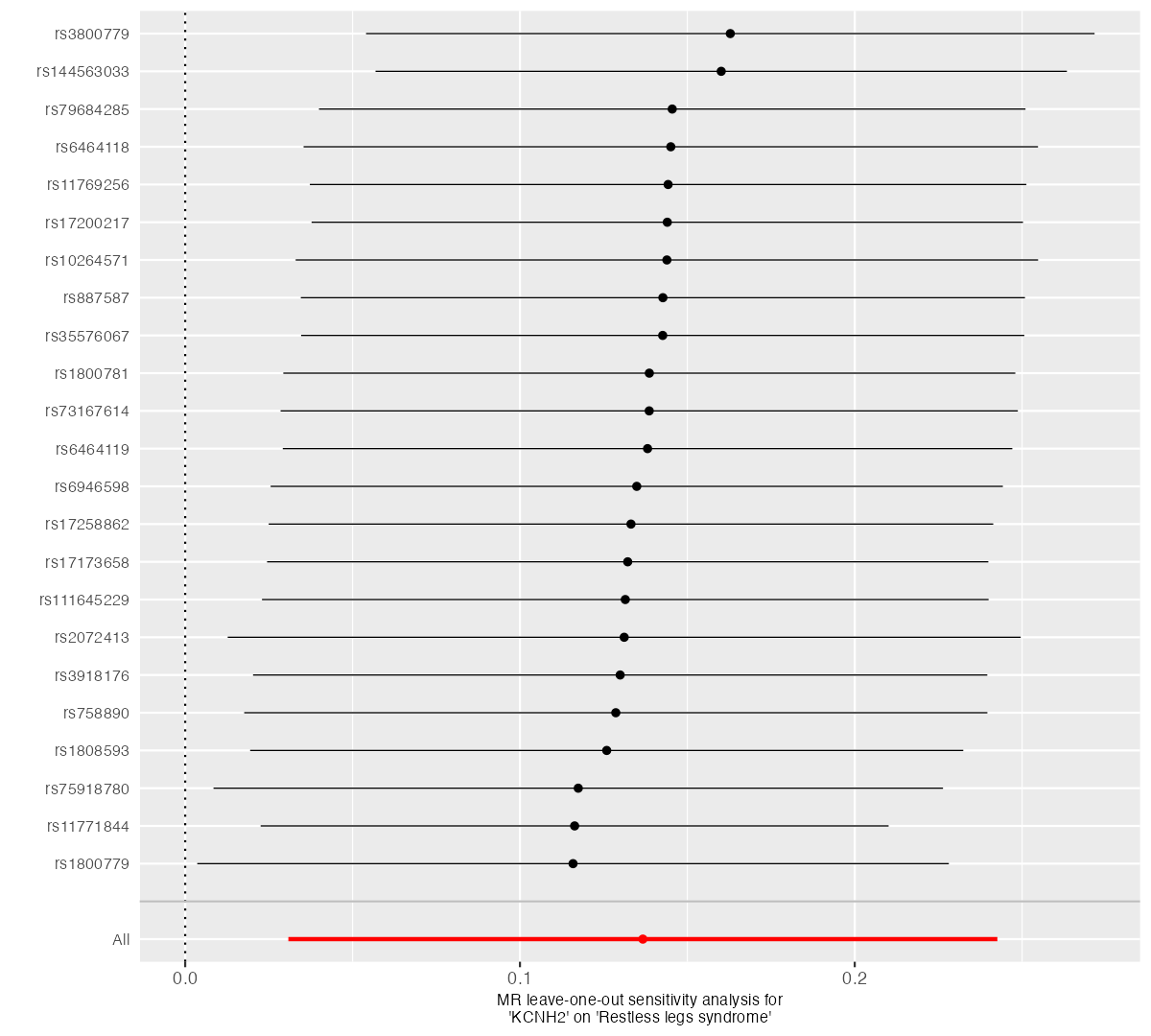

### leaveoneout_plot.png

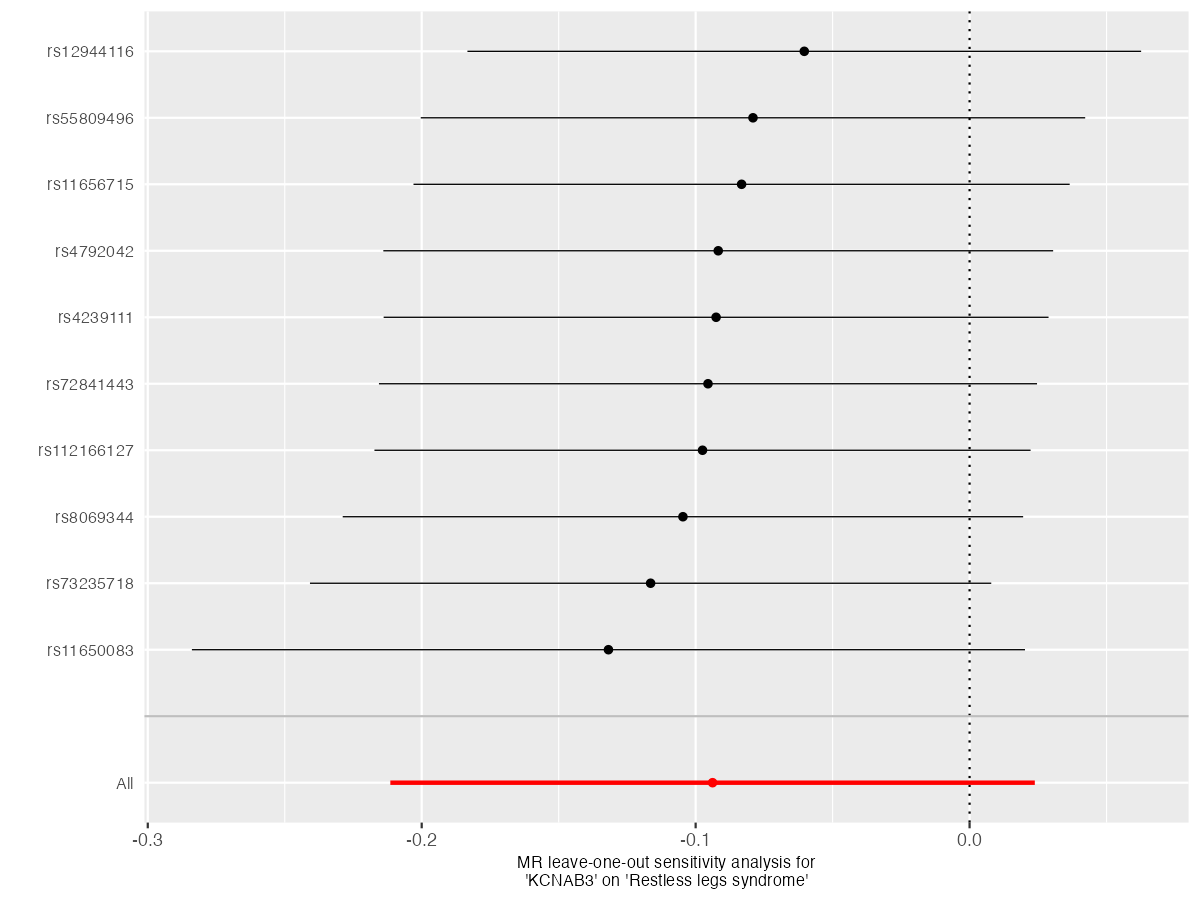

### leaveoneout_plot.png

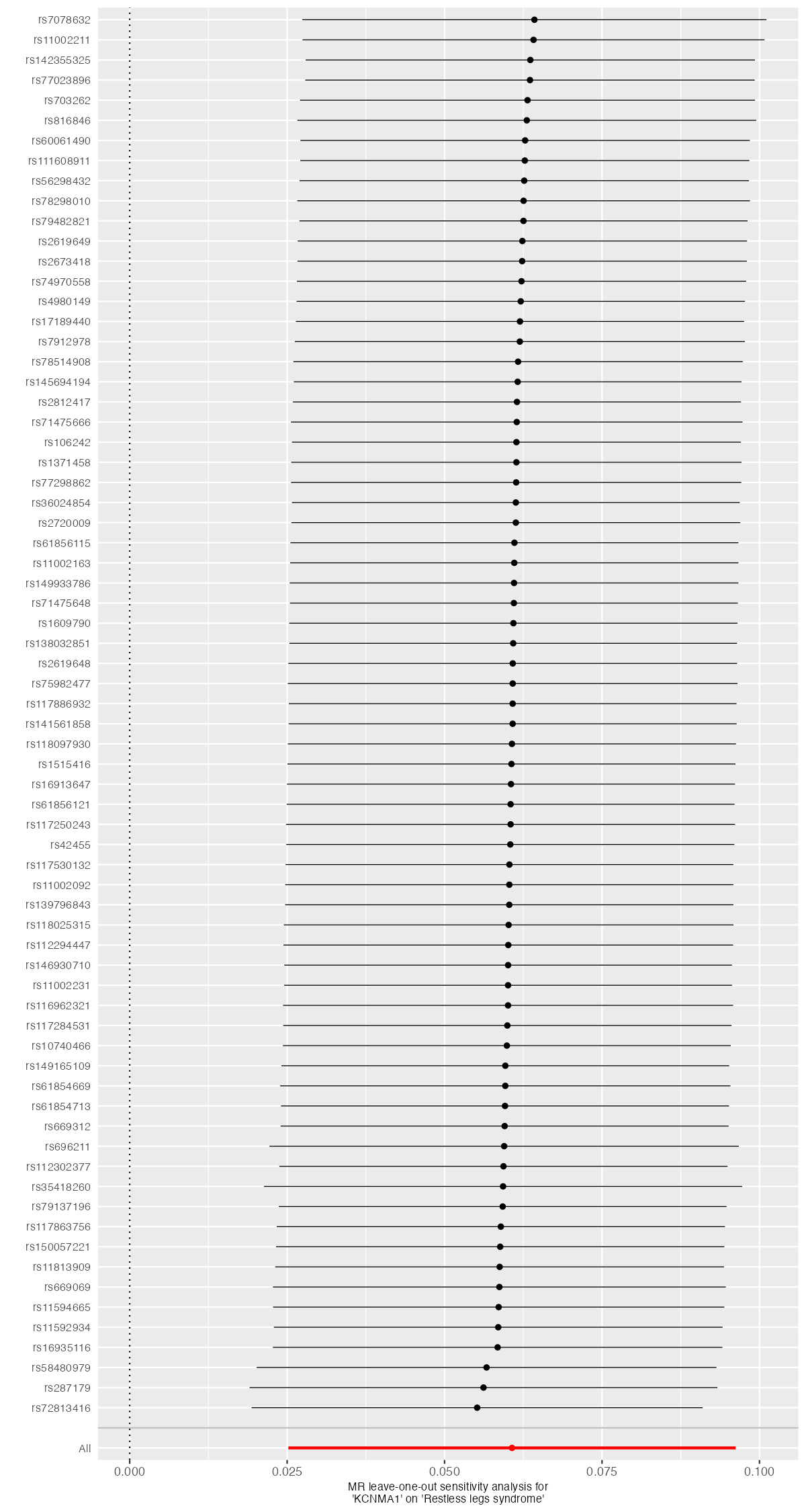

### leaveoneout_plot.png

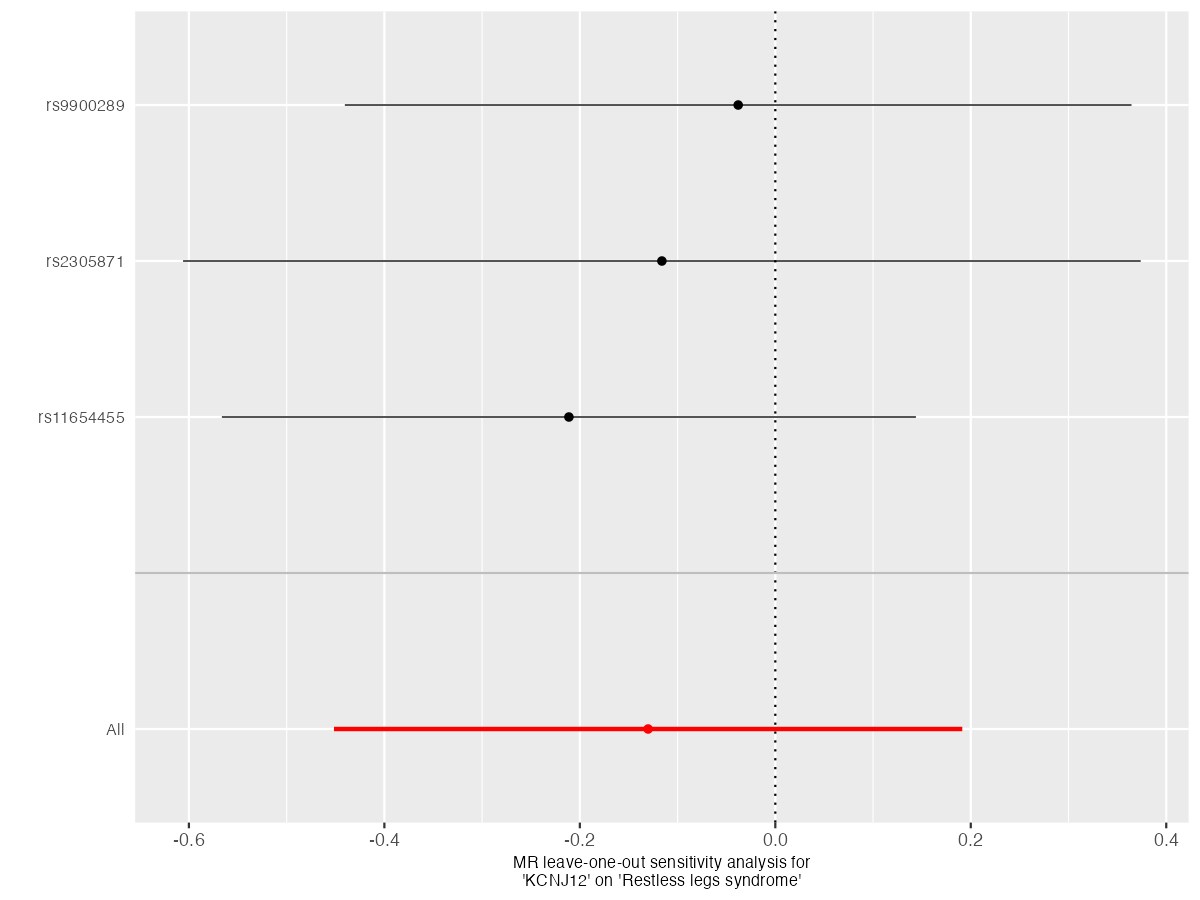

### leaveoneout_plot.png

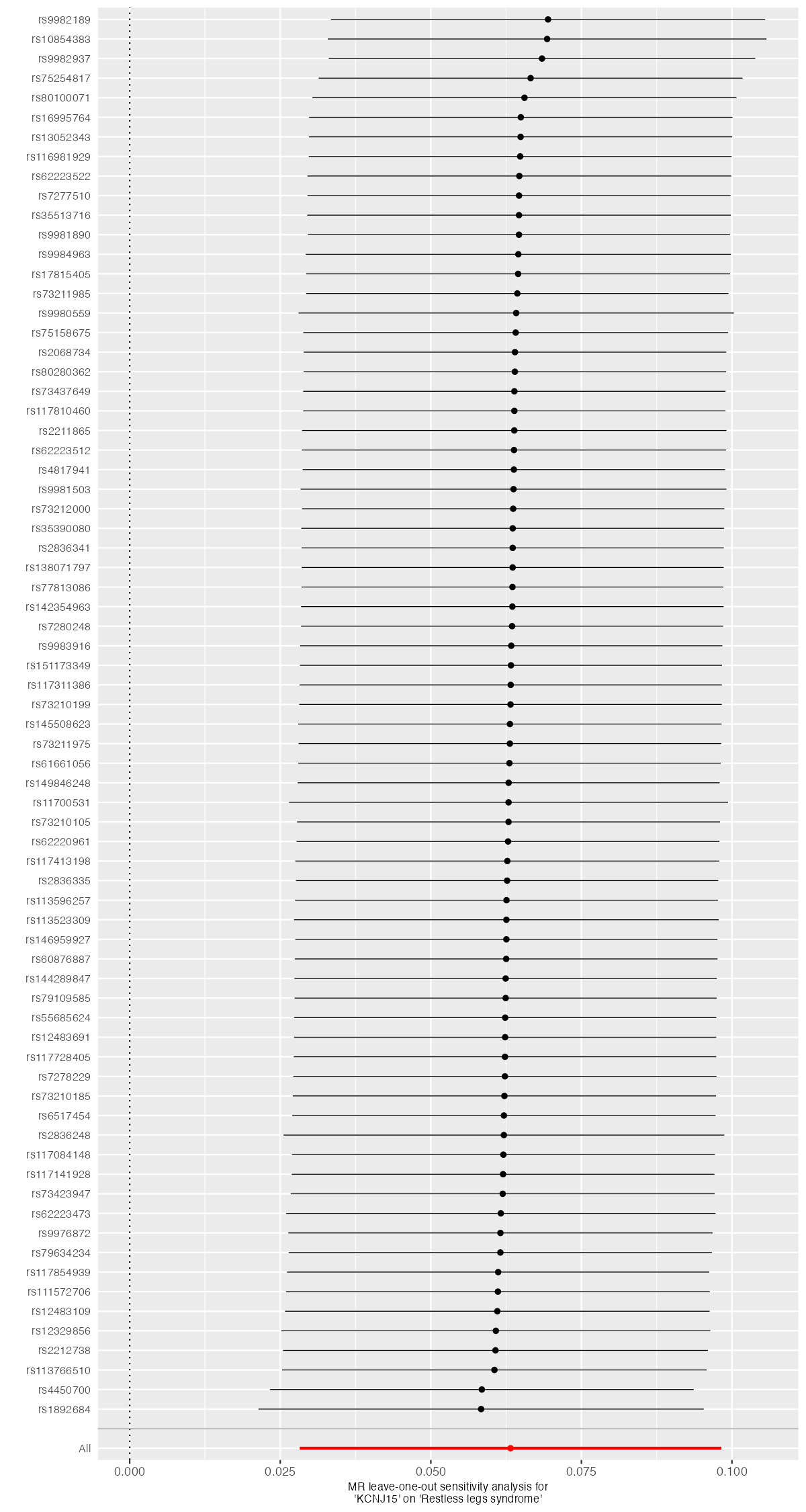

### leaveoneout_plot.png

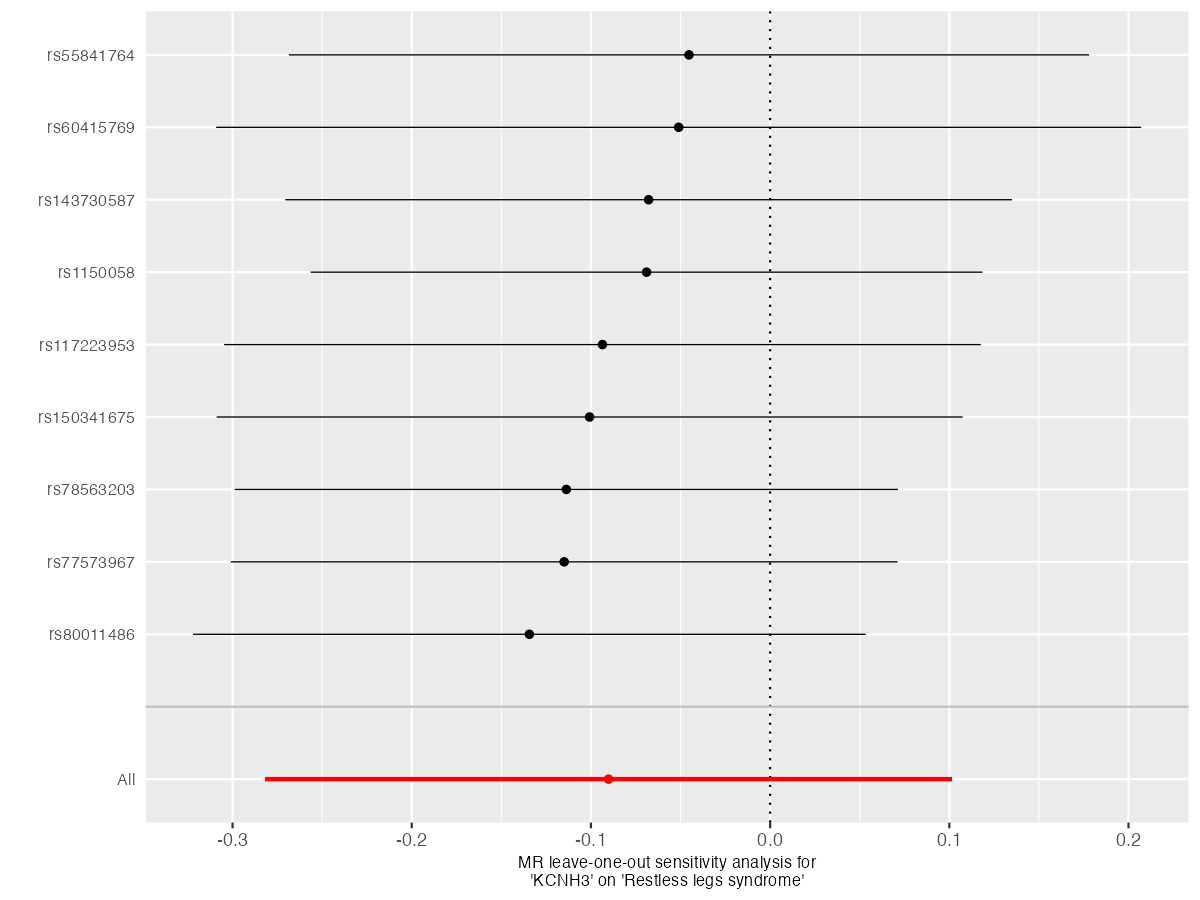

### scatter_plot.png

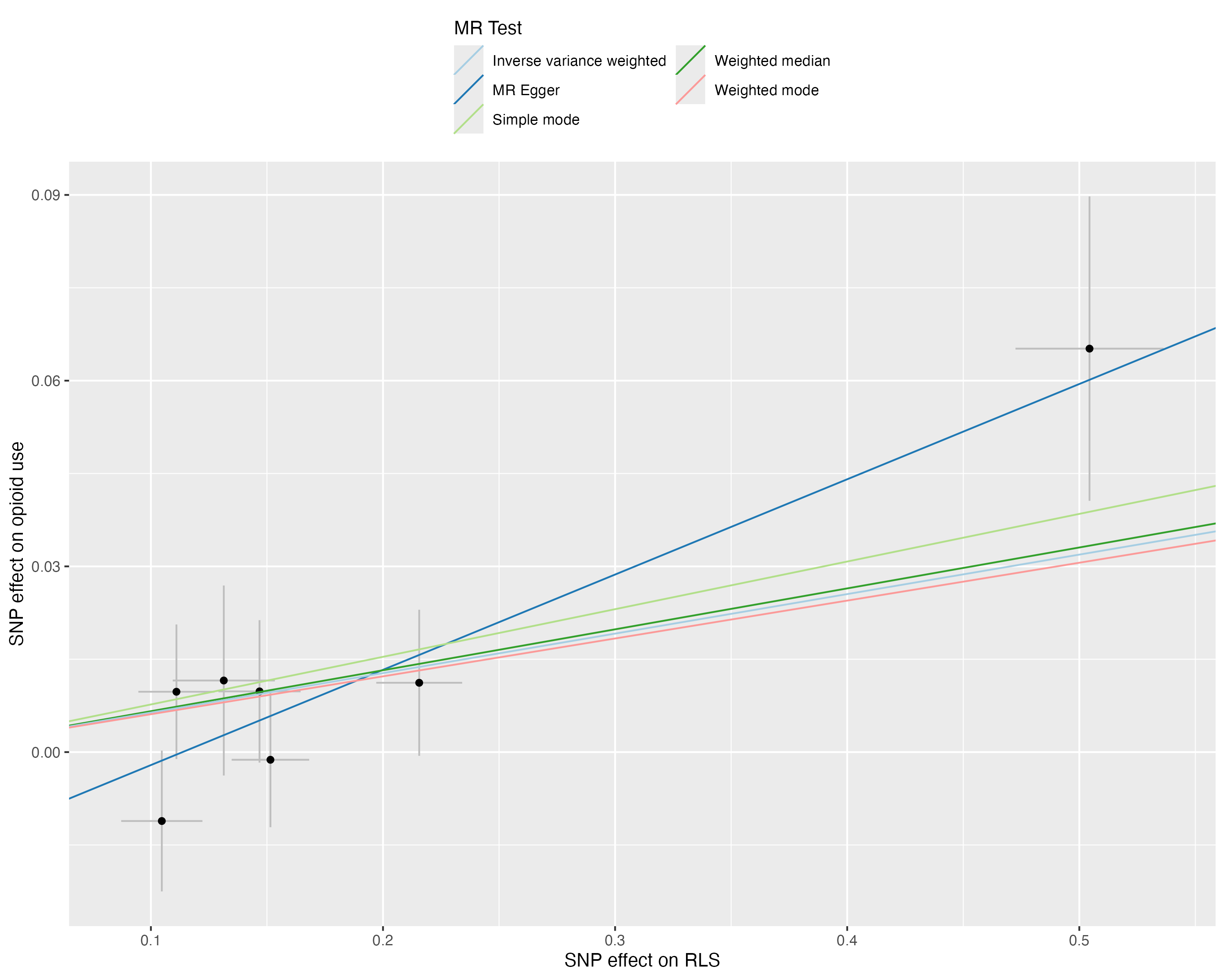

### scatter_plot_1.png

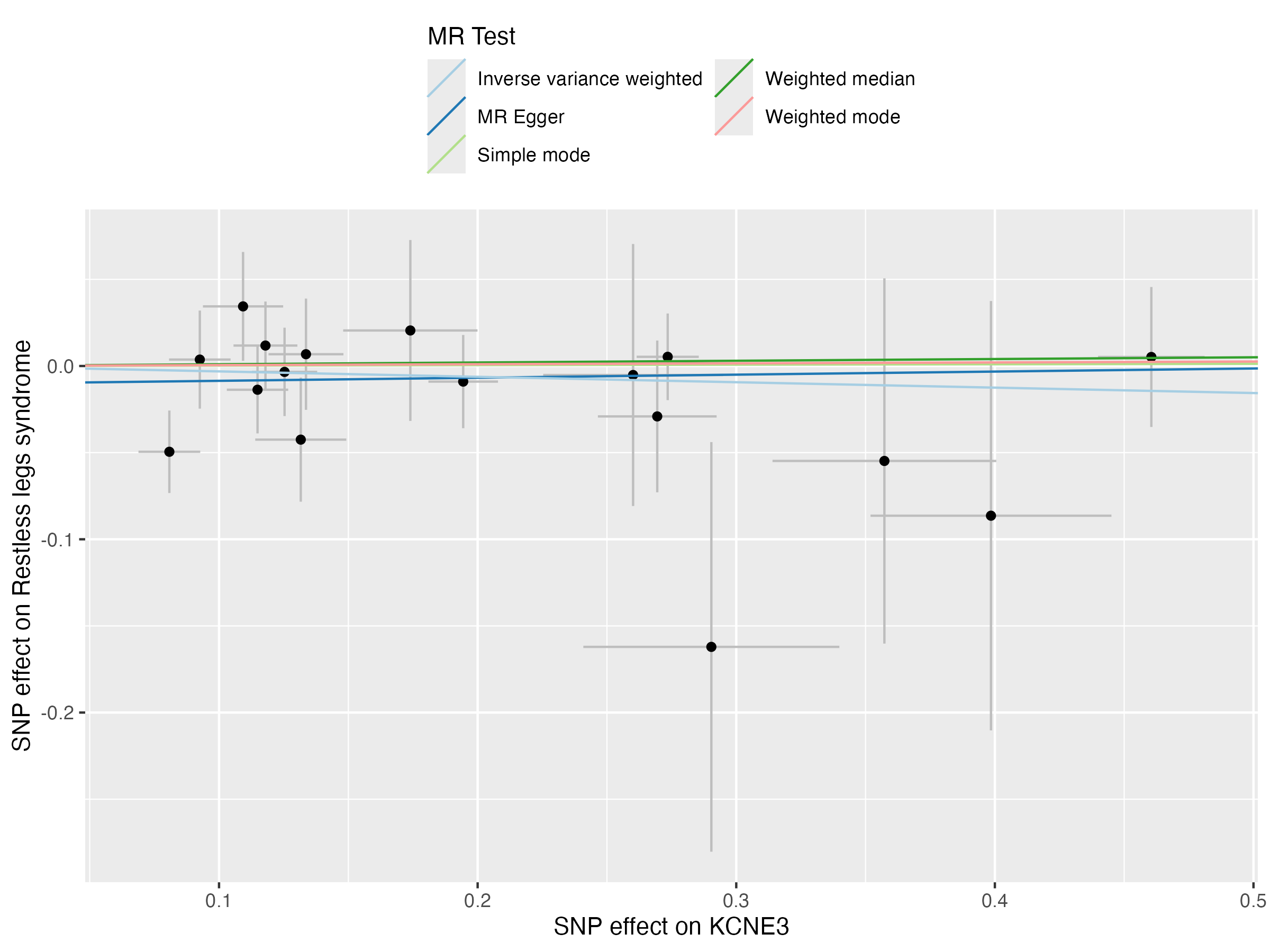

### scatter_plot_1.png

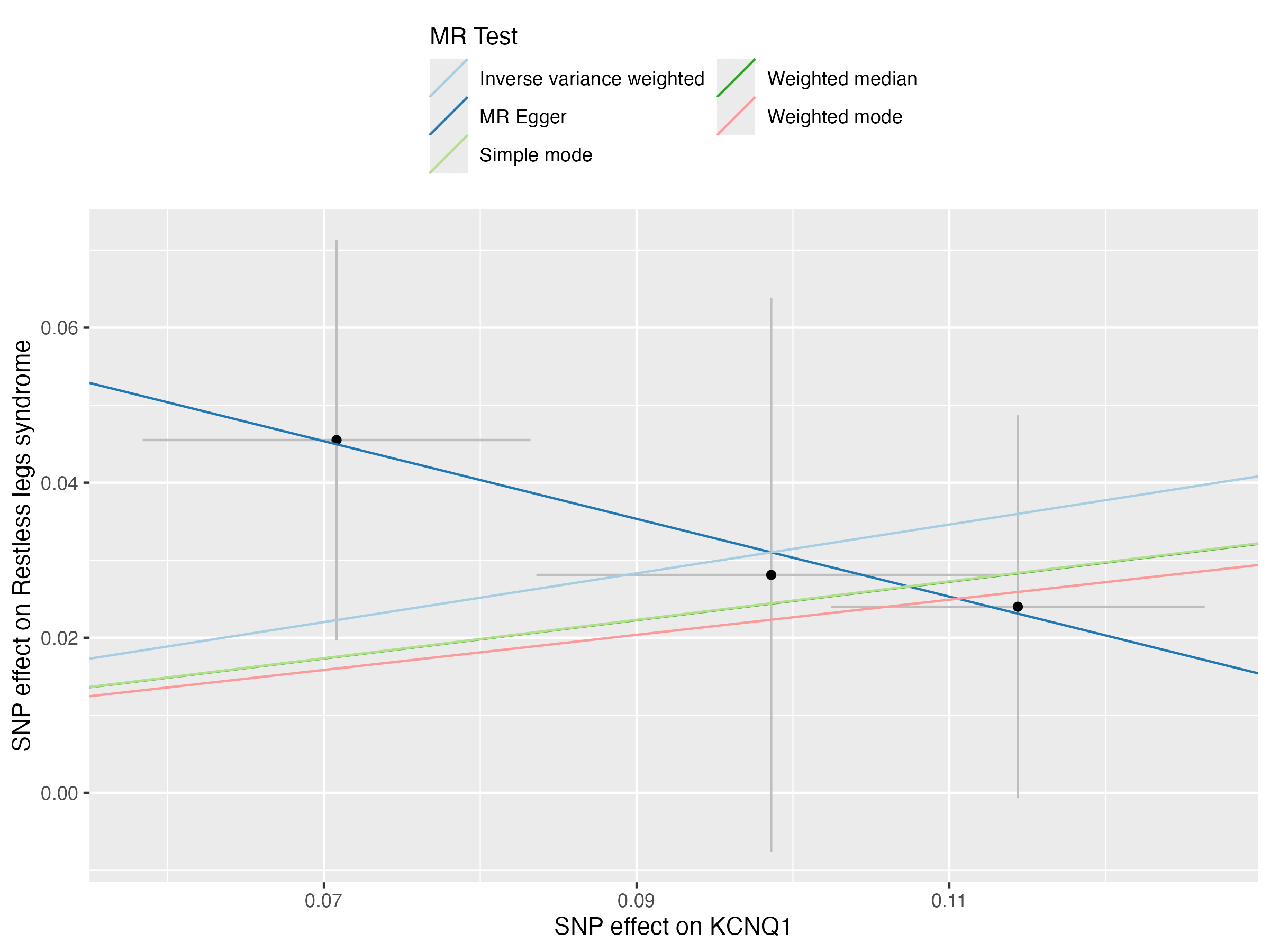

### scatter_plot_1.png

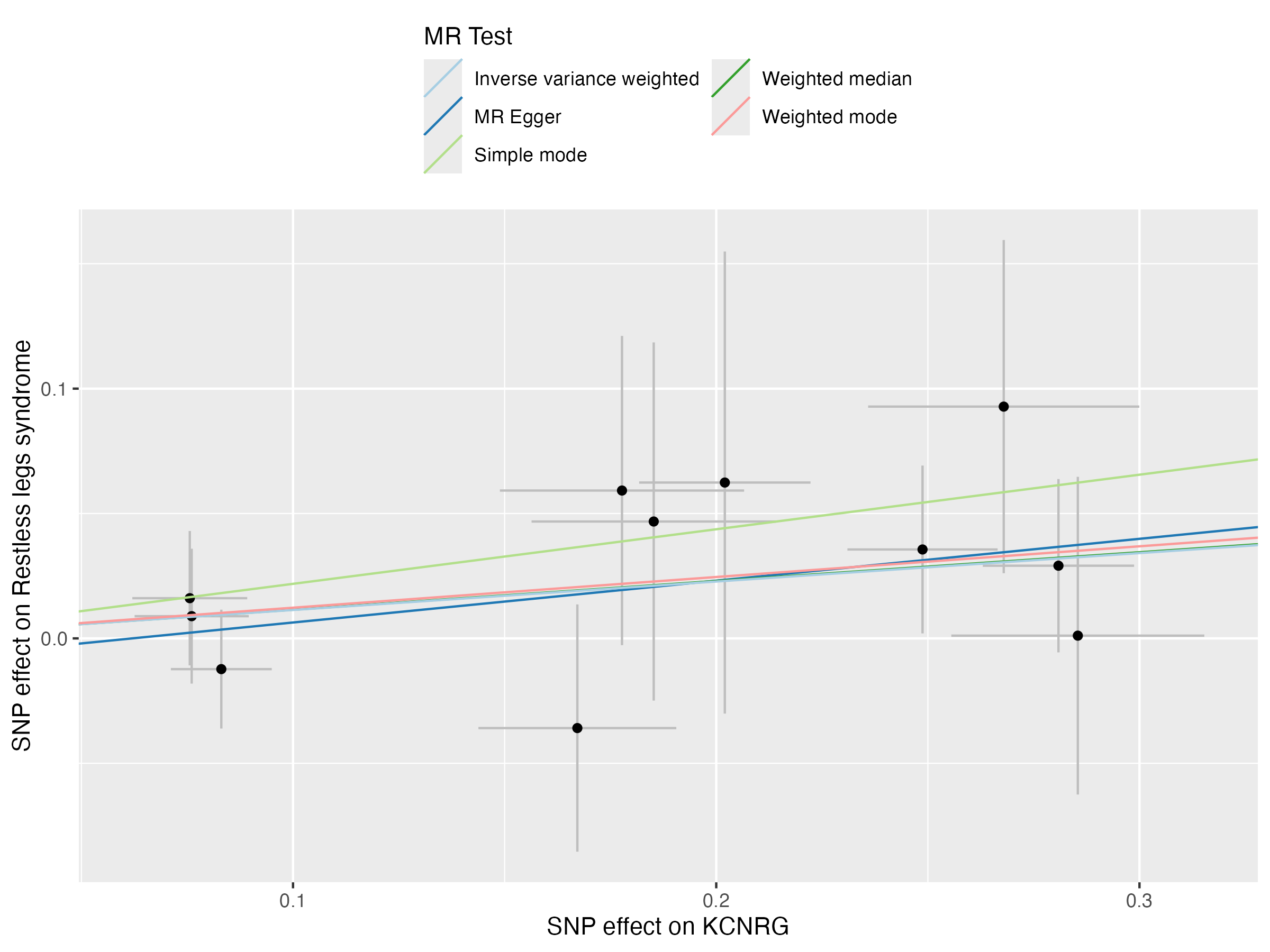

### scatter_plot_1.png

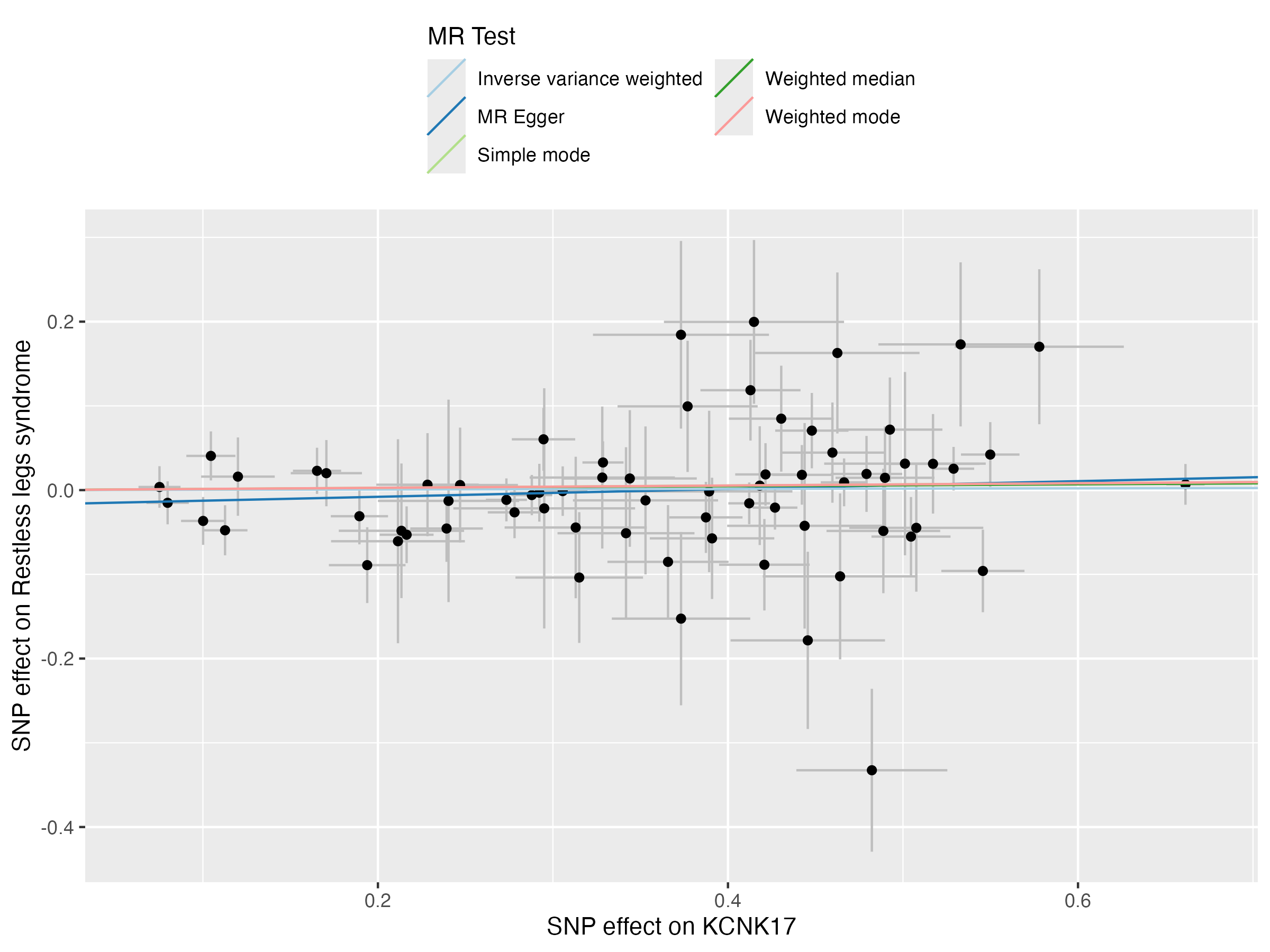

### scatter_plot_1.png

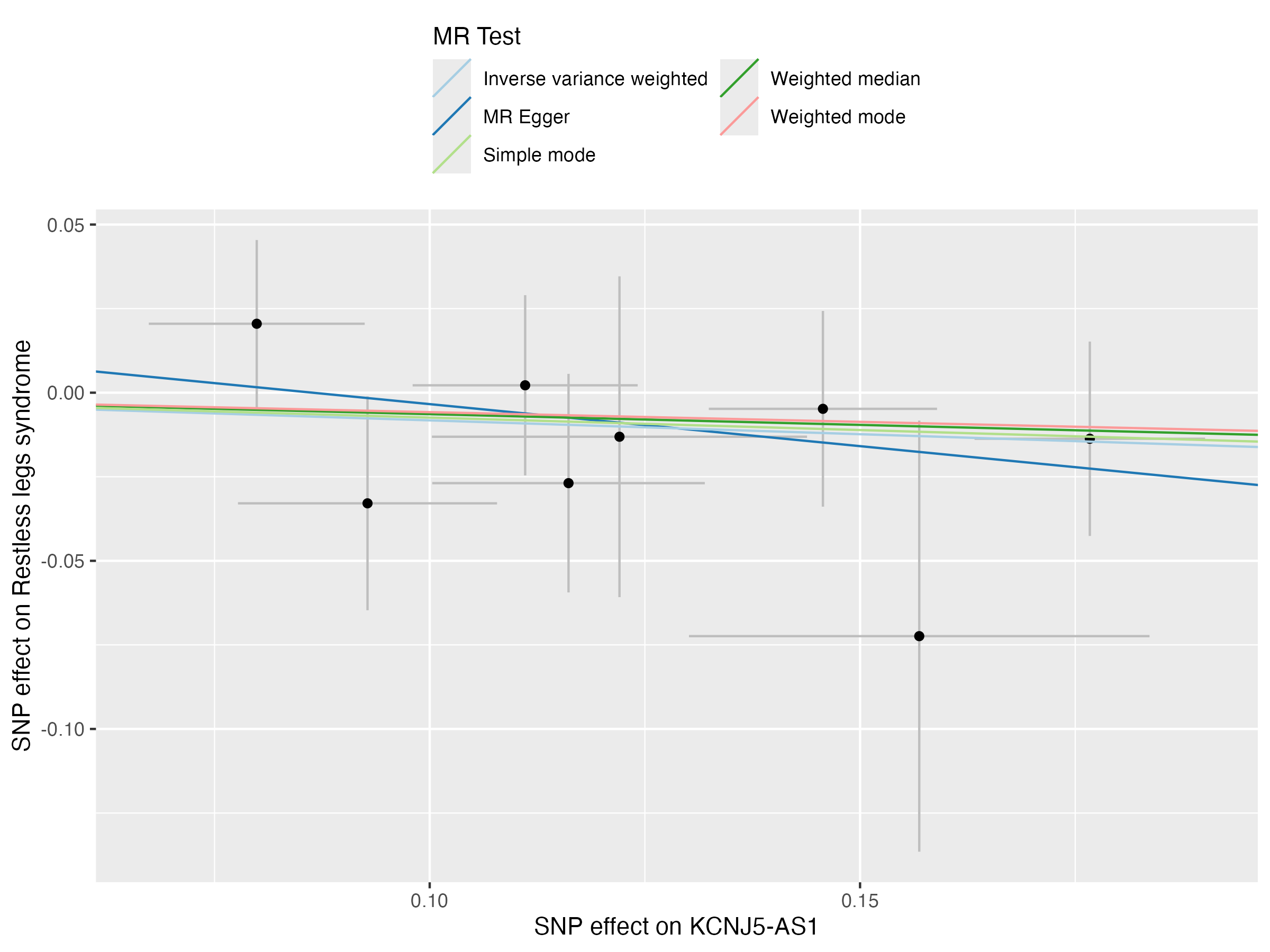

### scatter_plot_1.png

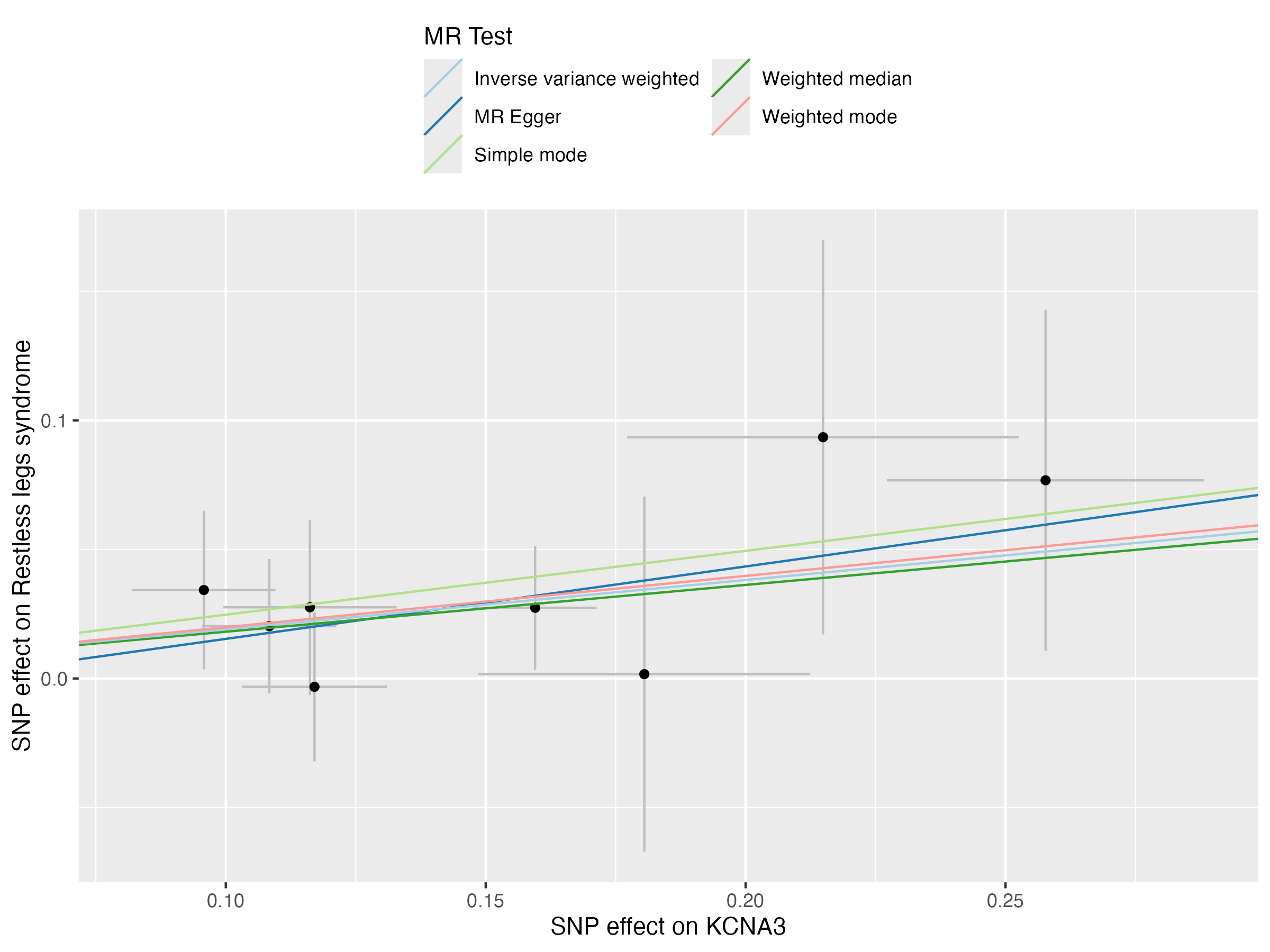

### scatter_plot_1.png

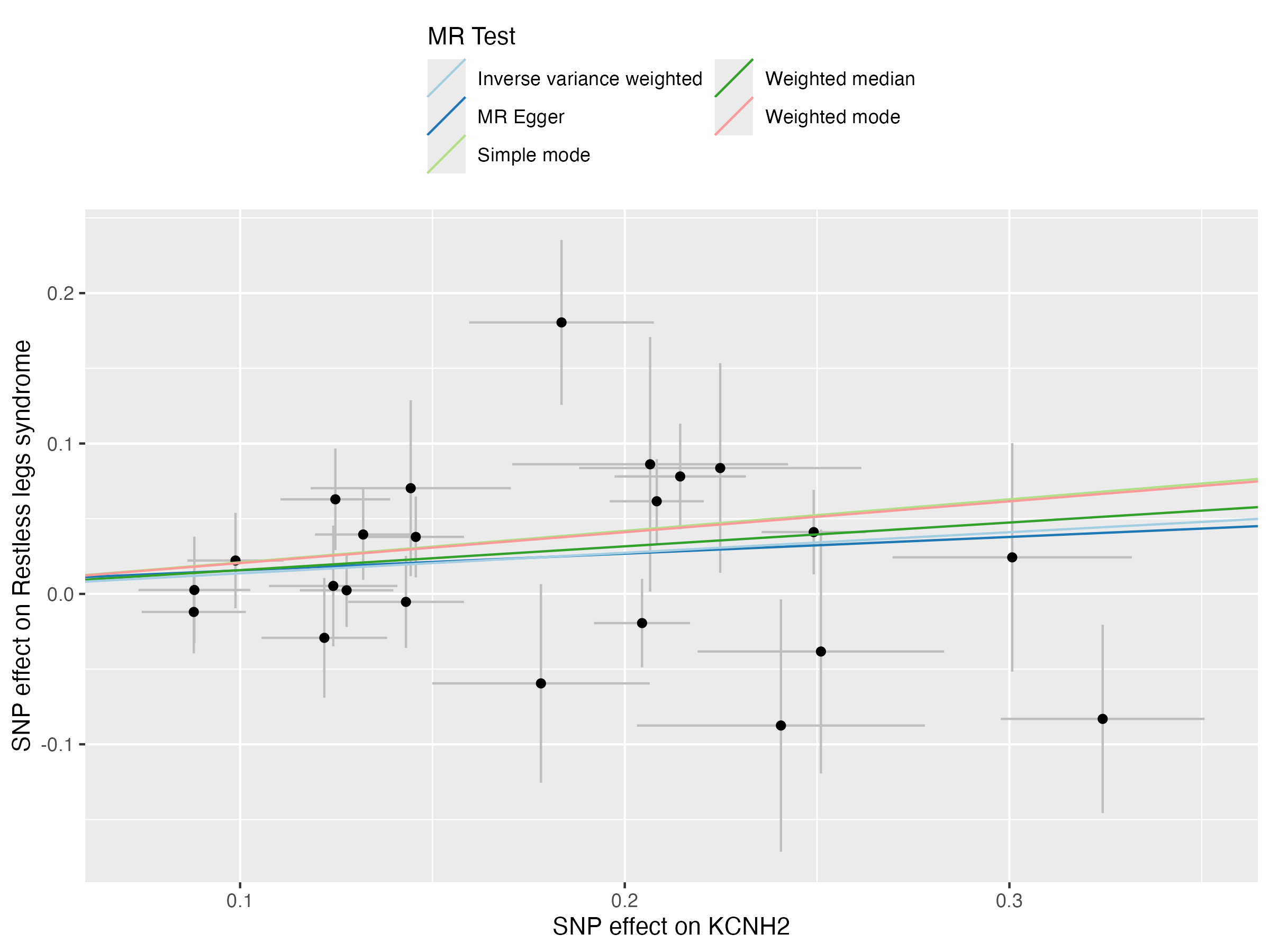

### scatter_plot_1.png

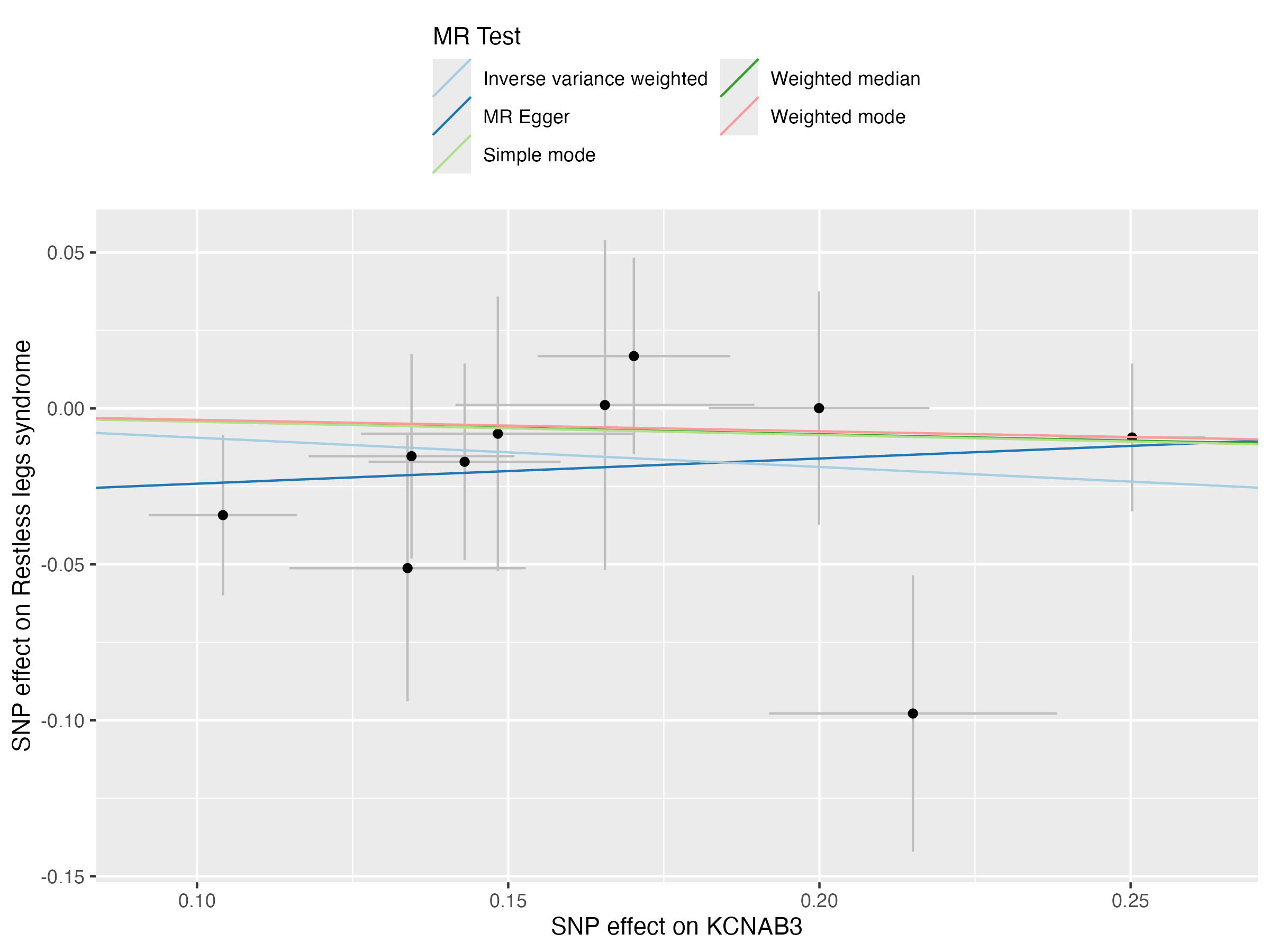

### scatter_plot_1.png

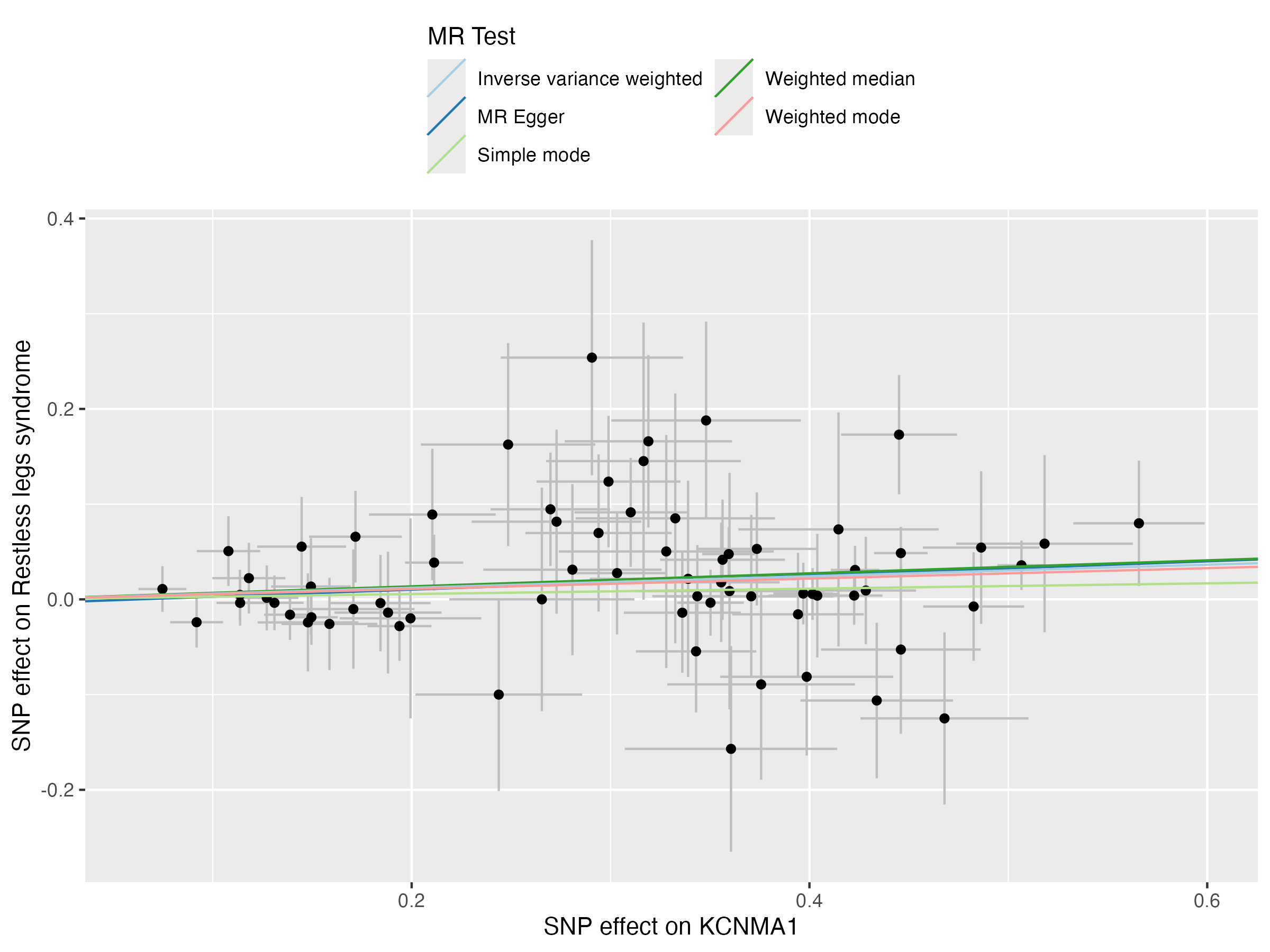

### scatter_plot_1.png

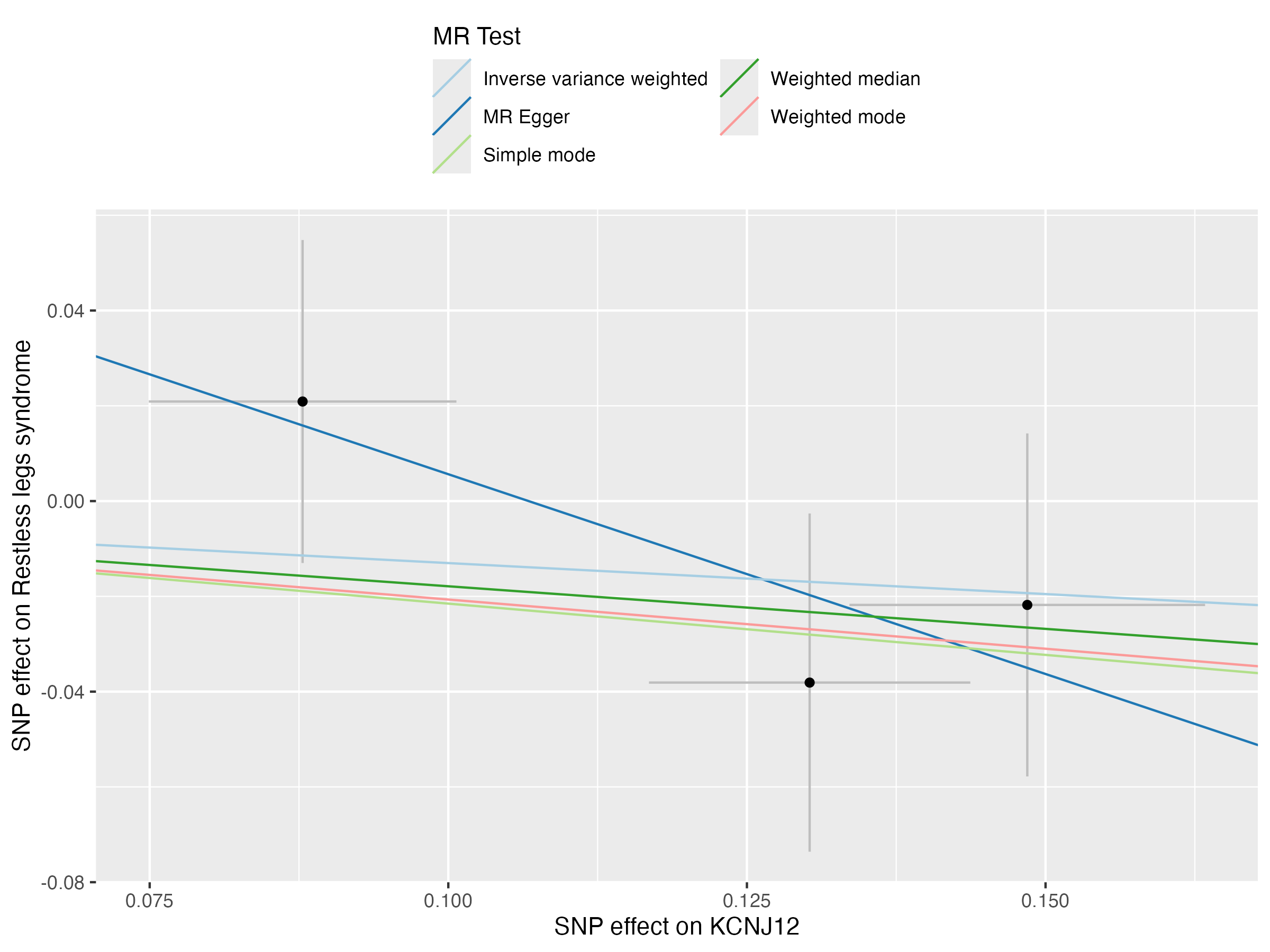

### scatter_plot_1.png

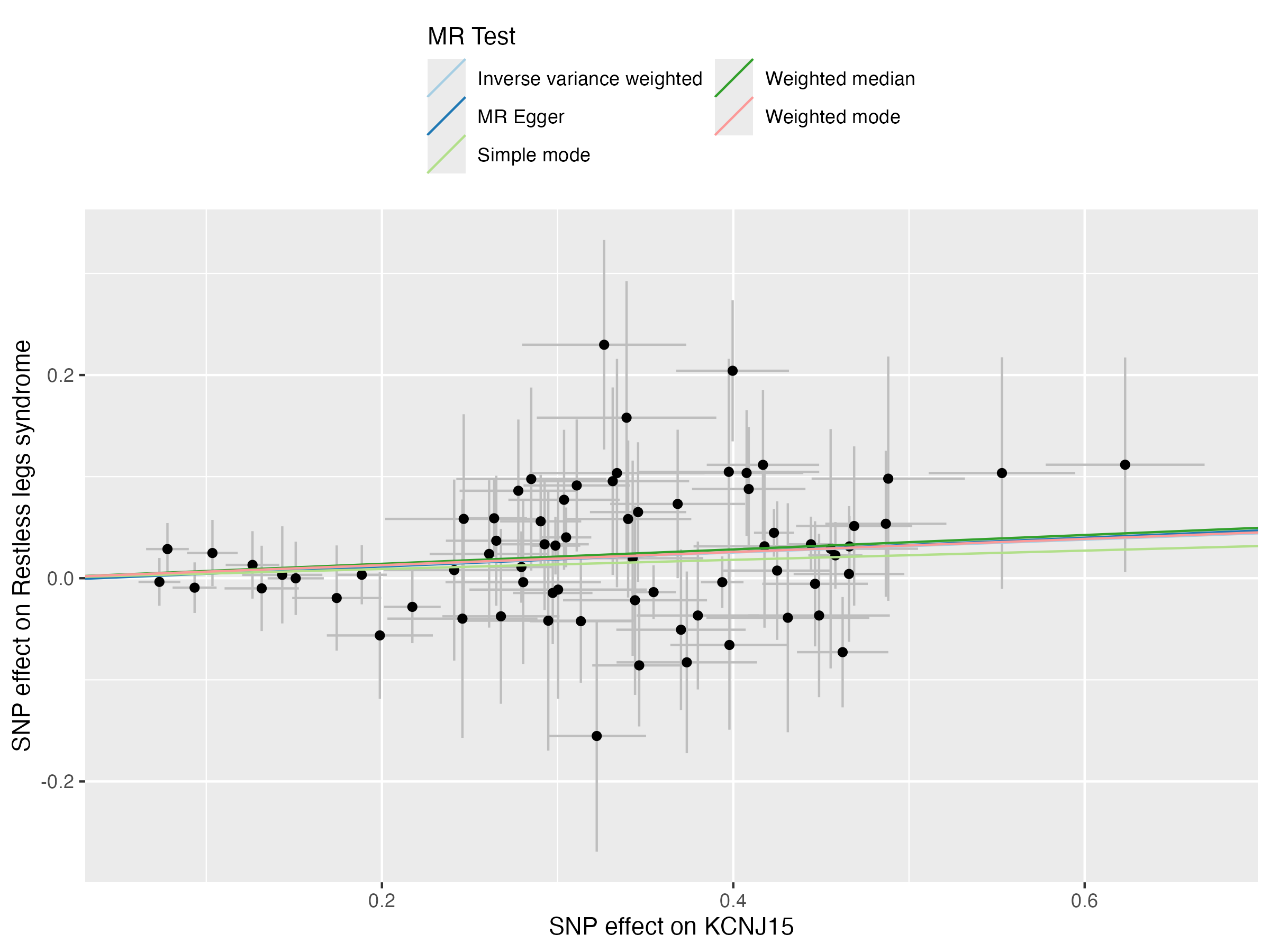

### visualization_circular_heatmap_plot.png

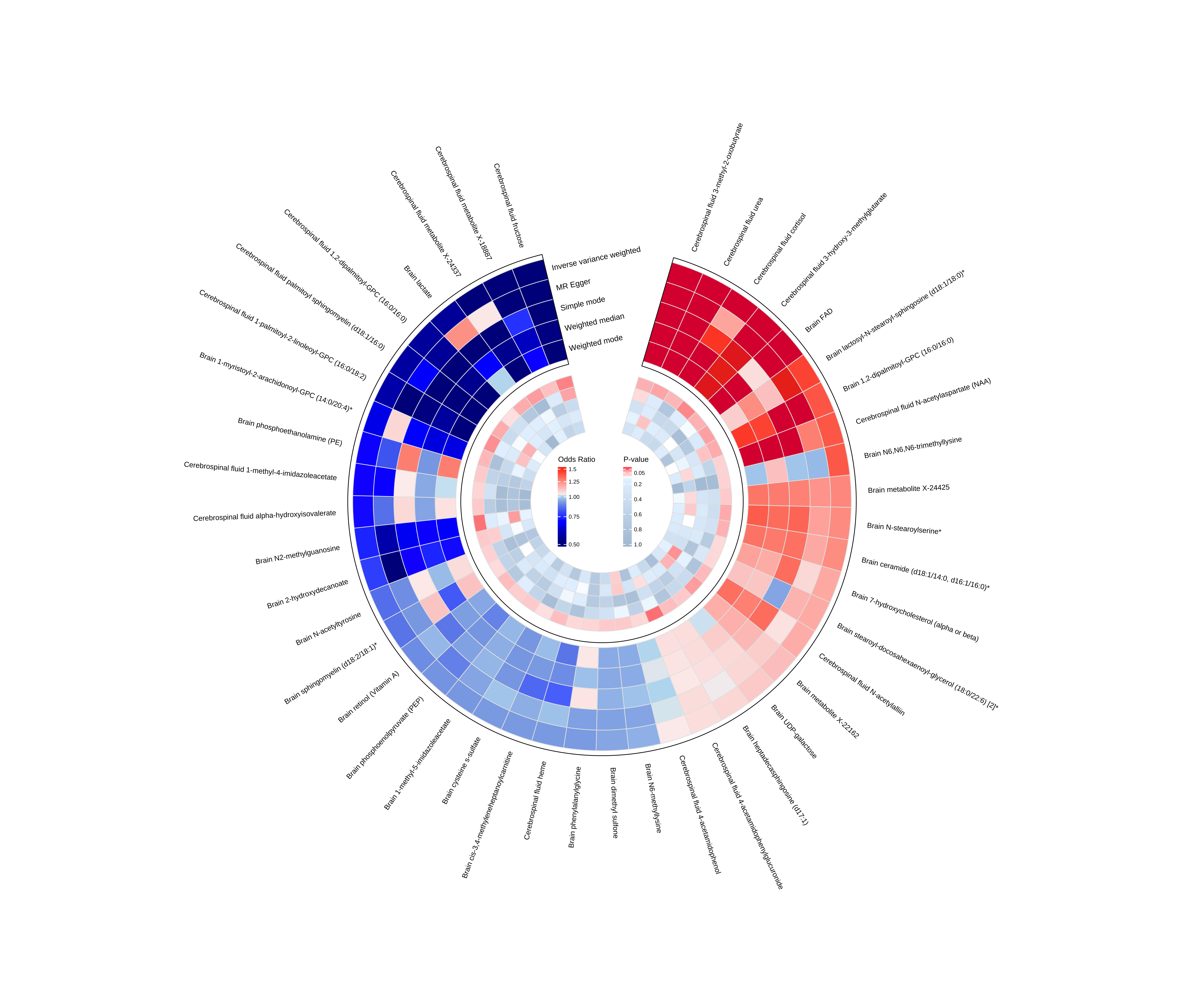

### visualization_circular_heatmap_plot.png

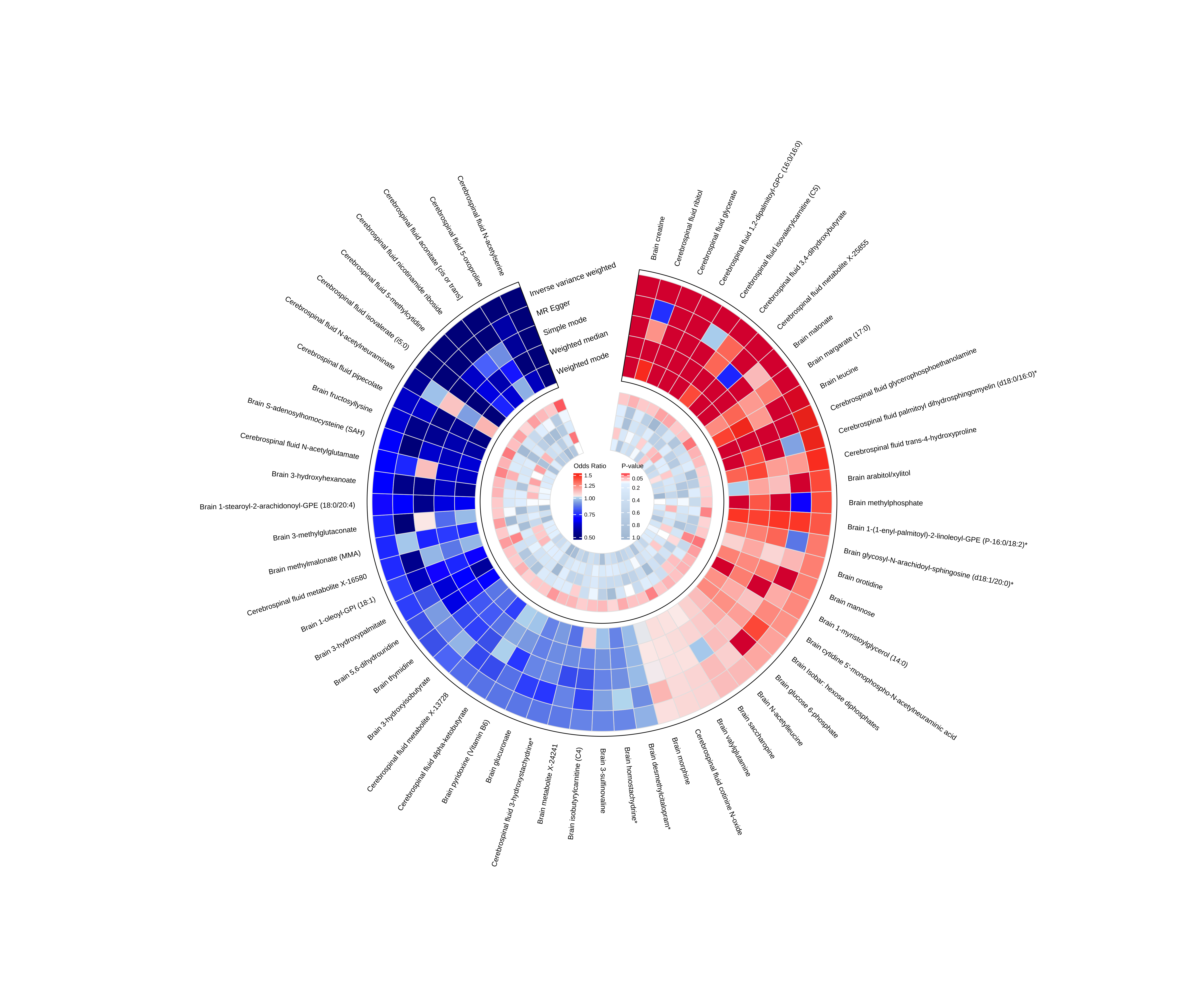

### visualization_forest_plot.png

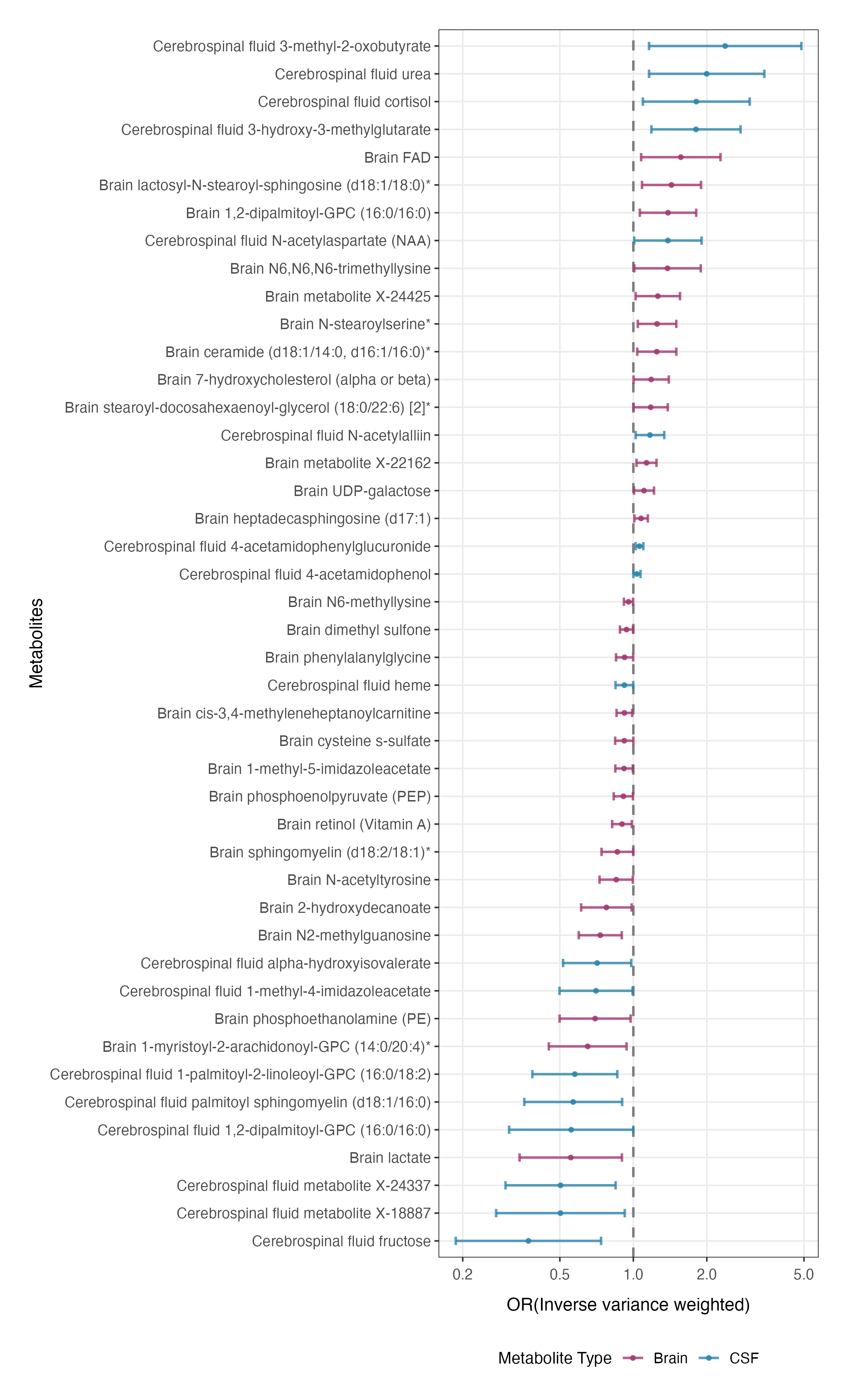

### visualization_forest_plot.png

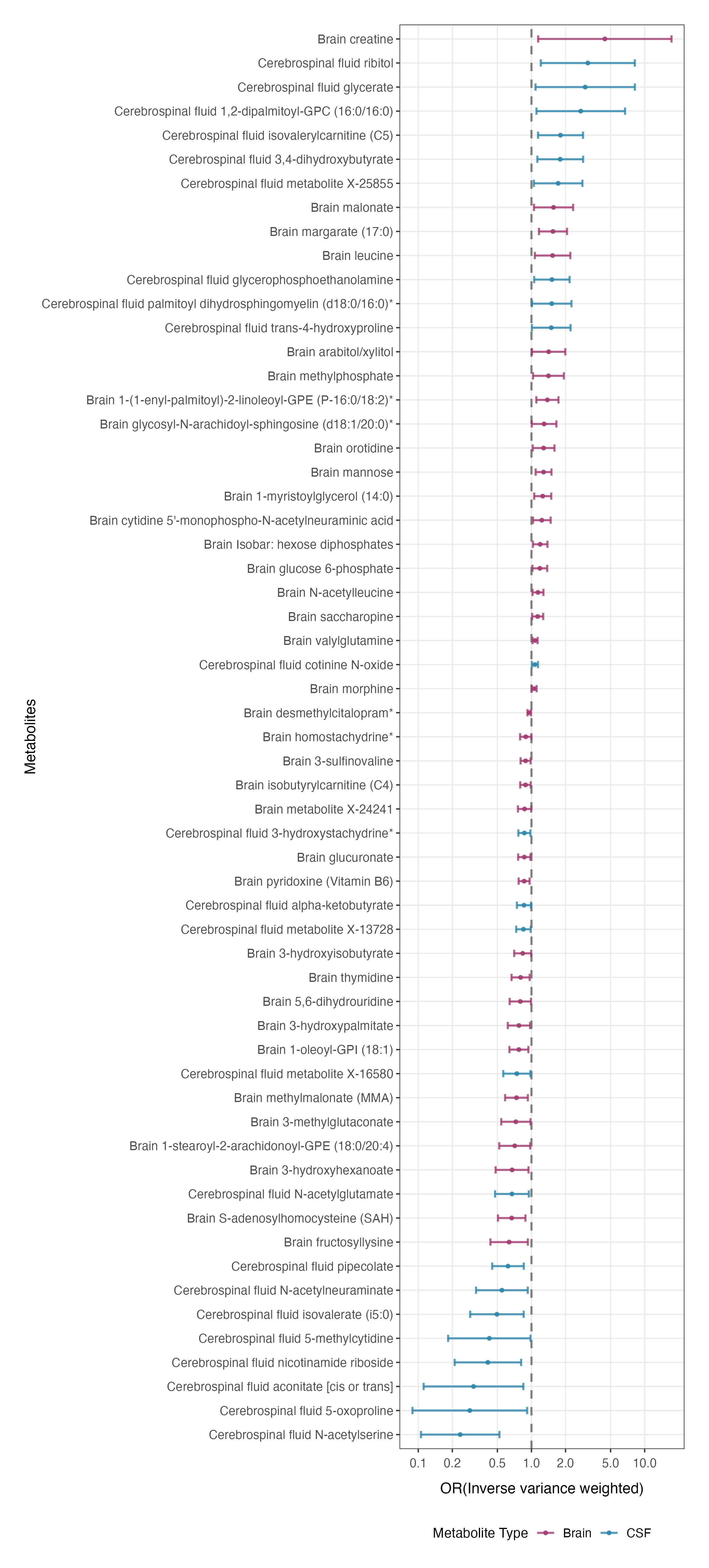

### visualization_table.png

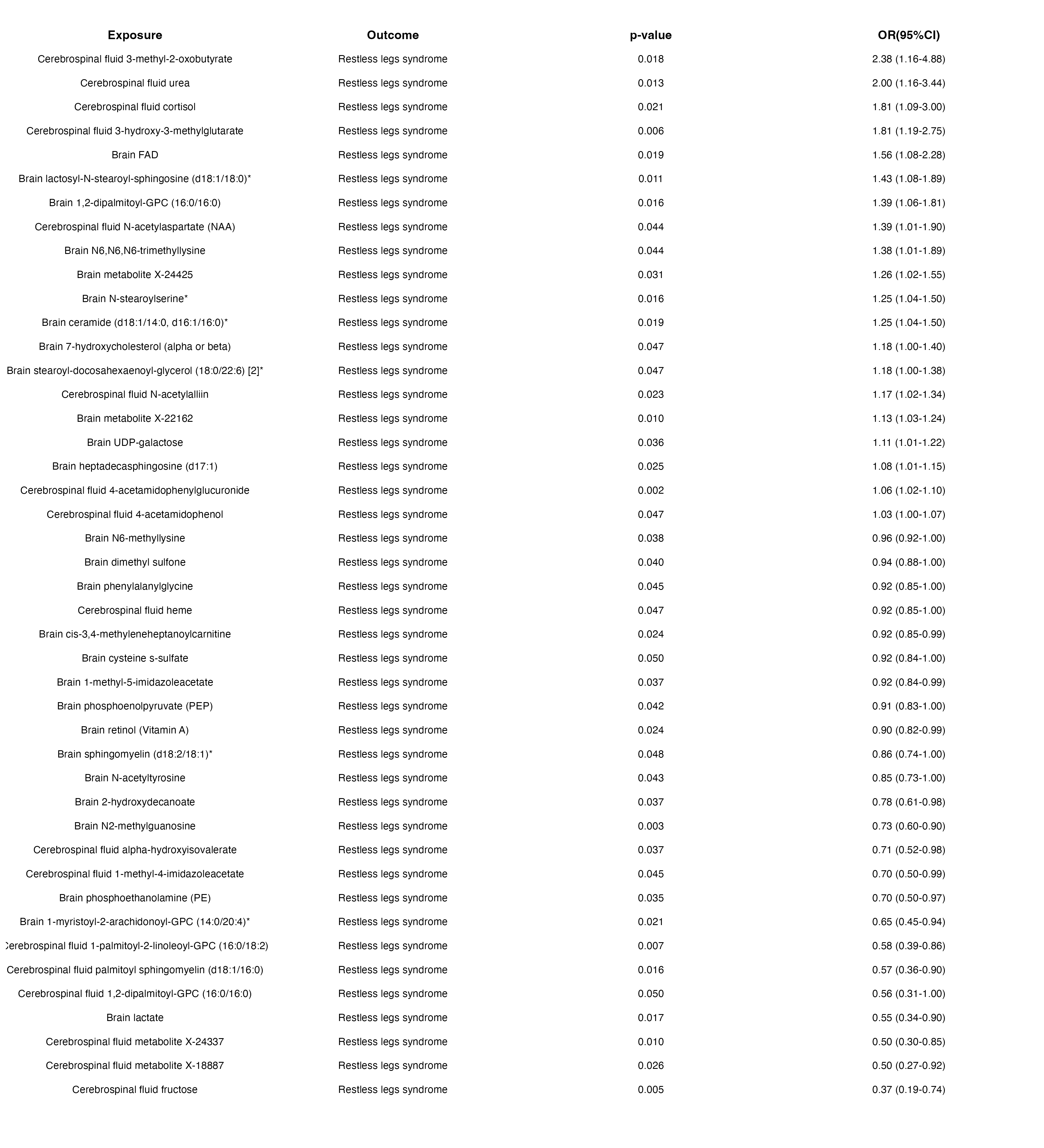

### visualization_table.png

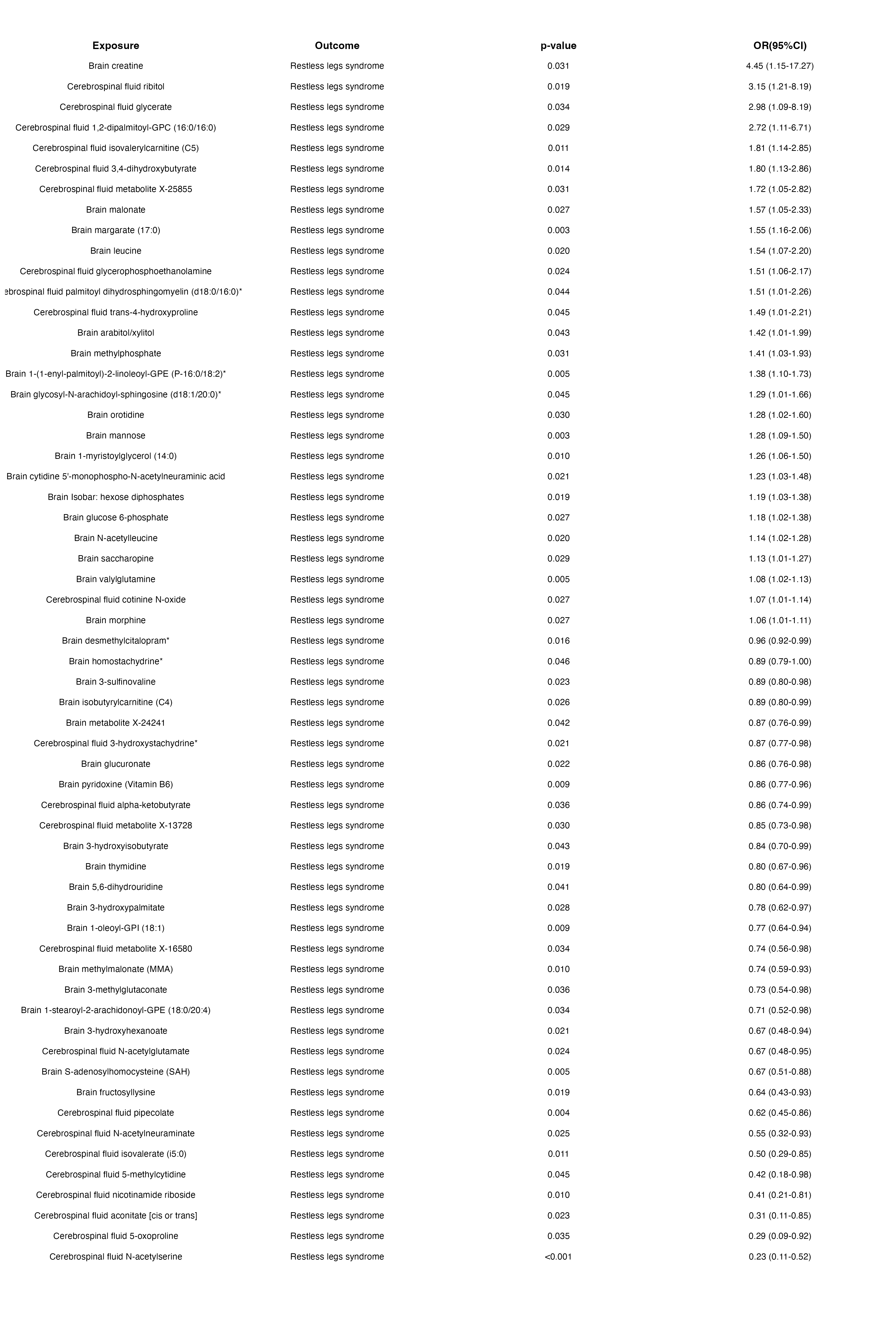

### visualization_volcano_plot.png

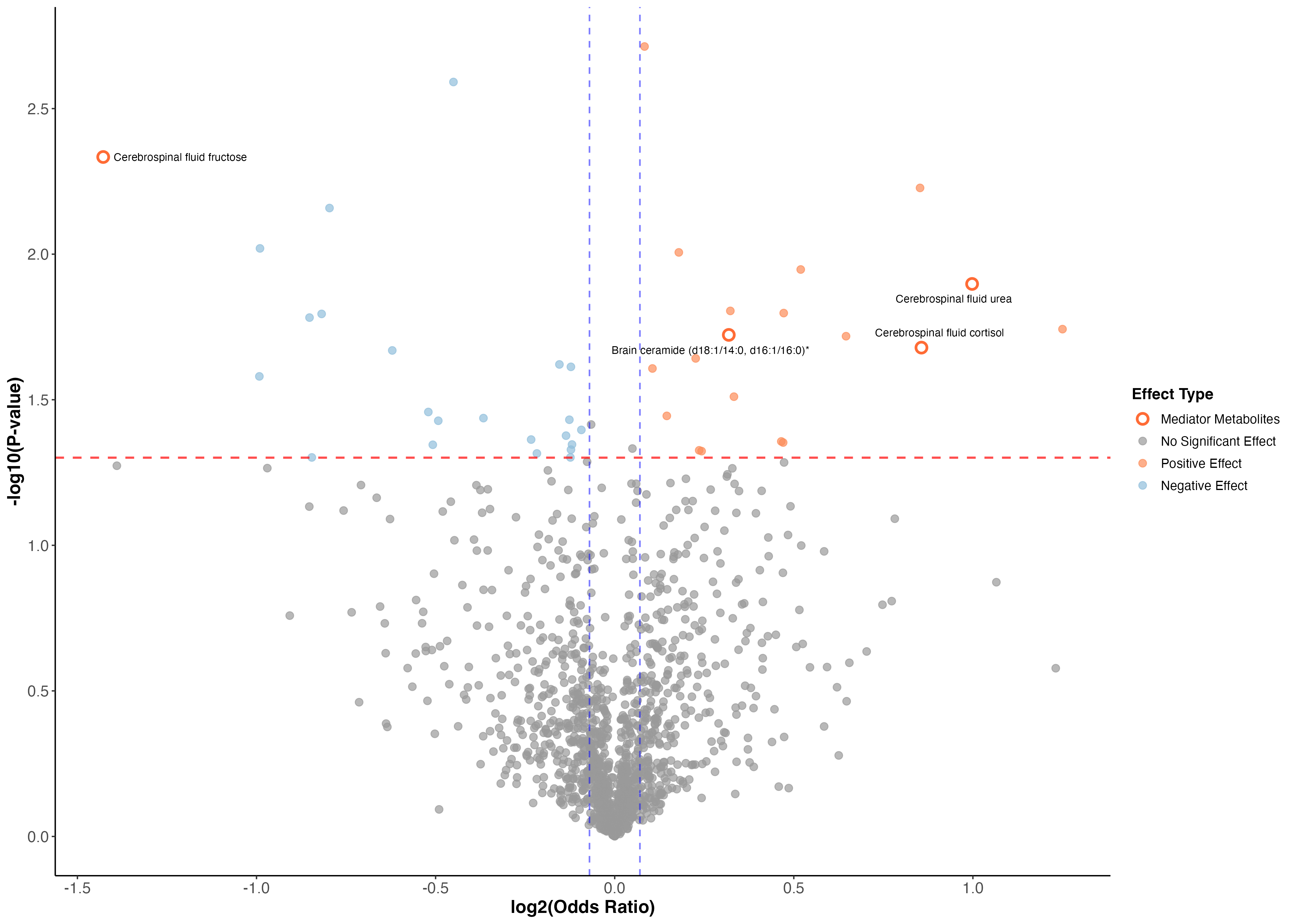

### visualization_volcano_plot.png

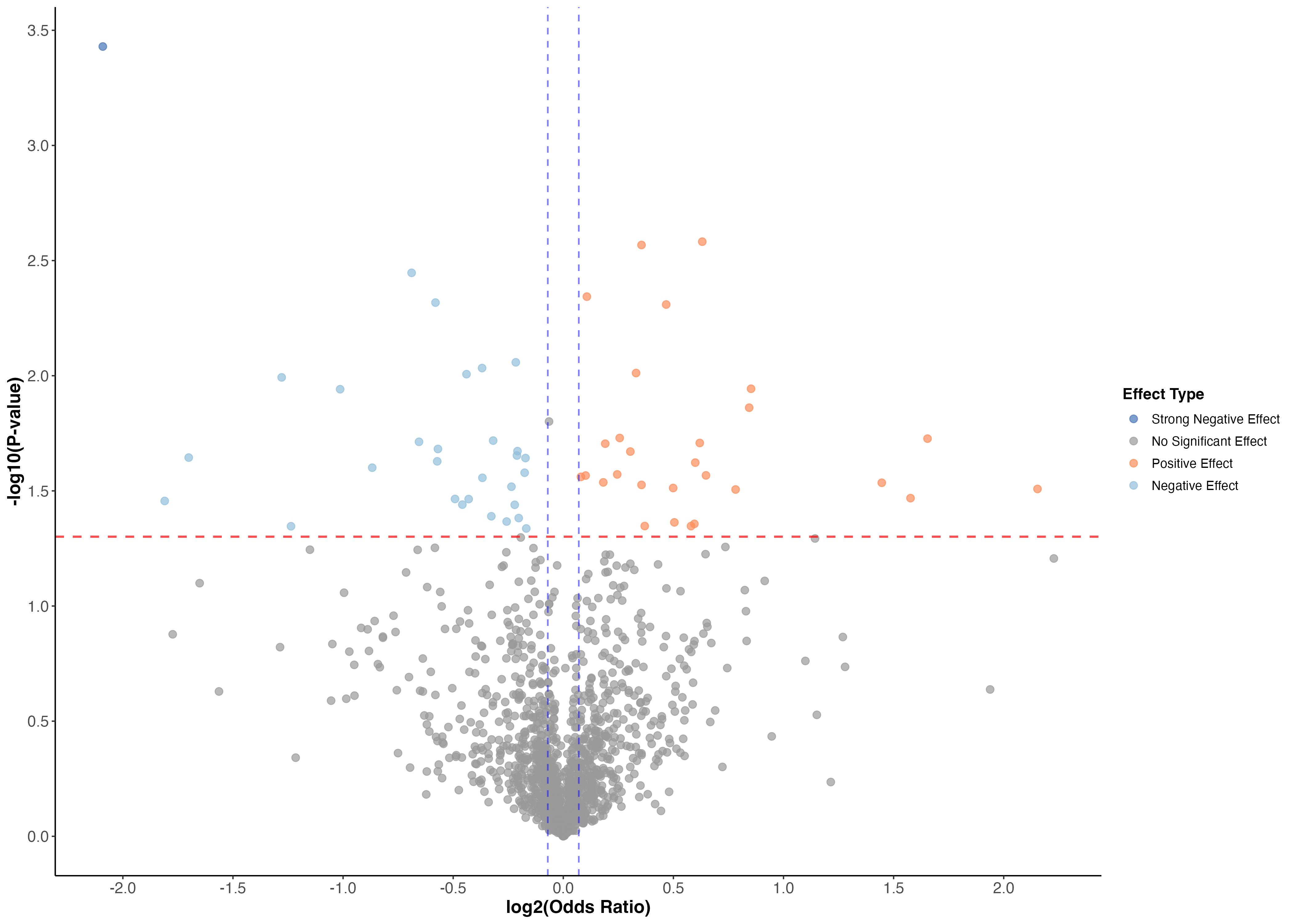
